# Automated, standardized, quantitative analysis of cardiovascular borders on chest X-rays using deep learning for assessing cardiovascular disease

**DOI:** 10.1101/2024.07.17.24310314

**Authors:** June-Goo Lee, Tae Joon Jun, Gyujun Jeong, Hongmin Oh, Sijoon Kim, Heejun Kang, Jung Bok Lee, Hyun Jung Koo, Jong Eun Lee, Joon-Won Kang, Yura Ahn, Sang Min Lee, Joon Beom Seo, Seong Ho Park, Min Soo Cho, Jung-Min Ahn, Duk-Woo Park, Joon Bum Kim, Cherry Kim, Young Joo Suh, Iksung Cho, Marly van Assen, Carlo N. De Cecco, Eun Ju Chun, Young-Hak Kim, Dong Hyun Yang, the ADC Investigators

## Abstract

**OBJECTIVE:** The analysis of cardiovascular borders (CVBs) on chest X-rays (CXRs) has traditionally relied on subjective assessment, and the cardiothoracic (CT) ratio, its sole quantitative marker, does not reflect great vessel changes and lacks established normal ranges. This study aimed to develop a deep learning-based method for quantifying CVBs on CXRs and to explore its clinical utility.

**DESIGN:** Diagnostic/prognostic study

**SETTING:** Pre-validated deep learning for quantification and z-score standardization of CVBs: the superior vena cava/ascending aorta (SVC/AO), right atrium (RA), aortic arch, pulmonary artery, left atrial appendage (LAA), left ventricle (LV), descending aorta, and carinal angle.

**PARTICIPANTS:** A total of 96,129 normal CXRs from 4 sites were used to establish age- and sex-specific normal ranges of CVBs. The clinical utility of the z-score analysis was tested using 44,567 diseased CXRs from 3 sites.

**MAIN OUTCOMES MEASURES:** The area under the curve (AUC) for detecting disease, differences in z-scores for classifying subtypes, and hazard ratio (HR) for predicting 5-year risk of death or myocardial infarction.

**RESULTS:** A total of 44,567 patients with disease (9964 valve disease; 32,900 coronary artery disease; 1299 congenital heart disease; 294 aortic aneurysm; 110 mediastinal mass) were analyzed. For distinguishing valve disease from normal controls, the AUC for the CT ratio was 0.79 (95% CI, 0.78-0.80), while the combination of RA and LV had an AUC of 0.82 (95% CI, 0.82-0.83). Between mitral and aortic stenosis, z-scores of CVBs were significantly different in LAA (1.54 vs. 0.33, p<0.001), carinal angle (1.10 vs. 0.67, p<0.001), and SVC/AO (0.63 vs. 1.02, p<0.001), reflecting distinct disease pathophysiology (dilatation of LA vs. AO). CT ratio was independently associated with a 5-year risk of death or myocardial infarction in the coronary artery disease group (z-score ≥2, adjusted HR 3.73 [95% CI, 2.09-6.64], reference z-score <-1).

**CONCLUSIONS:** Fully automated, deep learning-derived z-score analysis of CXR showed potential in detecting, classifying, and stratifying the risk of cardiovascular abnormalities. Further research is needed to determine the most beneficial clinical scenarios for this method.

**What is already known on this topic?:** Previous deep learning research in the diagnosis of cardiovascular disease using chest X-rays has focused on predicting specific disease categories, forecasting cardiovascular outcomes, and automatically measuring the cardiothoracic (CT) ratio. The end-to-end learning methods that predict disease categories or outcomes are typically limited to specific conditions and often lack explainability. While the CT ratio is traditionally used in chest X-ray analysis, it often lacks well-defined normal ranges and may not effectively detect conditions such as aortic dilatation or pulmonary trunk enlargement.

**What this study adds:** To the best of our knowledge, this is the first study to propose age- and sex-specific normal values for all cardiovascular borders (CVBs) as well as the CT ratio. Utilizing 96,129 normal chest X-rays from multiple centers, we have established normal ranges for CVBs and standardized these values into z-score mapping. This approach simplifies and enhances the practicality of clinical application. The z-score mapping of CVBs has demonstrated clinical utility in diagnosing and categorizing diseases, as well as in predicting prognosis. The AI software that automatically analyzes CVBs from CXR is available for external validation and free trial use through our dedicated research website (www.adcstudy.com). This study has transformed the interpretation of cardiovascular configuration on chest X-ray from subjective expert assessments to objective, quantifiable, and standardized measurements expressed as z-scores.

## INTRODUCTION

Advancements in artificial intelligence (AI) have significantly changed the way chest X-rays (CXR) are analyzed, enabling the automatic diagnosis of diseases affecting the lungs, pleura, and bones.^1–3^ Recent studies have also demonstrated AI’s potential in cardiovascular disease for diagnosing heart failure, predicting cardiovascular disease risks, and identifying various types of valvular diseases using CXRs.^4–8^ AI systems trained to predict cardiovascular abnormalities in CXRs can provide saliency maps for their explainability, which highlight the areas focused on making diagnoses.^5^ ^7^ However, it is important to note that these heatmaps might have limitations, particularly in pinpointing specific abnormalities or diagnosing rare diseases.^9^

The cardiothoracic (CT) ratio, a traditional metric derived from CXRs, often lacks specific reference values and may not effectively reveal changes in cardiovascular borders (CVBs) such as dilatation of the aorta or pulmonary trunk.^10^ ^11^ We have developed a fully automated, deep learning-based software that analyzes CVBs comprehensively.^12^ This AI software might offer us an opportunity to establish precise normal ranges and detect various patterns of CVB enlargement associated with cardiovascular diseases. Z-scores, which represent the number of standard deviations a data point is from the mean of a normally distributed population, are frequently used to compare quantitative test results with reference data. The precise normal ranges of CVBs may help the standardization of all CVBs into simple z-scores for newly inputted CXRs. We therefore conducted the ADC (“Automated Diagnosis of Cardiovascular abnormalities using chest X-ray”) study to develop a deep learning-based method for quantifying CVBs on CXRs and to explore its clinical utility.

## METHODS

### Study design

The ADC was a retrospective, multicenter study initiated by investigators and included 140,696 CXRs from three academic centers in two countries (South Korea, USA), as well as two public US datasets.^13^ ^14^ The study protocol received ethical approval from the Institutional Review Boards of all participating institutions, and informed consent was waived for all participants (Asan Medical Center, Seoul, Korea; 2023-1001; Severance Hospital, Seoul, Korea; 4-2020-0628; Emory University, Atlanta, GA; STUDY00005513). The study design is summarized in **Figure 1**. Briefly, we utilized a pre-validated deep learning model to automatically delineate CVBs on 96,129 normal CXRs.^12^ This deep learning-based analysis enabled the quantification of CVBs and the establishment of age- and sex-specific normal ranges for both Korea and the USA. These normal ranges facilitated the standardization of individual CVBs into simple z-scores for newly inputted CXRs (**Figure 2**). The clinical utility of the z-score mapping was evaluated across various disease groups, including valvular heart disease (VHD), coronary artery disease (CAD), congenital heart disease (CHD), aortic aneurysm, and mediastinal mass.

**Figure 1.**
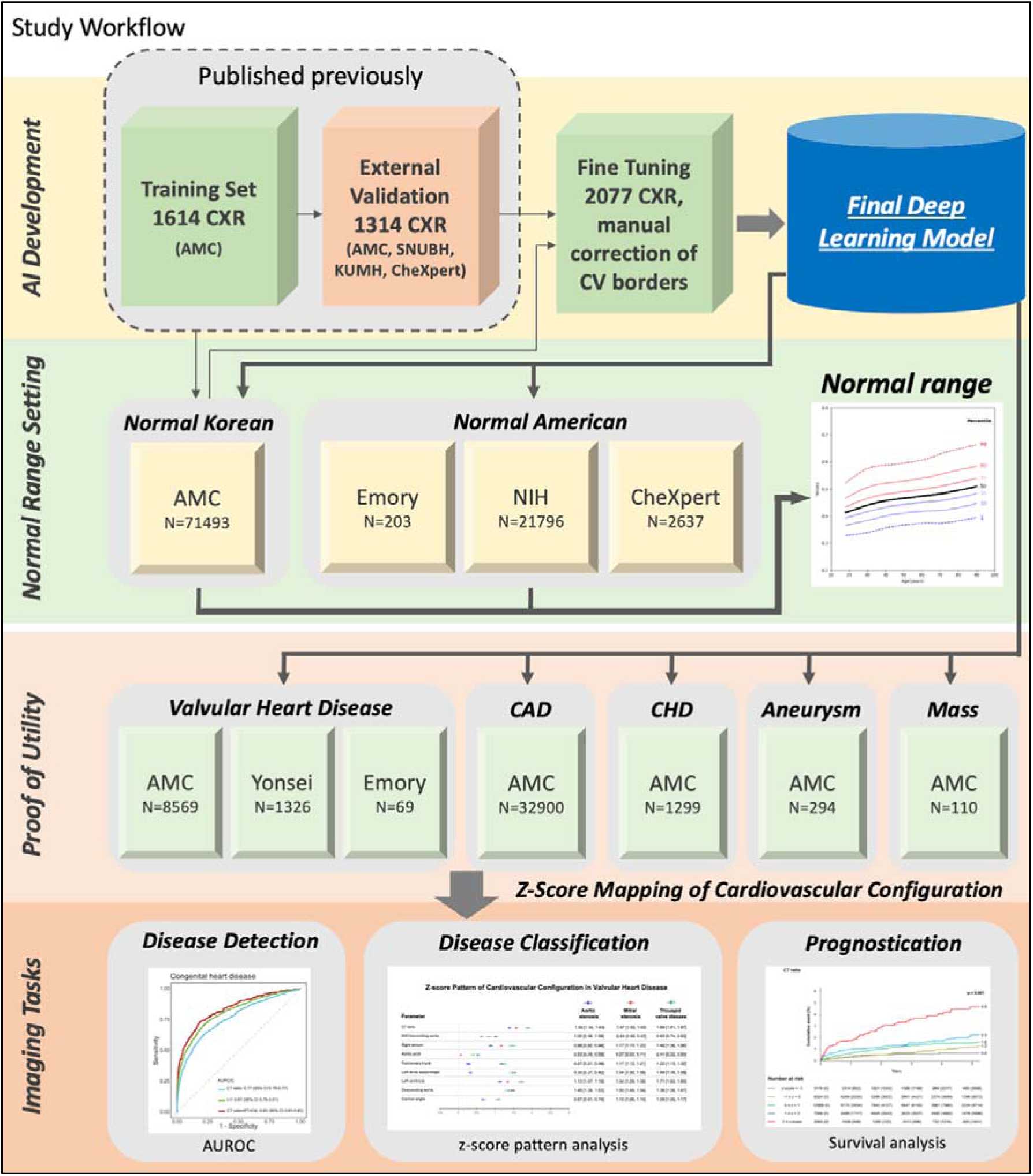
Study workflow. AI=artificial intelligence. AMC=Asan Medical Center. CAD=coronary artery disease. CHD=congenital heart disease. CheXpert=Chest eXpert (public dataset from Stanford University Hospital). CV=cardiovascular. CXR=chest X-ray. Emory=Emory University Hospital. KUMH=Korea university Ansan Hospital. NIH=National Institute of Health Clinical Center. SNUBH=Seoul National University Bundang Hospital. Yonsei=Yonsei University Severance Hospital.

**Figure 2.**
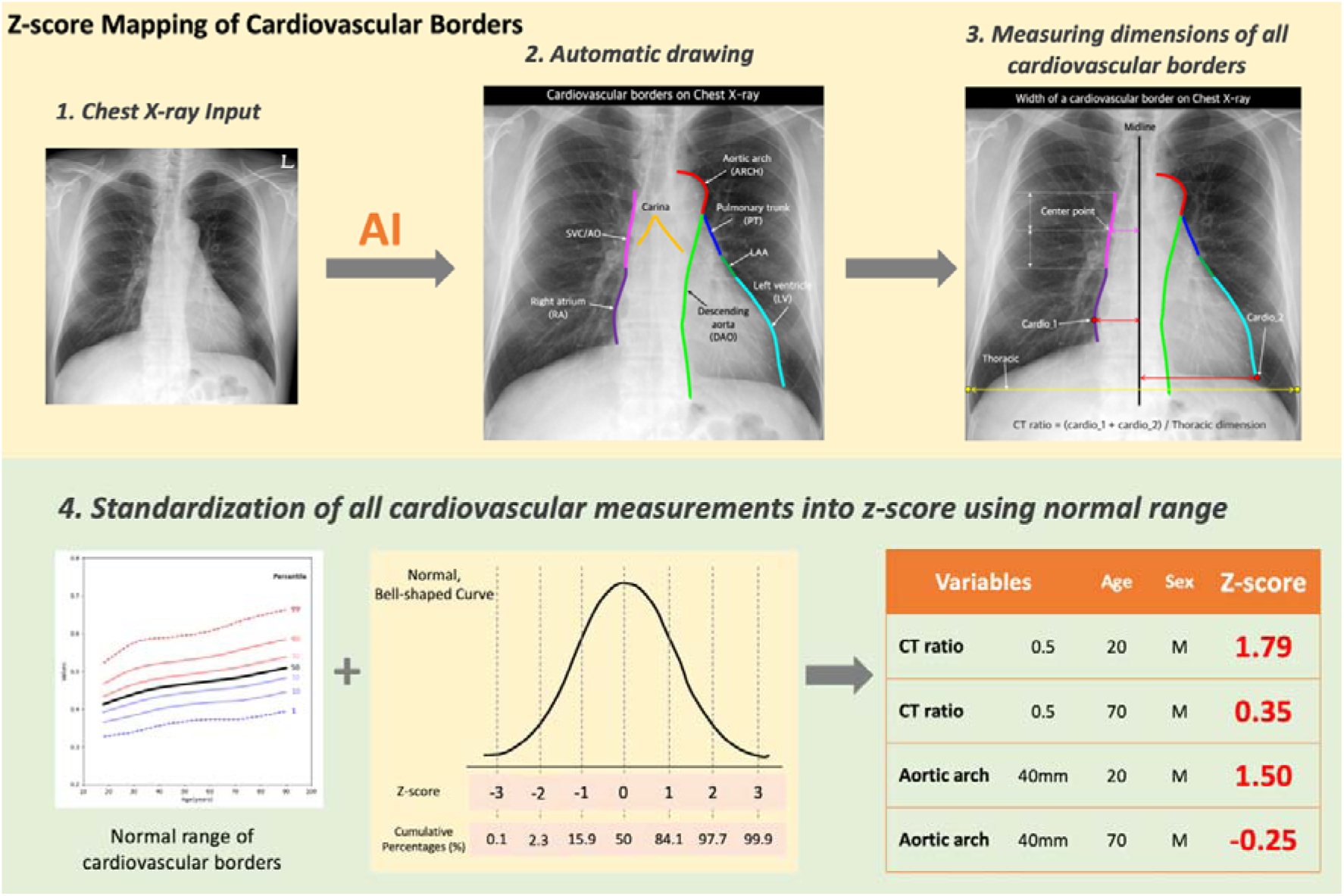
Z-score mapping process for cardiovascular borders in chest X-rays. 1. A standard postero-anterior chest X-ray is used as the input for the AI analysis. 2. AI algorithms automatically identify and delineate the CVBs on the chest X-ray. 3. The software measures the dimensions from the midline to key points on the CVBs to calculate the cardiothoracic (CT) ratio and the dimensions of individual CVBs. The width of each CVB is defined as the distance between the center points of each CVB and the midline of the CXR. The CT ratio was calculated by dividing the maximum width of the right lower CVB (corresponding to the right atrium) and the left lower cardiovascular border (corresponding to the left ventricle) by the maximal horizontal thoracic diameter. 4. The measurements are then standardized into z-scores based on the normal range, allowing for comparison according to age and sex. LAA=left atrial appendage. SVC/AO=superior vena cava/ascending aorta.

### Study cohorts

The normal cohorts used to establish reference ranges of CVBs encompassed data from Asan Medical Center (Seoul, Korea) labeled as “Normal Korean” (n=71,493) and three American datasets collectively labeled as “Normal American” (n=24,636). The dataset from Asan Medical Center spanned from 2002 to 2016, including 428,000 individuals who underwent both CXR and transthoracic echocardiography within a six-month period, with 71,493 meeting the criteria for normality in both tests (eTable 1 and eFigure 1 in Supplement). Data extraction and analysis were performed by the Big Data Research Center at Asan Medical Center utilizing the CardioNet database, a meticulously curated database integrated within the electronic health records.^15^ The selection criteria for normal CXRs involved a comprehensive review of structured echocardiography records, radiological reports, and international classification of disease codes, carefully excluding any cases indicative of cardiac, pleuropulmonary diseases, or skeletal anomalies such as scoliosis. The Normal American dataset was derived from two publicly accessible datasets—one from the National Institutes of Health Clinical Center (NIH subgroup)^13^ and another from Stanford University Hospital (CheXpert Subgroup)^14^—as well as a dataset from Emory University Medical Center, Atlanta, USA (Emory Subgroup) (eTable 2 in Supplement). For these datasets, CXRs without lung lesions and cardiomegaly were chosen after evaluations of structured radiological reports and labels. Individuals in the Emory subgroup were selected based on having normal results in both CXR and echocardiography.

The study enrolled five disease groups including the VHD group (n=9964), patients evaluated for CAD with coronary computed tomography angiography (CCTA) (CAD group, n=32,900),^16^ individuals who had undergone surgery for atrial or ventricular septal defects (CHD group, n=1299), patients confirmed with thoracic aortic aneurysms by computed tomography (Aneurysm group, n=294), and patients with biopsy-proven mediastinal masses (Mass group, n=110). The VHD group was recruited from three institutions: Asan Medical Center, Severance Hospital, and Emory University Medical Center, while the rest were from Asan Medical Center. Further details about each disease subgroup are provided in the Supplement (eTable 3-5 and eFigure 2) and summarized in **Table 1**. The VHD group was further categorized into the aortic stenosis (AS), aortic regurgitation (AR), mitral stenosis (MS), mitral regurgitation (MR), and tricuspid valve (TV) subgroups. The CAD group data, which was used for the prognostication testing in this study, included a median follow-up of 2.9 years (interquartile range, 1.0–4.5) and was segmented into significant CAD subgroups based on >50% stenosis observed in CCTA.^16^ The primary long-term clinical outcome was the composite of death from any cause or myocardial infarction at 5 years after CCTA.^16^ The Aneurysm group was composed of patients with an ascending aorta greater than 4.5 cm or a descending aorta/arch larger than 4 cm as confirmed by CT. The Mass group retrospectively enrolled patients with mediastinal masses confirmed by CT-guided biopsy.

**Table 1.**
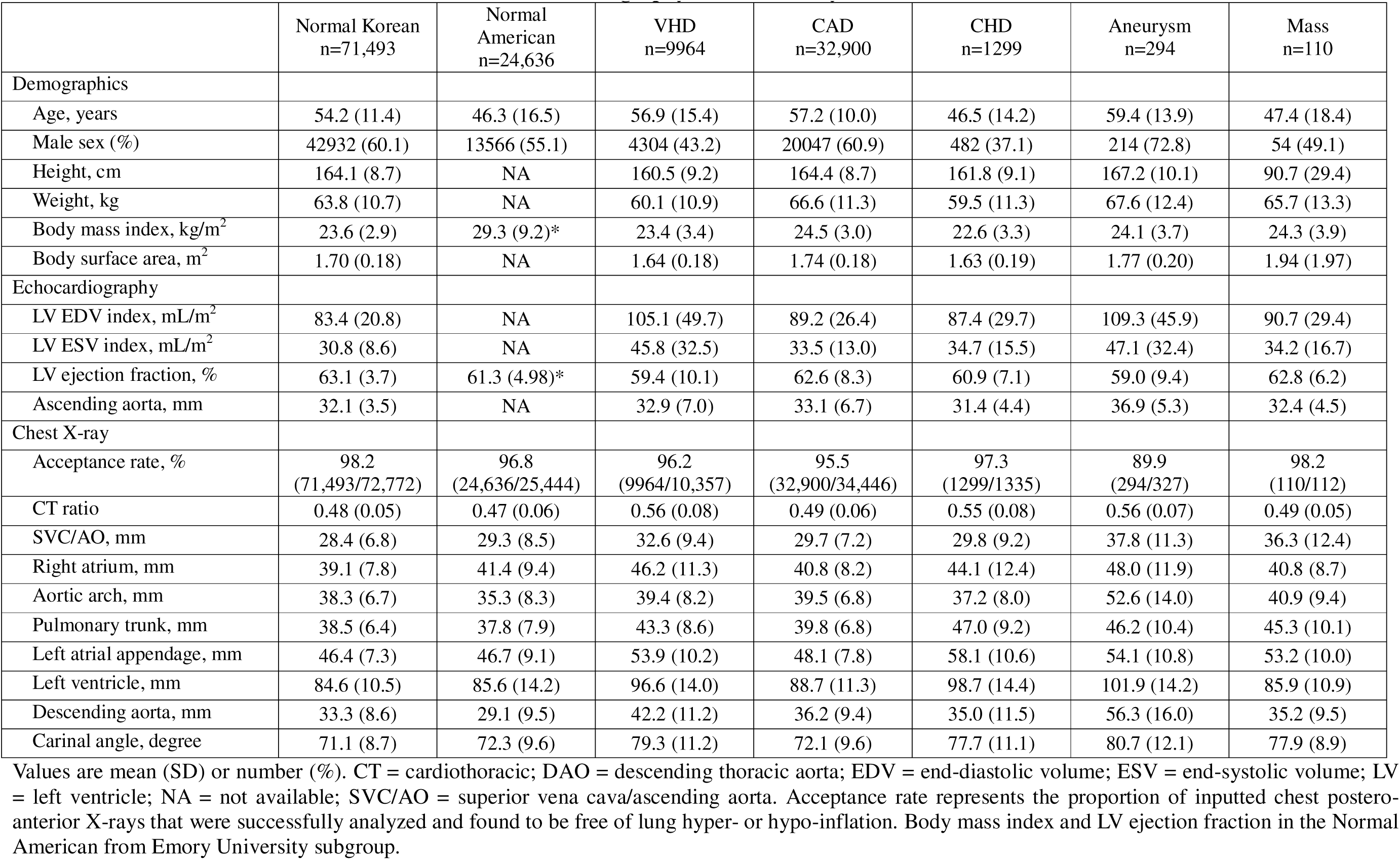
Baseline characteristics and measurements of echocardiography and chest X-ray.

### AI model

The CVB analysis software has been previously validated against multi-institutional datasets.^12^ This AI software automatically delineates each CVB when a CXR is inputted. The width of each CVB was calculated by measuring the distance from the midline of the CXR to the centerpoint of the height (**Figure 2**). Each CVB was named based on its normal anatomical location as follows: superior vena cava/ascending aorta (SVC/AO), right atrium (RA), aortic arch (Arch), pulmonary trunk (PT), left atrial appendage (LAA), left ventricle (LV), descending aorta (DAO), and the carinal angle (the angle between the lower borders of the right and left main bronchi). The definitions of each CVB are detailed in eTable 6. For CVB analysis, only CXRs taken in the postero-anterior direction with the patient standing and with proper lung inflation were analyzed. Therefore, a separate imaging filter was developed to exclude inappropriate CXRs. Detailed information on the deep learning algorithm and imaging analysis workflow is provided in the eMethods (eFigure 3 in Supplement). This AI model is available for external validation and public use via our non-commercial research website (www.adcstudy.com), which provides real-time CXR analysis capabilities (eFigure 4 in Supplement).

### Analysis of AI measurements

While most of the extracted CVB metrics approximated a symmetrical distribution, some variations in kurtosis across different metrics as well as skewness in DAO were noted (eFigure 5 and 6 in Supplement). To account for these discrepancies, each CVB metric underwent a transformation to a Box-Cox normal distribution using Generalized Additive Models for Location, Scale, and Shape.^17^ Percentile curves were plotted for individual measurements, and z-scores were computed.^17^ ^18^ Then, the dimensions of each CVBs were standardized into z-scores.

### Statistical analysis

Continuous variables are presented as means and standard deviations, while categorical variables are presented as counts and percentages. Z-scores for each disease group are shown along with their means and 95% confidence intervals (CIs). The diagnostic performance of CVB metrics in detecting specific diseases was evaluated using the area under the receiver operating characteristic (AUC), calculated with the pROC package (version 1.18.5) and included sensitivity, specificity, accuracy, positive predictive value (PPV), and negative predictive value (NPV) with cut-off point determined by the maximum Youden index. Diagnostic performance was assessed for the VHD, CAD, and CHD groups, as well as for subgroups within VHD. For each disease category, a control group three times the size of the disease group was randomly selected from the Normal Korean cohort. Multivariate logistic regression analysis was used to identify CVBs significantly associated with the presence of disease. Only CVB metrics that demonstrated a p-value <0.01 in univariate analysis and had low inter-correlations (r<0.2) were included in the multivariate analysis. The multivariate model was developed using 60% of the randomly divided data and validated using the remaining 40%.

For the CAD group, Kaplan–Meier survival analyses were conducted using the survival package (version 3.5.5), and Cox proportional-hazards regression models were used to examine the relationship between CVB z-scores and patient outcomes, independent of known cardiovascular risk factors. These analyses focused on the composite outcome of death from any cause or myocardial infarction following CCTA. The Framingham Risk Score, body mass index (BMI), the presence of diabetes mellitus, estimated glomerular filtration rate, symptoms at CCTA, and obstructed CAD (defined as ≥50% diameter stenosis) on CCTA were incorporated into the multivariate regression models, consistent with previously published results.^16^ The CVB z-scores were categorized as follows: z-score < -1, -1 ≤ z-score < 0, 0 ≤ z-score < 1, 1 ≤ z-score < 2, and z-score ≥ 2.

## RESULTS

### Study population

The study population comprised 96,129 individuals in the normal cohorts and 44,567 patients in the disease cohorts (**Table 1**, **Figure 1**). The mean age ranged from 46.5 years in the CHD group to 59.4 years in the Aneurysm group. The VHD group included 1432 AS (14.4%), 1756 AR (17.6%), 2897 MS (29.1%), 2971 MR (29.8%), 785 TV disease (7.9%), 72 PV disease (0.7%), and 51 multi-valve disease (0.5%) cases (eTable 7 in Supplement). Echocardiography results show LV ejection fraction and other cardiac dimensions, with disease groups often showing enlarged measurements compared to normal.

### Normal range of CVBs

eTable 8 in Supplement summarizes the normal ranges for CVBs on postero-anterior CXR for different age groups in both Korean and American populations according to sex. **Figure 3** presents a set of graphs depicting age-related percentile curves for various CVBs in normal individuals; detailed graphs for Normal American and Korean cohorts were provided in the eFigure 7-10 in Supplement. For both populations, the CT ratio tends to increase with age; similarly, the diameters for SVC/AO, RA, Arch, LV, and DAO also increased with age, reflecting physiological changes in the cardiovascular system as age advances. Inter-cohort comparisons revealed slightly larger CVBs in the American group, differences that were mitigated after adjusting for CT ratio.

**Figure 3.**
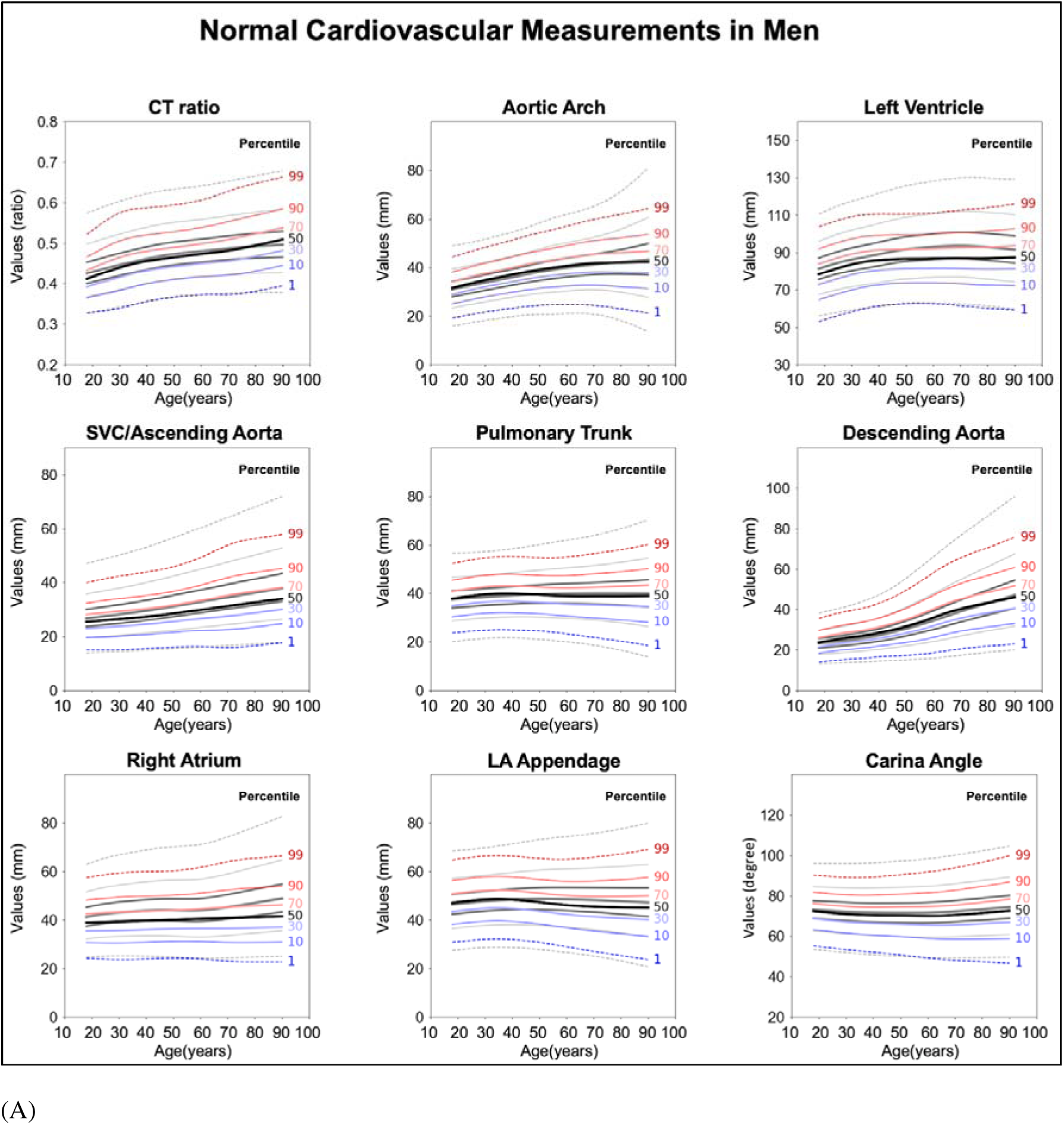
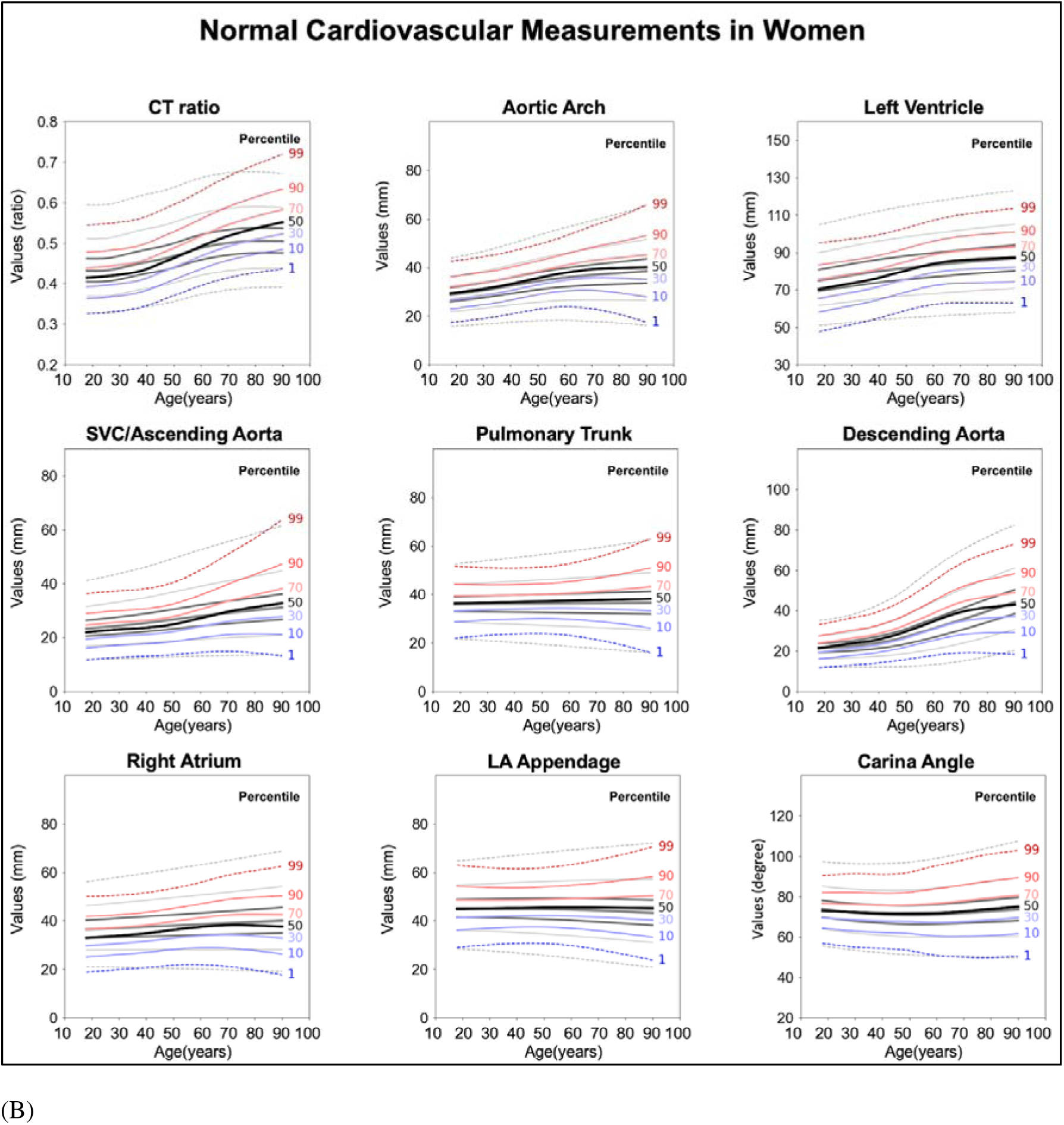
Age-related percentile curves for cardiovascular borders in normal chest X-ray. Percentile graphs of cardiovascular borders according to age for normal men (A) and women (B). The data in red or blue represents Normal Korean individuals, and the data overlapped in gray represents that of Normal Americans.

### Z-scores of CVBs in disease groups

In the analysis of disease groups, z-scores for CVB were generally elevated, with the VHD and CHD groups displaying significantly higher z-scores compared to the CAD group (**Figure 4** and eTable 9 in Supplement). Specifically, the mean z-scores for the CT ratio were 0.39 in CAD, 1.27 in CHD, and 1.40 in VHD. **Figure 4** highlights the variations in z-scores across diseases, showcasing the disease-specific changes in CVB parameters. MS, often accompanied by left atrial enlargement, showed marked increases in the LAA (z-score=1.54) and carinal angle (z-score=1.10) as a result of the left atrial pushing upwards; this was in marked contrast to AS where the increase in the SVC/AO (z-score=1.02) indicated dilation of the ascending aorta. In CHD, including atrial or ventricular septal defects, the z-score of the aortic arch (0.01) was relatively low, reflecting the reduced cardiac output of the left heart due to left-to-right shunt disease. The aortic aneurysm group showed significant increases in the arch (1.95) and DAO (2.65) z-scores, indicating aneurysmal changes. Mediastinal mass conditions also demonstrated elevated z-scores, especially for the SVC/AO (1.04) and the pulmonary trunk (1.03), which may indicate a mass shadow or compression caused by the tumor.

**Figure 4.**
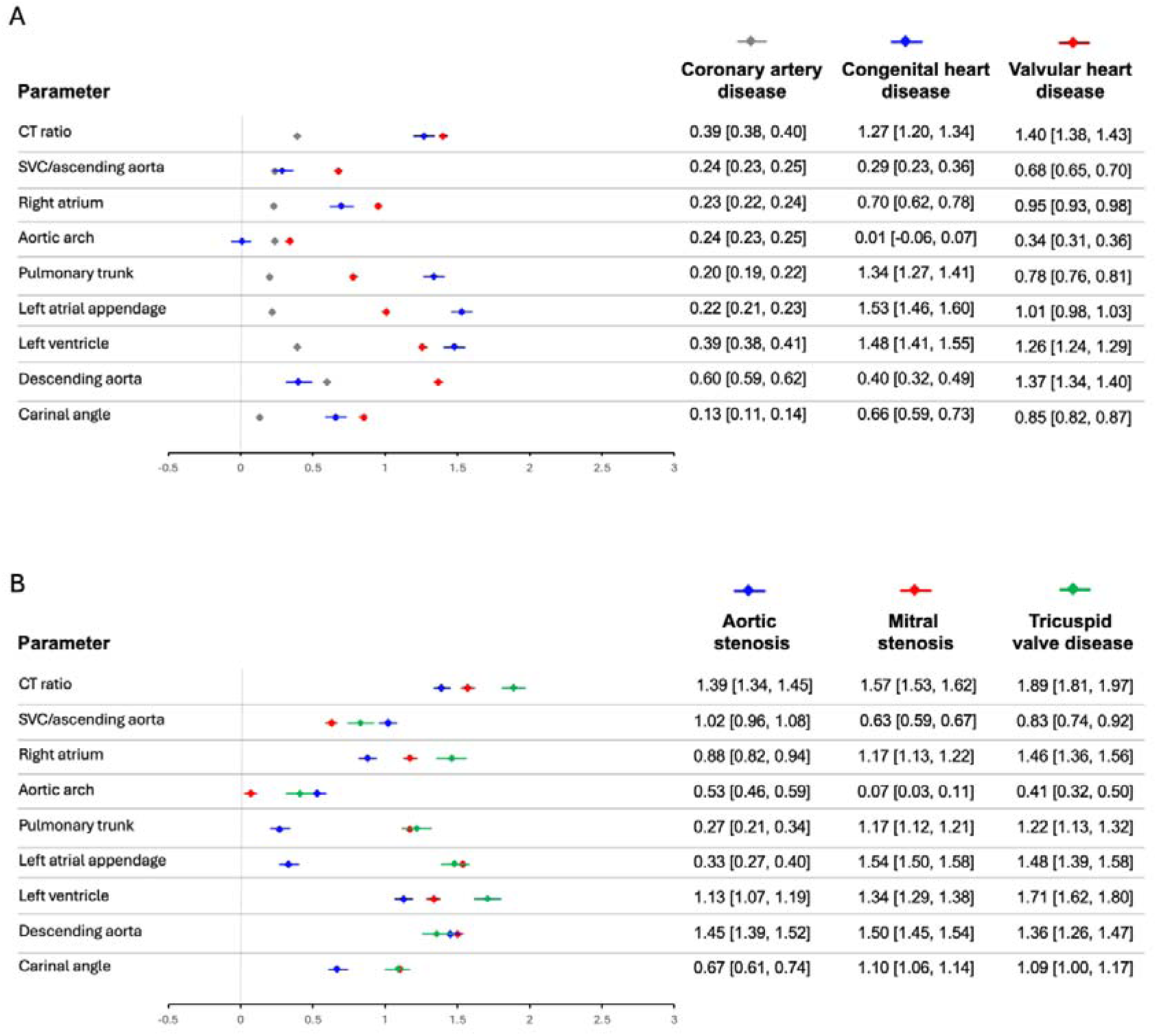
Comparative z-score forest plot for disease classification. Each parameter is represented by a horizontal line, with data points indicating the mean z-score and error bars showing 95% confidence intervals. (A) Comparison across different disease groups: coronary artery disease, congenital heart disease, and valvular heart disease. (B) Comparison across specific valvular heart disease: aortic stenosis, mitral stenosis, and tricuspid valve disease.

### Diagnostic performance

The diagnostic evaluation of CVBs highlighted the CT ratio z-score as a robust metric across VHD, CAD, and CHD groups (**Figure 5**). The AUC for detecting VHD using the CT ratio reached 0.79 (95% CI, 0.78–0.80), which was increased to 0.82 (95% CI, 0.82–0.83) when combined with RA and LV metrics. CHD detection benefited from a CT ratio AUC of 0.77 (95% CI, 0.74–0.79), which improved to 0.81 (95% CI, 0.79–0.84) when PT and carinal angle were added. Among the subgroups of VHD, TV disease detection had the highest AUC of 0.87 (95% CI, 0.84–0.89) using the CT ratio. Detailed information on demographics, AUC, sensitivity, specificity, cut-off, PPV, and NPV is provided in eTable 10-17 in Supplement.

**Figure 5.**
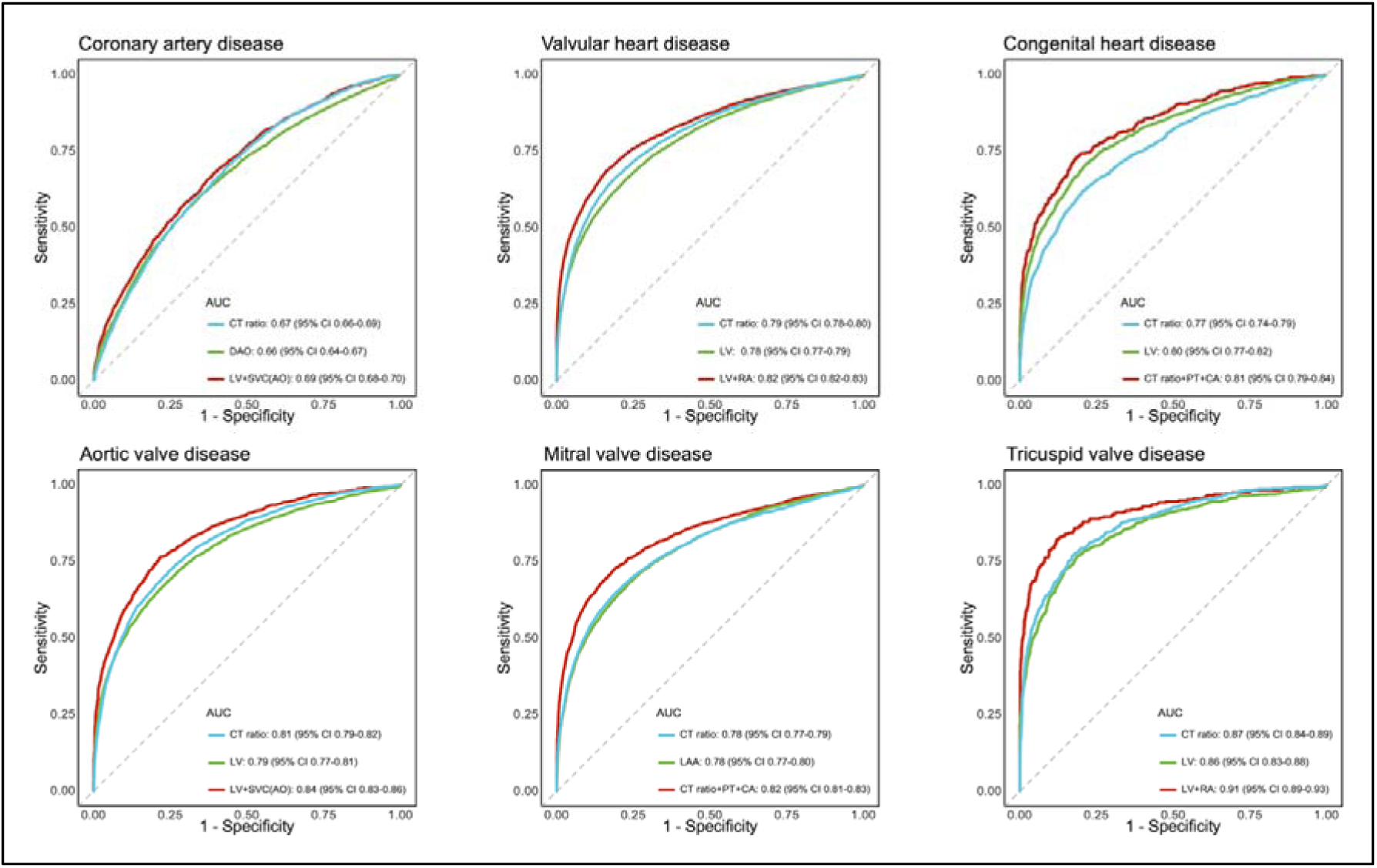
Performance of the z-score mapping of cardiovascular borders for the detection of cardiovascular disease. AUC=area under the receiver operating characteristic curve. CA=carinal angle. CT=cardiothoracic. DAO=descending aorta. LAA=left atrial appendage. LV=left ventricle. PT=pulmonary trunk. RA=right atrium. SVC=superior vena cava (SVC/aorta).

### Prognostic value

In the cohort of 32,900 CAD patients, there were 390 (1.18%) instances of all-cause death or myocardial infarctions. CT ratio z-scores indicated an increasing risk with higher scores (**Figure 6**). Patients with a CT ratio z-score of 2 or higher were at a significantly elevated risk (adjusted HR 3.73, 95% CI, 2.09–6.64), showing a higher percentage of cumulative events (4.6% vs. 0.6%, p<0.001) over 5 years compared to the reference group with a z-score less than -1 (HR 1.00). Elevated risks were also observed with higher z-scores (≥2) for SVC/AO, RA, DAO, and carinal angle, while the Arch, PT, LAA, and LV z-scores not reaching statistical significance (eTable 18 and eFigure 11-18 in Supplement).

**Figure 6.**
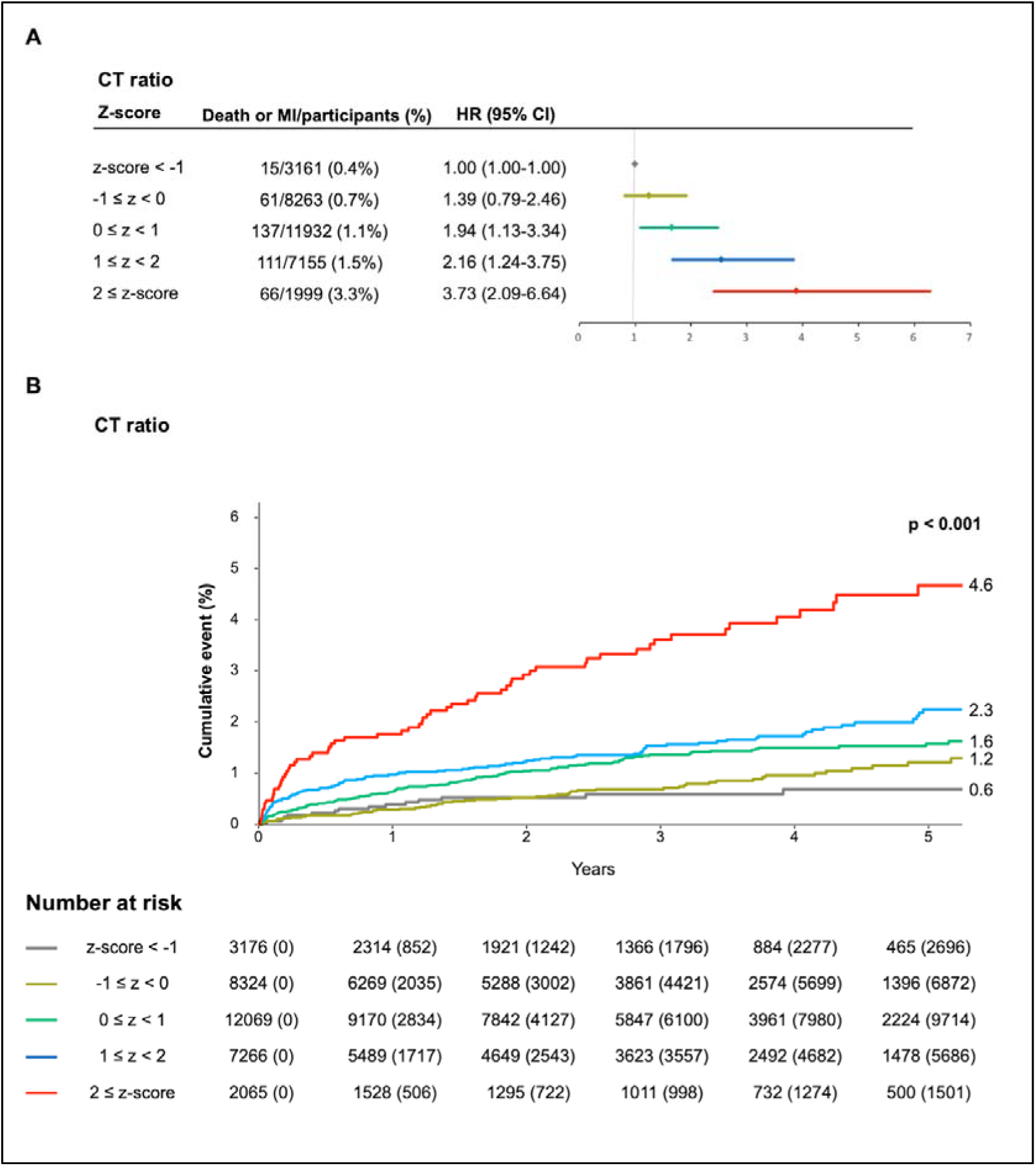
All-cause death or myocardial infarction stratified by cardiothoracic ratio z-score. In the coronary artery disease group, all-cause death or myocardial infarction were stratified by z-score categories of the CT ratio. Adjusted HRs were compared with the lowest z-score group (<-1). (A) Percent of death or myocardial infarction and adjusted HR increased across ascending z-score categories. (B) Cumulative event rate for each z-score category of the CT ratio during a follow-up duration of 5 years. CT=cardiothoracic. HR=hazard ratio.

### Case examples

We presented eight CXR case examples (eFigure 19-26 in Supplement), illustrating the application of z-score mapping in diagnosing various cardiomediastinal diseases. The cases span a range of conditions, including AS, MS, AR, atrial septal defect, aortic aneurysms, and mediastinal masses.

## DISCUSSION

In the ADC study, we established normal values for CVBs and introduced a new methodology for utilizing CXRs in cardiovascular disease diagnosis. Our main findings are as follows. First, z-score mapping for CVBs was feasible in disease diagnosis. In certain cases, combining different CVBs enhanced diagnostic accuracy beyond the CT ratio. Second, variations in z-scores, reflecting the underlying disease pathophysiology, indicate that CXRs could be useful in classifying diseases, such as distinguishing between aortic and mitral valve diseases. As demonstrated through our case presentations, the changes in individual CVB z-scores may be correlated with the pathophysiological changes observed in patients’ echocardiograms or CT scans. The z-score mapping allows for a more objective and quantifiable method of interpretation compared to traditional approaches to CXR analysis. Lastly, measures of CVB, including the CT ratio, showed potential in predicting clinical outcomes, adding value to traditional risk scoring systems.

Regarding the quantitative analysis of CXR, previous studies have focused on automatically extracting the CT ratio^19–22^ and biological age^23^ from CXRs using AI. As demonstrated in the ADC study, the variability of the CT ratio’s normal values based on age and sex indicates limitations in applying a single cutoff 0.5. Moreover, conditions such as pulmonary trunk and ascending aortic dilatation cannot be adequately assessed by the CT ratio alone. The significance of this ADC study lies in standardizing various CVBs into a single parameter of z-score, not just the CT ratio, particularly showing some success in making differential diagnoses that were not previously possible with the CT ratio. Extracting biological age from CXR has shown promising prognostic value when added to existing cardiovascular risk matrices, offering a potential new utility for CXR.^23^ Since, CXR-derived biological age and CVB z-scores are numerical data and likely independent, combining them could offer potential for clinical practice and research applications.

The use of “end-to-end” supervised learning, where AI directly learns from CXRs with abnormalities compared to a control group, is a widely adopted approach in current AI research. This method has been extensively applied in the field of cardiovascular disease to predict conditions such as acute chest pain syndrome^24^, aortic dissection^25^, LV systolic dysfunction^6^, structural LV disease^7^, valvular heart disease^5^, aortic stenosis^26^, and atrial fibrillation^27^ using CXRs. Other studies have also tried to predict the 10-year risk for major adverse cardiovascular events using CXRs.^8^ These studies often employ saliency maps to improve the explainability of AI, indicating the specific areas of the image that the AI prioritized to reach its decision. However, saliency maps can struggle with the precise localization of abnormalities and may pose interpretative challenges when applied to diseases not included in the algorithm’s training.^9^ Z-score mapping, by providing interpretable numerical values independent of specific diseases, can help overcome these limitations, offering broader applicability across various cardiomediastinal conditions. This advancement may offer a modernized approach to interpreting CXRs, aligning with clinicians’ preference for quantifiable metrics, such as blood tests and echocardiographic parameters. Moreover, this numerical approach facilitates a more objective comparison during the follow-up of CXRs, making it easier to interpret changes over time in a patient’s condition.

For the utilization of z-score mapping of CXR in real-world clinical practice, it is crucial to establish the most appropriate clinical application scenarios. For example, z-score mapping of CXRs could serve as a gatekeeper before proceeding to more costly and complex tests such as echocardiography. Another promising scenario could involve using z-score mapping of CXRs as a screening tool to detect left-to-right shunt diseases before they progress to irreversible pulmonary hypertension. Such applications could significantly enhance the utility of CXR, providing a cost-effective, accessible, and non-invasive method. Particularly, using CXRs for VHD or CHD in screening scenarios could be a viable alternative in underdeveloped countries where healthcare infrastructure is insufficient.^28^

This study has the following limitations: First, the CVB analysis is subject to limitations of the CXR modality compared to echocardiography or CT. As demonstrated in Case 4 (ASD) and Cases 6 and 7 (mediastinal mass), CVBs can be influenced by adjacent structures. Therefore, the interpretation of CVB analysis must be based on understanding of the specific disease’s pathophysiology and topographical anatomical knowledge in CXR. Second, although this study presents diagnostic performance, z-score pattern analysis, and prognostic value, it has not provided definitive cut-off values refined enough for application in actual practice. This is because, although the normal ranges and disease cohorts included data from multiple institutions, they did not encompass a wide variety of ethnicities and real-world conditions, including disease groups. Future research should conduct more extensive studies across a wide range of clinical application scenarios.

The ADC study has introduced a fully automated, deep learning-derived z-score analysis of CXR showed potential in detecting, classifying, and stratifying the risk of cardiovascular abnormalities.

Further research is needed to determine the most beneficial clinical scenarios for this method.

## Data Availability

All data produced in the present study are available upon reasonable request to the authors

https://www.adcstudy.com

## Acknowledgements

Professor Tae-Hwan Lim provided critical feedback, expertise, and encouragement for the ADC study.

## Author contributions

DHY, SHP, and YHK conceived and designed the study. JGL, GJ, and HO developed and applied the deep learning model. TJJ, HJK, JEL, JWK, YA, SML, JBS, MSC, JMA, DWP, JBK, CK, YJS, IC, MA, CDC, and EJC provided datasets. SK, HK, and JBL performed statistical analyses. DHY, JGL, TJJ, and YHK drafted the manuscript. DHY and YHK supervised the study. All authors contributed to the acquisition and interpretation of data, and critical revision of the manuscript. DHY and YHK had access to and verified all the data in the study.

## Data sharing

AI software developed for this study is freely available on the ADC study website (www.adcstudy.com) for external validation. The patient-level data collected during the study will not be made publicly accessible. However, the research team is open to considering collaborative projects and specific data-sharing requests. Inquiries regarding data access should be directed to the lead investigator, Dong Hyun Yang.

