## Supplementary Appendix for "Automated, standardized, quantitative analysis of cardiovascular borders on chest X-rays using deep learning for assessing cardiovascular disease"

This appendix has been provided by the authors to give readers additional information about their work.

**TABLE OF CONTENTS**

**eMethods**

**eTable 1.** Summary of the Normal Korean cohort……………………………………….4
**eFigure 1.** Flow chart of the Normal Korean cohort……………………..........................5
**eTable 2.** Summary of the Normal American cohort……………………………………..6
**eTable 3.** Summary of the valvular heart disease group……………….............................7
**eFigure 2.** Flow chart of the valvular heart disease group (AMC)……...............................8
**eTable 4.** Summary of the coronary artery disease group……………………………….9
**eTable 5.** Summary of the congenital heart disease, aortic aneurysm, and mediastinal mass groups……………………………………………………………………………10
**AI model for analyzing cardiovascular borders on chest X-ray**………………………..11
**eTable 6.** Definitions of cardiovascular borders……………………..............................12
**eFigure 3.** Chest X-ray analysis workflow…….............................................................13
**eFigure 13.** All-cause death or myocardial infarction stratified by aortic arch in chest X-ray……..................................................................................................................40
**eFigure 14.** All-cause death or myocardial infarction stratified by pulmonary trunk in chest X-ray……...........................................................................................................41
**eFigure 15.** All-cause death or myocardial infarction stratified by left atrial (LA) appendage in chest X-ray……...........................................................................................42

**eFigure 16.** All-cause death or myocardial infarction stratified by left ventricle in chest X-ray……..............................................................................................................43
**eFigure 17.** All-cause death or myocardial infarction stratified by descending aorta in chest X-ray……........................................................................................................44

**eTable 1. Summary of the Normal Korean cohort**

| **Cohorts name** | **Normal Korean** |
|---|---|
| **Institution** | Asan Medical Center (Seoul, Korea) |
| **Providers** | Tae Joon Jun, Dong Hyun Yang, Young-Hak Kim |
| **Date** | 2000 – 2016 |
| **Subject number** | N=71493 |
| **Data building and cleansing** | - From manually curated big data platform 'CardioNet', N=42800[1]<br>- Review of disease codes (International Classification of Disease, 10$^{th}$ version, ICD-10)<br>- Review of structured report of transthoracic echocardiography<br>- Text mining in radiological report of chest X-ray<br>- Image postprocessing of chest X-ray for excluding hyper-or hypo-inflated lung and anterior-to-posterior chest X-ray |
| **Inclusion criteria** | - Subjects aged ≥ 18 who underwent thoracic echocardiography and chest X-ray within 6 months<br>- Normal results of echocardiography and chest X-ray (posterior-to-anterior) |
| **Exclusion criteria** | - Based on ICD-10: cardiac disease (e.g., valve, congenital anomaly, heart failure, arrhythmia…), malignant tumor of thorax, pleuropulmonary lesions (e.g., pneumonia, emphysema, pneumothorax…), skeletal abnormality (e.g., scoliosis)<br>- Based on structured report of echocardiography: abnormal quantitative value of cardiovascular function or dimension (e.g., ejection fraction, ventricular size)<br>- Previous cardiac operation, percutaneous coronary artery intervention<br>- Based on text mining in radiology report: atelectasis, collapse, consolidation, pneumothorax, endotracheal tube, chest tube. |

**eFigure 1. Flow chart of the normal Korean cohort**

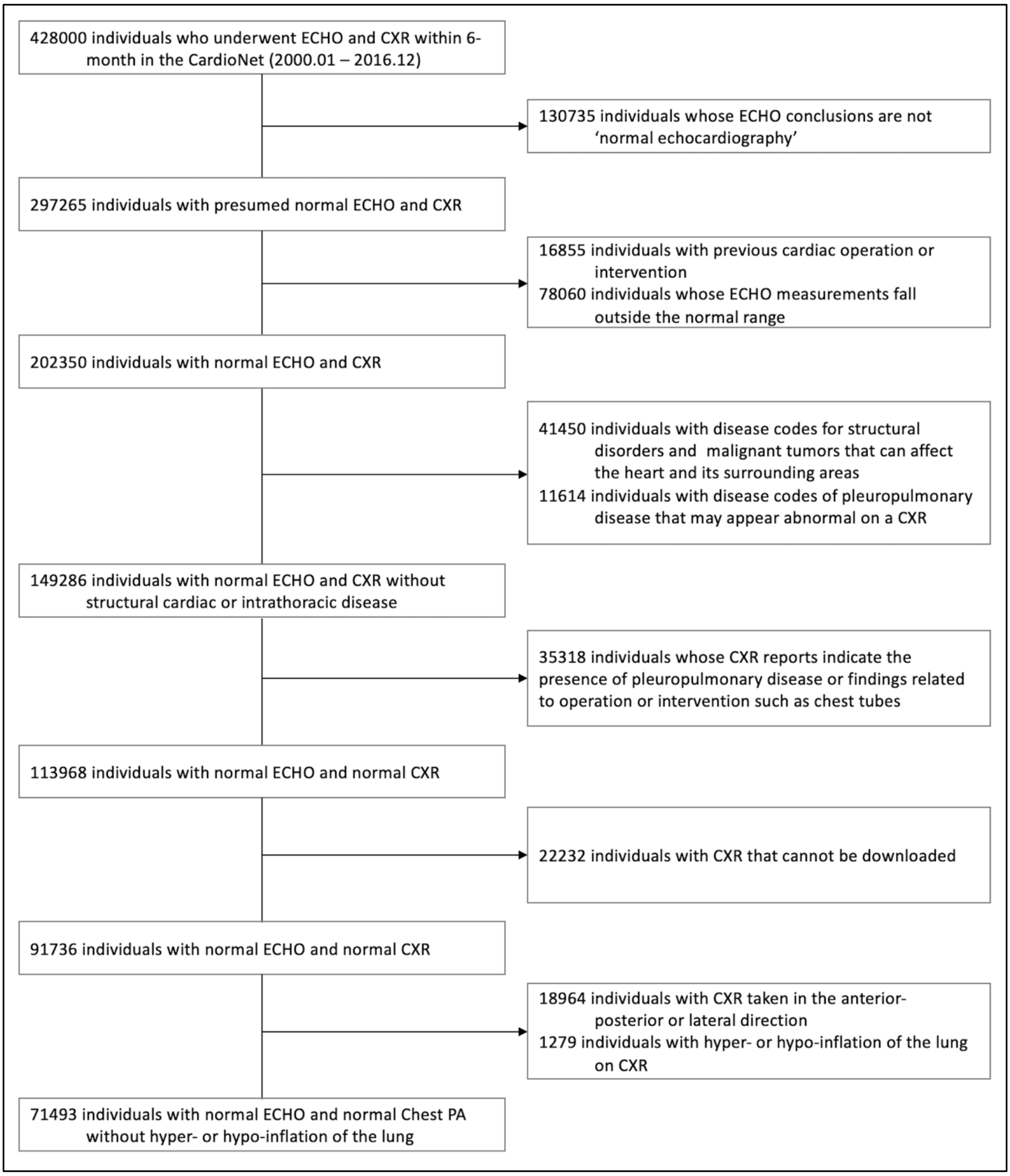

CardioNet[1] = a manually curated database integrated within the electronic health records by the Big Data Research Center at Asan Medical Center; CXR = Chest X-ray; ECHO = transthoracic echocardiography.

**eTable 2. Summary of the Normal American cohort**

| **Cohorts name** | **Normal American (Emory subgroup)** |
|---|---|
| **Institution** | Emory University Hospital, Atlanta, GA, US |
| **Providers** | Cherry Kim, Marly van Assen, Carlo N. De Cecco |
| **Date** | 2019 - 2022 |
| **Subject number** | N=203 |
| **Data building and cleansing** | - Review of structured report of transthoracic echocardiography<br>- Review of chest X-ray image and radiological report<br>- Image postprocessing of chest X-ray for excluding hyper-or hypo-inflated lung and anterior-to-posterior chest X-ray |
| **Inclusion criteria** | - Subjects aged ≥ 18 who underwent thoracic echocardiography and chest X-ray within 6 months<br>- Normal results of echocardiography and chest X-ray (posterior-to-anterior) |
| **Exclusion criteria** | - Based on structured report of echocardiography: structural cardiac disease (e.g., valve, congenital anomaly,), abnormal quantitative value of cardiovascular function or dimension (e.g., ejection fraction, ventricular size)<br>- Based on image review of chest X-ray by an expert (Cherry Kim): atelectasis, collapse, consolidation, pneumothorax, endotracheal tube, chest tube, and skeletal abnormality… |
| **Cohorts name** | **Normal American (NIH subgroup)** |
| **Institution** | Open dataset from National Institute of Health[2] |
| **Data download** | https://nihcc.app.box.com/v/ChestXray-NIHCC |
| **File format** | - a png file (there was no pixel spacing information; only calculation of relative ratio such as CT ratio was possible) |
| **Date** | 1992 – 2015 |
| **Data building and cleansing** | - Review of chest X-ray image and radiological report<br>- Image postprocessing of chest X-ray for excluding hyper-or hypo-inflated lung and anterior-to-posterior chest X-ray |
| **Subject number** | N=21796 |
| **Inclusion criteria** | - Based on structured radiological report of chest X-ray, images without lung lesion or cardiomegaly were included. |
| **Cohorts name** | **Normal American (CheXpert subgroup)** |
| **Institution** | Open dataset from the Stanford University Medical Center[3] |
| **Data download** | https://stanfordmlgroup.github.io/competitions/chexpert/ |
| **File format** | - dicom file |
| **Date** | 2002 – 2017 |
| **Data building and cleansing** | - Review of chest X-ray image and radiological report<br>- Image postprocessing of chest X-ray for excluding hyper-or hypo-inflated lung and anterior-to-posterior chest X-ray |
| **Subject number** | N=2637 |
| **Inclusion criteria** | - Based on structured radiological report of chest X-ray, images without lung lesion or cardiomegaly were included. |

**eTable 3. Summary of valvular heart disease group**

| **Cohorts name** | **Valvular Heart Disease (VHD)** |
|---|---|
| **Institution** | Asan Medical Center, Seoul, Korea (AMC)<br>Severance Hospital, Yonsei University, Seoul, Korea (Yonsei)<br>Emory University Hospital, Atlanta, GA, US (Emory) |
| **Providers** | Joom Bum Kim, Hyun Jung Koo, Joon-Won Kang, Young-Hak Kim (AMC)<br>Young Joo Suh, Iksung Cho (Yonsei)<br>Cherry Kim, Marly van Assen, Carlo N. De Cecco (Emory) |
| **Date** | 2000 – 2022 (AMC); 2000 – 2022 (Yonsei); 2019 – 2022 (Emory) |
| **Subject number** | N=9964<br>8569 (AMC), 1326 (Yonsei), 69 (Emory) |
| **Data building and cleansing** | - Review of disease codes (International Classification of Disease, 10th version, ICD-10)<br>- Review of structured report of transthoracic echocardiography<br>- Review of chest X-ray image and radiological report<br>- Image postprocessing of chest X-ray for excluding hyper-or hypo-inflated lung and anterior-to-posterior chest X-ray |
| **Inclusion criteria** | - Subjects aged ≥ 18 who underwent thoracic echocardiography and chest X-ray within 6 months<br>- Based on disease codes (ICD-10) and structured report of echocardiography: moderate or severe valvular heart disease (AMC & Emory)<br>- Based on disease codes (ICD-10) and structured report of echocardiography: moderate or severe mitral stenosis (Yonsei) |
| **Exclusion criteria** | - Previous cardiac operation<br>- Based on image review of chest X-ray by an expert: atelectasis, collapse, consolidation, pneumothorax, endotracheal tube, chest tube, and skeletal abnormality… |

**eFigure 2. Flow chart of the valvular heart disease group (AMC)**

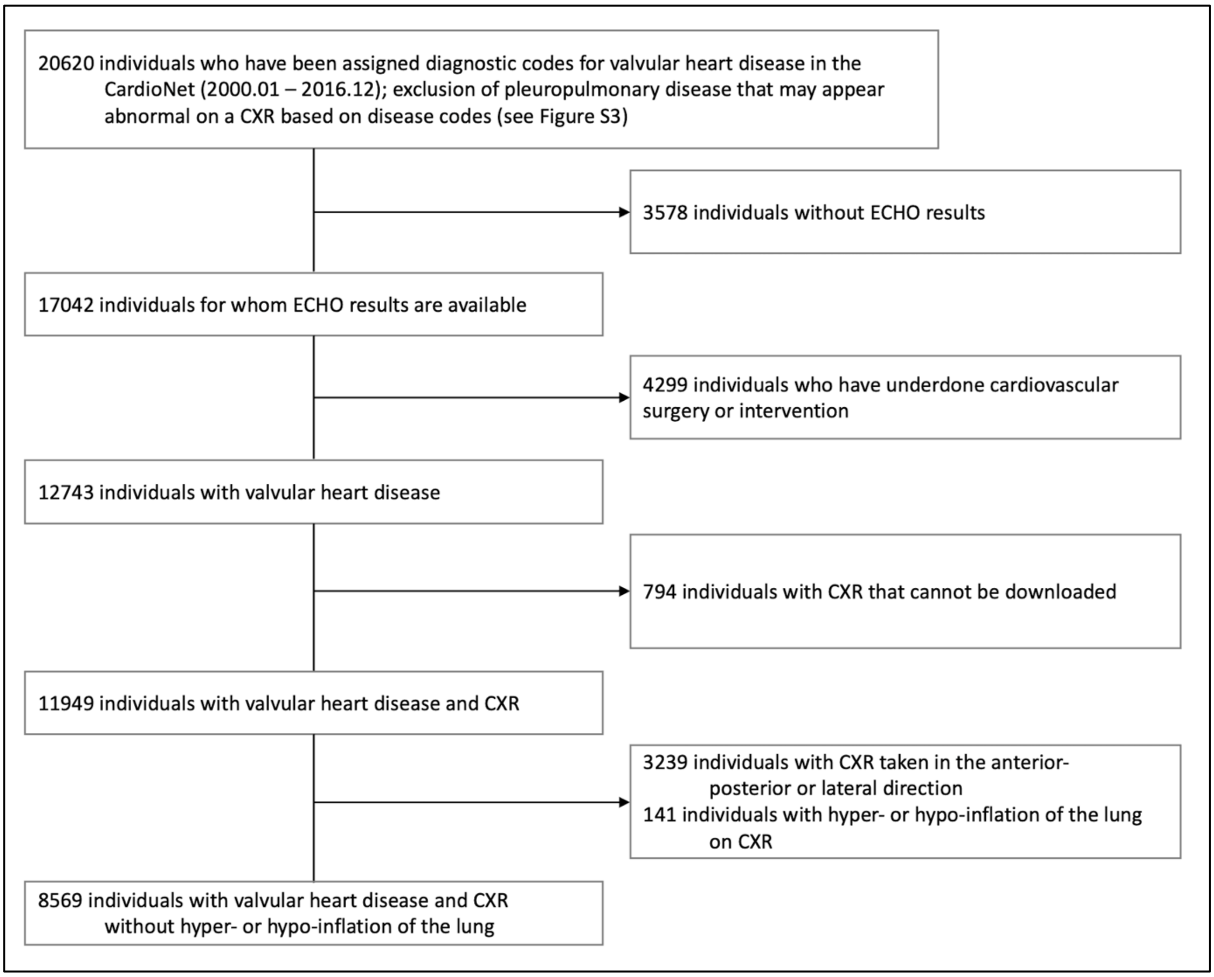

CardioNet[1] = a manually curated database integrated within the electronic health records by the Big Data Research Center at Asan Medical Center; CXR = Chest X-ray; ECHO = transthoracic echocardiography.

**eTable 4. Summary of coronary artery disease group**

| Cohorts name | Coronary Artery Disease (CAD) |
|---|---|
| **Institution** | Asan Medical Center, Seoul, Korea (AMC) |
| **Providers** | Min Soo Cho, Jung-Min Ahn, Duk-Woo Park, Joon-Won Kang |
| **Date** | 2007 - 2013 |
| **Subject number** | N=32900 |
| **Data building and cleansing** | - From previously published registry[4]: consecutive patients who underwent coronary computed tomography angiography (CCTA) without known or documented heart disease.<br>- Review of structured report of CCTA<br>- Review of chest X-ray image and radiological report<br>- Image postprocessing of chest X-ray for excluding hyper-or hypo-inflated lung and anterior-to-posterior chest X-ray |
| **Inclusion criteria** | - Subjects aged ≥ 18 who underwent CCTA and chest X-ray within 6 months<br>- Subjects who CCTA without known or documented heart disease |
| **Exclusion criteria** | - History of myocardial infarction, percutaneous coronary intervention, coronary artery bypass grafting, cardiac transplantation, or valvular surgery<br>- Based on image review of chest X-ray by an expert: atelectasis, collapse, consolidation, pneumothorax, endotracheal tube, chest tube, and skeletal abnormality… |

**eTable 5. Summary of congenital heart disease, aortic aneurysm, and mediastinal mass groups**

| Cohorts name | **Congenital Heart Disease (CHD)**<br>**Aortic Aneurysm (Aneurysm)**<br>**Mediastinal Mass (Mass)** |
|---|---|
| **Institution** | Asan Medical Center, Seoul, Korea |
| **Providers** | Joom Bum Kim, Hyun Jung Koo (CHD)<br>Joom Bum Kim, Hyun Jung Koo, Joon Won Kang (Aneurysm)<br>Yura Ahn, Sangmin Lee (Mass) |
| **Date** | 2000 - 2017(CHD); 2000 - 2017(Aneurysm); 2017 - 2021 (Mass) |
| **Subject number** | N=1299 (CHD), N=294 (Aneurysm), N=110 (Mass) |
| **Data building and cleansing** | - Review of disease codes (International Classification of Disease, $10^{th}$ version, ICD-10)<br>- Review of structured report of transthoracic echocardiography or computed tomography<br>- Review of chest X-ray image and radiological report<br>- Image postprocessing of chest X-ray for excluding hyper-or hypo-inflated lung and anterior-to-posterior chest X-ray |
| **Inclusion criteria** | - Subjects aged ≥ 18<br>- Patients who underwent cardiac surgery for ventricular septal defect or atrial septal defect (CHD)<br>- Patients who diagnosed thoracic aortic aneurysm on CT (> 4.5cm in ascending aorta or > 4cm in arch or descending thoracic aorta) (Aneurysm)<br>- Patients who confirmed mediastinal mass in CT-guided biopsy (Mass) |
| **Exclusion criteria** | - Previous cardiothoracic operation<br>- Based on image review of chest X-ray by an expert: atelectasis, collapse, consolidation, pneumothorax, endotracheal tube, chest tube, and skeletal abnormality… |

**AI Model for analyzing cardiovascular borders on chest X-ray**

The definitions of cardiovascular borders (CVB) including superior vena cava/ascending aorta (SVC/AO), right atrium (RA), aortic arch or knob (Arch), pulmonary trunk (PT), left atrial appendage (LAA), left ventricle (LV), descending aorta (DAO), carina angle (Carina), and cardiothoracic (CT) ratio were provided in the eTable 6. Before CVB analysis, we applied a rigorous image selection protocol to ensure appropriate CXR (eFigure 3). Automatic exclusions were performed for CXRs obtained in anterior-to-posterior or lateral directions, as well as for images showing hyper- or hypo-inflation of the lungs using an in-house deep learning filter algorithm. The analysis software for CVB on X-ray comprises a deep learning model that automatically delineates cardiovascular borders and a software that calculates the width from the midline of the CXR to each individual CVB. The mask region-based convolutional neural network, which performs the drawing of each CVB, was trained using 1614 CXRs from Asan Medical Center, Seoul, Korea.[5] The model was tested on 1182 multi-institutional datasets of CXR that included both normal and valvular heart disease cases. The ground truth for CVB was generated by an expert radiologist (Dong Hyun Yang). The absolute percentage measurement error for CVB parameters ranged between 2.7% for the left ventricle (LV) and 11.1% for the carinal angle compared to the measurements by an expert radiologist.[6] The initial AI version was fine-tuned using 71413 Normal Korean CXRs. All CBs drawn by the AI was inspected by an experienced radiology technologist. 2077 (2.9%) CXRs of 71413 showed suboptimal results of CB drawing and manually corrected. The final deep learning model used in this study was retrained using all 71413 images. A trial of the web-based software is available on the ADC study website (www.adcstudy.com). (eFigure 4)

**eTable 6. Definitions of cardiovascular borders**

| **Name** | **Definitions** |
|---|---|
| Superior vena cava/ascending aorta **(SVC/AO)** | The right upper cardiovascular border (CVB) is delineated by the azygos vein and superior vena cava (SVC). The right upper CVB starts approximately 1cm above the right main bronchus because the azygos vein drains into the SVC just above the right main bronchus. Normally, the SVC forms this border, but if the ascending aorta enlarges, it may influence the appearance of the right upper CVB. |
| Right atrium **(RA)** | The right atrium creates the right lower CVB. The junction of the right upper and lower CVBs is defined as the point where the right lower bronchus overlaps the right-sided cardiac silhouette. |
| Aortic arch or knob **(Arch)** | The aortic knob refers to the appearance of the distal aortic arch as it curves posterolaterally to continue as the descending thoracic aorta. It appears as a laterally-projecting bulge and constitutes the upper edge of the left cardiomediastinal contour. |
| Pulmonary trunk **(PT)** | A bulge protrudes laterally beneath the aortic knob, suggesting the central and left pulmonary artery. As the left pulmonary artery passes over the left main bronchus, the shadow of the pulmonary artery can be seen above the left main bronchus. |
| Left atrial appendage **(LAA)** | Small, normally flat region between the pulmonary artery and the lower left CVB. When the left atrium is enlarged, this region bulges laterally. |
| Left ventricle **(LV)** | The lower edge of the left cardiomediastinal contour formed by the left ventricle. |
| Descending aorta **(DAO)** | Lateral contour of the descending thoracic aorta. It starts from inferior end of the aortic knob. |
| Carina angle **(Carina)** | The angle formed between the right and left main bronchi's lower borders. Due to upward compression by the left atrium, an expansion of the left atrium may be indicated by a widening of the carinal angle. |
| Cardiothoracic ratio **(CT ratio)** | The ratio of the maximal horizontal cardiac diameter to the maximal horizontal thoracic diameter (inner edge of ribs/edge of pleura). In this study, the CT ratio was calculated by dividing the maximum width of the right lower CVB (corresponding to the RA) and the left lower CVB (corresponding to the LV) by the maximal horizontal thoracic diameter. |

**eFigure 3. Chest X-ray analysis workflow**

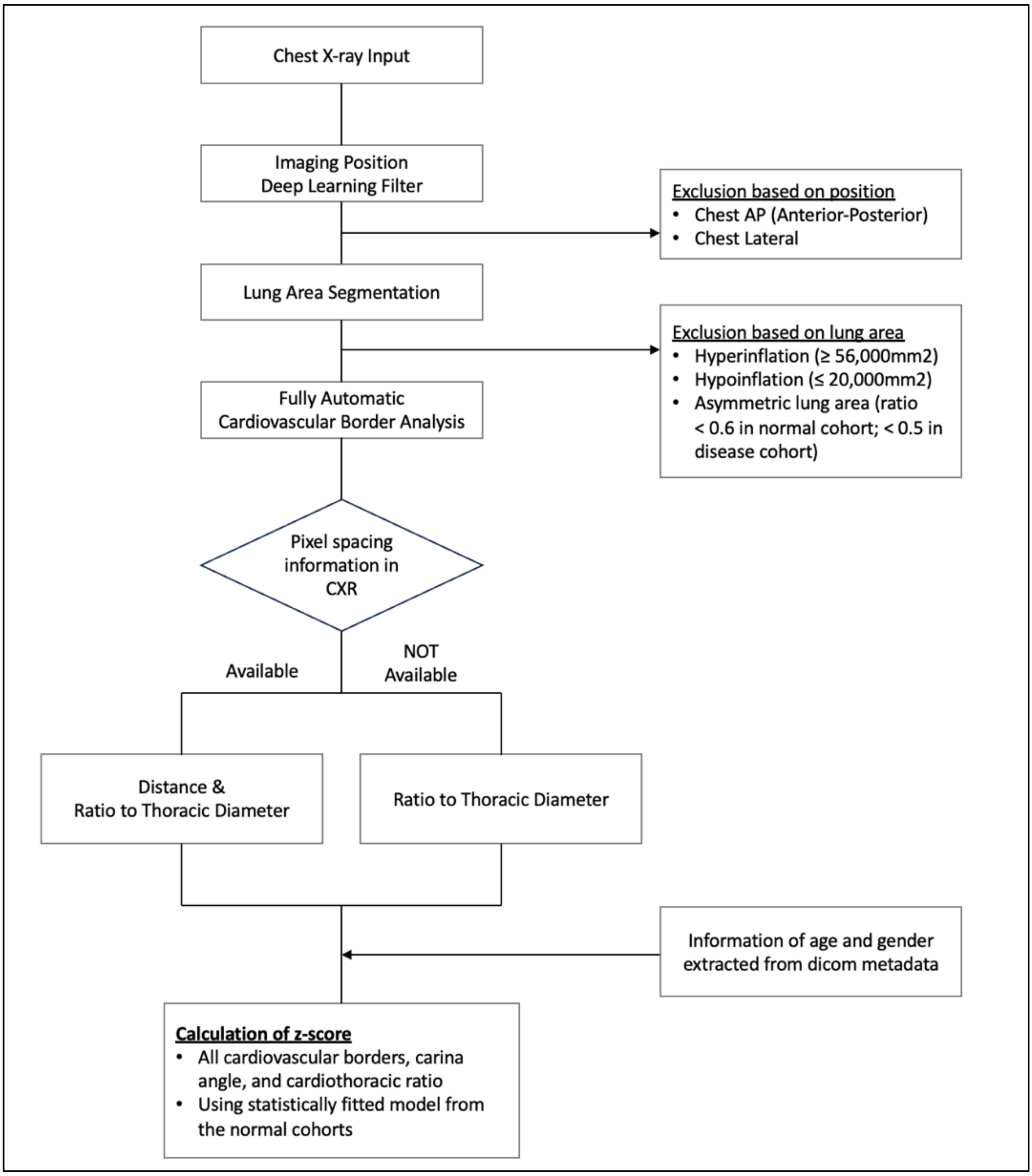

**eFigure 4. The ADC study website for the web-based trial of AI model**

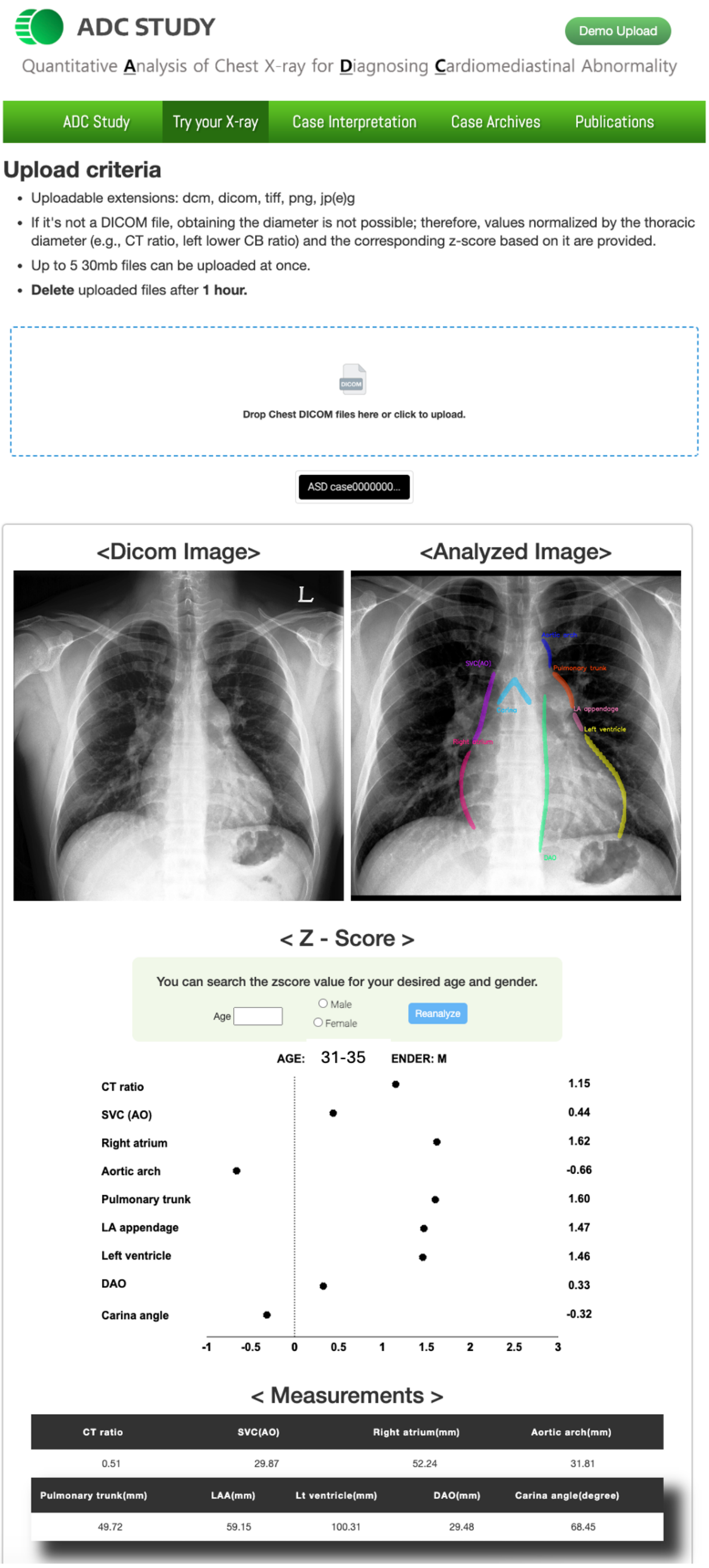

(www.adcstudy.com)

**eFigure 5. Histogram of cardiovascular borders in the Normal Korean**

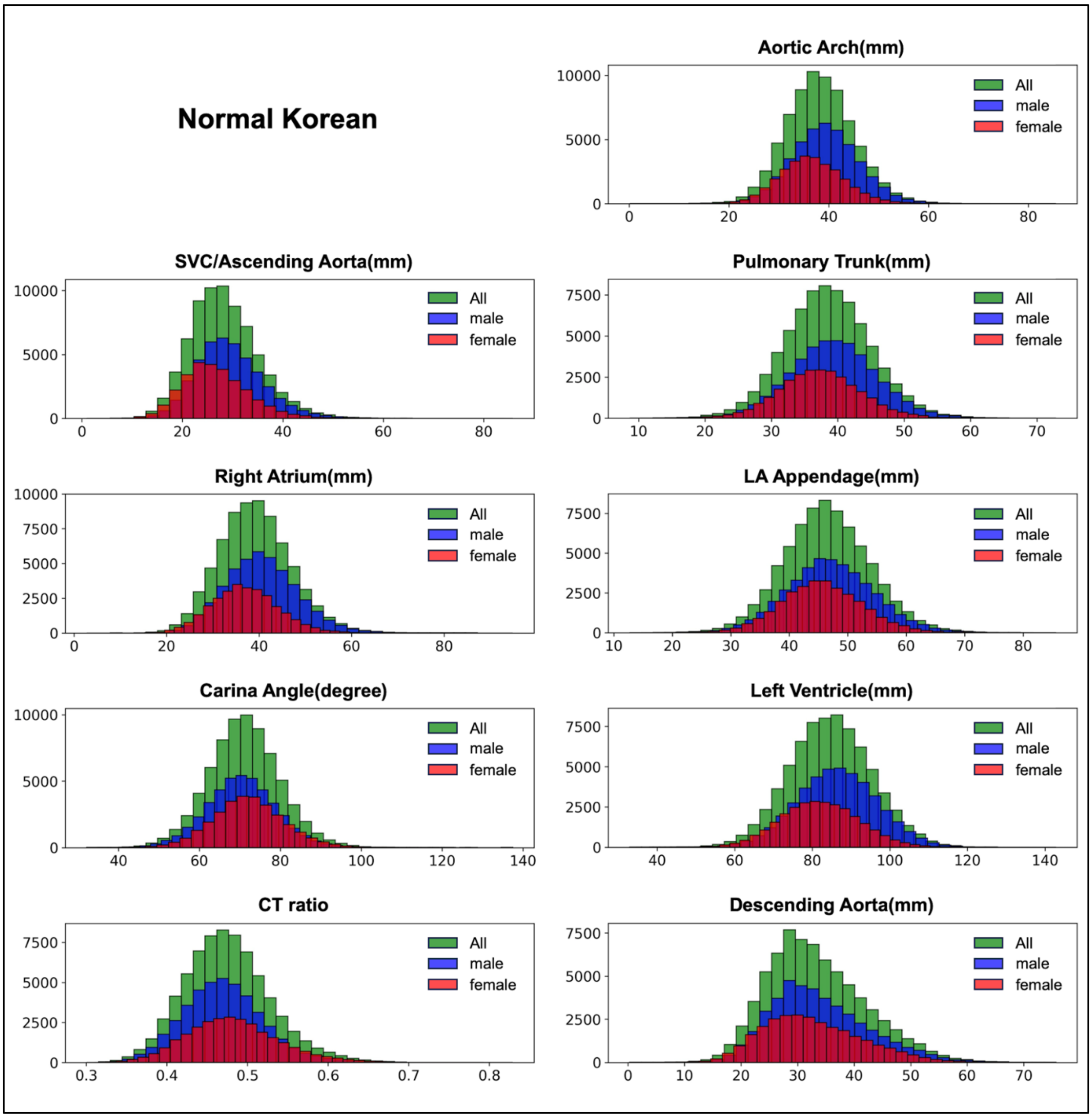

CT = cardiothoracic; LA = left atrium; SVC = superior vena cava

**eFigure 6. Histogram of cardiovascular borders in the Normal American**

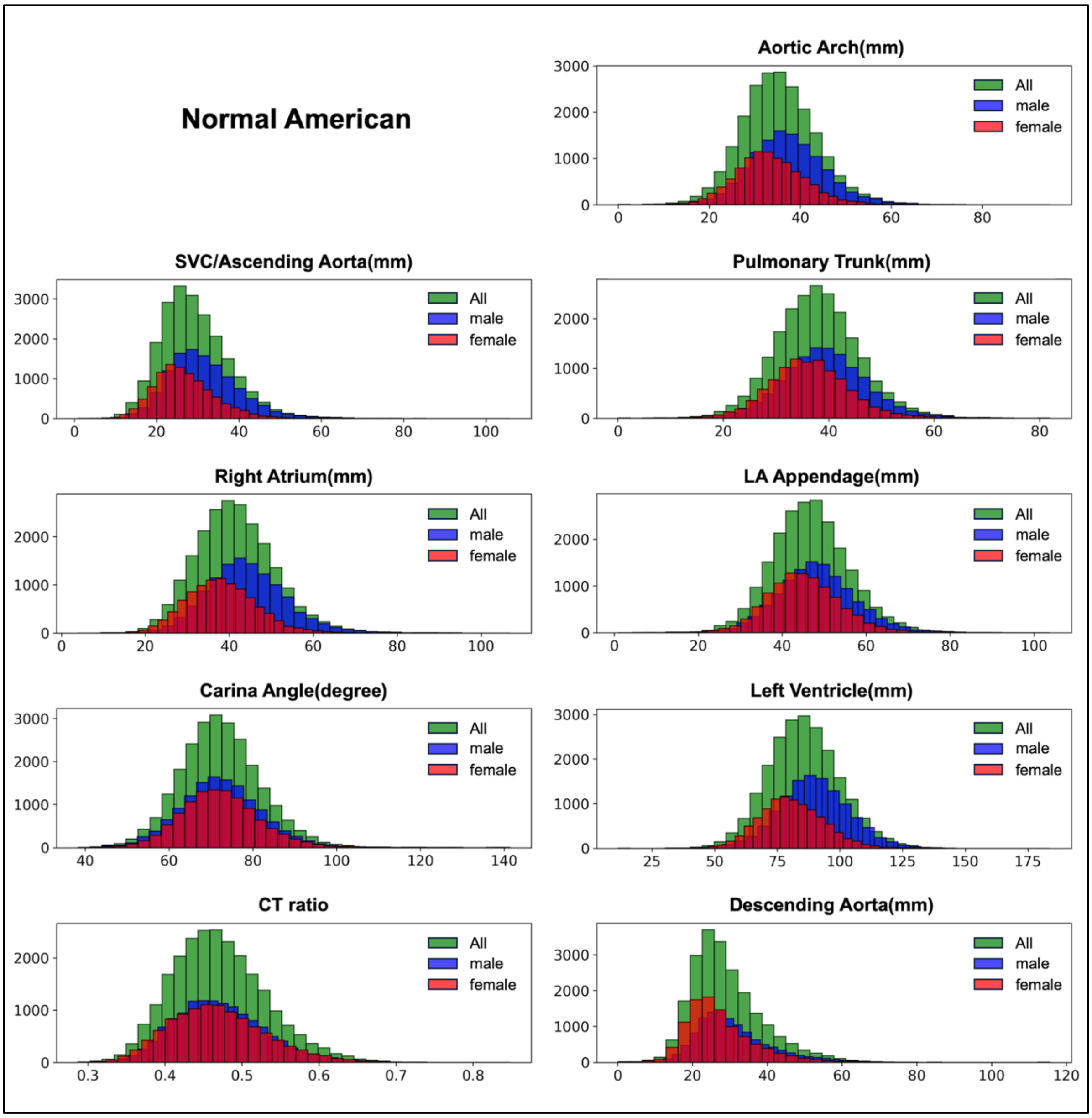

CT = cardiothoracic; LA = left atrium; SVC = superior vena cava

**eTable 7. Characteristics of valvular heart disease subgroups**

| | Aortic stenosis n=1432 | Aortic regurgitation n=1756 | Mitral stenosis n=2897 | Mitral regurgitation n=2971 | Tricuspid valve disease n=785 |
|---|---|---|---|---|---|
| Demographics | | | | | |
| Age, years | 67.9 (12.4) | 58.8 (15.3) | 54.1 (12.9) | 53.1 (15.9) | 58.0 (15.6) |
| Male gender, number (%) | 764 (53.4) | 979 (55.8) | 741 (25.6) | 1450 (48.8) | 307 (39.1) |
| Height, cm | 159.0 (9.0) | 162.0 (10.0) | 158.9 (8.0) | 162.2 (9.5) | 159.8 (9.6) |
| Weight, kg | 61.2 (10.3) | 61.4 (11.6) | 57.7 (9.7) | 61.2 (11.5) | 59.7 (11.2) |
| Body mass index, kg/$m^2$ | 24.1 (3.3) | 23.3 (3.3) | 22.9 (3.1) | 23.2 (3.5) | 23.3 (3.6) |
| Echocardiography | | | | | |
| LV EDV index, mL/$m^2$ | 98.3 (43.7) | 124.8 (60.6) | 83.4 (28.5) | 114.0 (49.5) | 79.5 (31.6) |
| LV ESV index, mL/$m^2$ | 41.9 (30.9) | 55.6 (37.7) | 35.2 (17.5) | 50.9 (35.2) | 32.2 (17.7) |
| LV ejection fraction, % | 59.6 (10.2) | 57.4 (10.0) | 60.9 (9.5) | 57.2 (11.3) | 60.4 (7.6) |
| LA diameter, mm | 41.0 (7.4) | 38.5 (6.7) | 48.8 (9.0) | 43.0 (9.2) | 42.3 (9.3) |
| LV mass index, g/$m^2$ | 120.1 (35.2) | 122.3 (45.3) | 86.7 (20.6) | 111.8 (34.7) | 91.5 (29.6) |
| Ascending aorta diameter, mm | 33.2 (11.5) | 35.6 (5.8) | 30.2 (4.0) | 32.5 (4.5) | 31.9 (4.1) |
| Combined functional MR, number (%) | 20 (1.4) | 13 (0.8) | 295 (8.4) | - | 11 (1.4) |
| Combined functional AR, number (%) | 62 (4.3) | - | 227 (7.8) | 16 (0.6) | 3 (0.4) |
| Combined significant TR, number (%) | 16 (1.1) | 8 (0.4) | 446 (45.4) | 60 (2.0) | - |
| Chest X-ray | | | | | |
| CT ratio | 0.55 (0.07) | 0.55 (0.07) | 0.57 (0.08) | 0.54 (0.08) | 0.59 (0.08) |
| SVC/AO, mm | 35.6 (9.3) | 35.1 (10.4) | 31.3 (8.4) | 30.7 (8.8) | 33.8 (10.7) |
| Right atrium, mm | 45.9 (10.3) | 45.6 (11.2) | 47.1 (10.8) | 44.7 (11.3) | 50.8 (13.7) |
| Aortic arch, mm | 40.6 (8.2) | 42.4 (8.7) | 37.4 (7.5) | 39.0 (7.8) | 39.7 (8.4) |
| Pulmonary trunk, mm | 40.2 (8.4) | 41.3 (8.4) | 45.4 (8.0) | 43.1 (8.3) | 46.4 (9.2) |
| Left atrial appendage, mm | 48.9 (9.7) | 50.4 (9.5) | 57.6 (9.1) | 53.8 (9.9) | 57.9 (10.9) |
| Left ventricle, mm | 95.3 (12.5) | 98.4 (14.4) | 96.4 (12.8) | 94.9 (14.9) | 101.2 (14.2) |
| Descending aorta, mm | 42.8 (10.5) | 44.1 (12.0) | 42.5 (10.0) | 40.7 (11.5) | 42.3 (12.5) |
| Carinal angle, degree | 77.7 (11.3) | 76.3 (10.8) | 81.8 (10.8) | 78.9 (11.1) | 81.7 (11.3) |

Values are mean (SD) or number (%). AR = aortic regurgitation; CT = cardiothoracic; EDV = end-diastolic volume; ESV = end-systolic volume; LV = left ventricle; MR = mitral regurgitation; SVC/AO = superior vena cava/ascending aorta; TR = tricuspid regurgitation

**eTable 8. Normal range of cardiovascular borders in chest X-ray**

| | Men | | | | Women | | | |
|---|---|---|---|---|---|---|---|---|
| Normal Korean | 20 years | 40 years | 60 years | 80 years | 20 years | 40 years | 60 years | 80 years |
| CT ratio | 0.41 (0.39–0.44) | 0.45 (0.42–0.48) | 0.47 (0.44–0.50) | 0.49 (0.46–0.53) | 0.41 (0.38–0.44) | 0.43 (0.40–0.46) | 0.49 (0.46–0.52) | 0.53 (0.50–0.57) |
| SVC/AO, mm | 25.6 (22.4–29.1) | 27.3 (23.7–31.2) | 29.8 (25.7–34.3) | 32.6 (27.6–38.0) | 22.1 (18.9–25.5) | 23.6 (20.4–27.1) | 27.1 (23.2–31.4) | 31.2 (25.8–37.2) |
| Right atrium, mm | 39.0 (34.5–43.7) | 39.7 (34.9–44.8) | 40.4 (35.4–45.7) | 41.1 (35.5–47.2) | 33.1 (28.9–37.6) | 34.8 (30.6–39.2) | 37.5 (32.8–42.3) | 38.1 (32.5–44.0) |
| Aortic arch, mm | 32.1 (28.7–35.5) | 37.0 (33.2–40.7) | 40.6 (36.3–44.9) | 42.0 (36.8–47.3) | 29.6 (26.2–33.1) | 33.1 (29.7–36.6) | 37.7 (33.8–41.7) | 39.7 (34.4–45.3) |
| Pulmonary trunk, mm | 38.1 (34.2–42.1) | 39.8 (35.7–44.0) | 38.9 (34.6–43.2) | 38.9 (33.7–44.0) | 36.3 (32.3–40.3) | 36.8 (33.1–40.5) | 37.3 (33.3–41.4) | 37.8 (32.5–43.3) |
| LAA, mm | 47.3 (42.6–52.0) | 48.3 (43.6–53.1) | 46.1 (41.2–51.2) | 45.1 (39.4–51.0) | 44.9 (40.3–49.6) | 45.2 (41.1–49.5) | 45.5 (40.9–50.1) | 45.3 (39.6–51.1) |
| Left ventricle, mm | 79.3 (72.2–86.4) | 85.9 (79.0–92.8) | 86.8 (80.0–93.6) | 87.0 (79.4–94.5) | 70.9 (64.5–77.5) | 76.4 (70.3–82.6) | 83.9 (77.7–90.2) | 86.7 (80.0–93.5) |
| DAO, mm | 24.2 (21.3–27.3) | 28.5 (25.0–32.2) | 35.9 (30.7–41.5) | 43.2 (36.8–50.0) | 21.8 (18.9–24.9) | 25.8 (22.3–29.3) | 34.5 (29.6–39.7) | 41.4 (34.7–48.5) |
| Carinal angle, degree | 72.1 (67.3–77.0) | 70.3 (65.2–75.4) | 70.0 (64.2–75.9) | 71.4 (64.7–78.2) | 72.8 (68.2–77.5) | 71.7 (66.8–76.8) | 71.8 (65.8–77.9) | 73.8 (66.9–80.9) |
| Normal American | 20 years | 40 years | 60 years | 80 years | 20 years | 40 years | 60 years | 80 years |
| CT ratio | 0.42 (0.39–0.46) | 0.46 (0.42–0.50) | 0.48 (0.44–0.51) | 0.49 (0.45–0.53) | 0.43 (0.39–0.47) | 0.45 (0.41–0.49) | 0.48 (0.45–0.53) | 0.50 (0.46–0.54) |
| SVC/AO, mm | 26.9 (23.0–31.3) | 29.5 (25.0–34.5) | 32.8 (27.6–38.6) | 36.2 (30.3–42.9) | 23.4 (19.9–27.4) | 25.3 (21.3–29.9) | 27.9 (23.2–33.3) | 30.1 (24.7–36.2) |
| Right atrium, mm | 41.6 (36.7–46.8) | 43.8 (38.3–49.7) | 43.9 (38.1–50.3) | 47.0 (40.3–54.3) | 36.5 (31.9–41.5) | 37.5 (32.4–43.0) | 38.5 (32.9–44.6) | 39.5 (33.4–46.2) |
| Aortic arch, mm | 31.5 (27.5–35.6) | 35.9 (31.7–40.3) | 39.9 (34.9–45.0) | 42.2 (35.8–48.9) | 29.0 (25.4–32.8) | 32.4 (28.3–36.7) | 35.8 (30.9–40.9) | 37.6 (31.8–43.7) |
| Pulmonary trunk, mm | 37.6 (33.1–42.2) | 39.1 (34.6–43.8) | 39.8 (34.6–45.2) | 39.9 (33.7–46.4) | 36.0 (32.0–40.1) | 36.1 (31.7–40.7) | 36.2 (31.3–41.5) | 36.3 (30.8–42.2) |
| LAA, mm | 46.6 (41.4–52.0) | 48.4 (43.0–54.0) | 48.5 (42.5–54.8) | 47.7 (40.8–54.9) | 45.0 (40.4–49.9) | 45.0 (39.9–50.4) | 44.6 (38.9–50.6) | 43.7 (37.5–50.3) |
| Left ventricle, mm | 82.2 (74.9–89.9) | 89.1 (81.0–97.4) | 93.0 (84.4–102.0) | 92.9 (83.8–102.3) | 75.4 (68.2–83.1) | 79.9 (72.2–88.1) | 83.2 (75.1–91.8) | 85.8 (77.3–94.8) |
| DAO, mm | 23.5 | 27.2 | 34.4 | 43.1 | 21.5 | 24.2 | 31.0 | 39.6 |

| | | | | | | | | |
|---|---|---|---|---|---|---|---|---|
| | (20.6–26.7) | (23.5–31.3) | (28.9–40.8) | (35.7–51.6) | (18.6–24.5) | (20.5–28.2) | (25.5–37.3) | (32.7–47.4) |
| Carinal angle, degree | 72.9 (67.3–78.7) | 71.6 (65.7–77.8) | 71.5 (65.2–78.1) | 73.2 (66.4–80.4) | 73.4 (68.2–79.0) | 71.2 (65.7–77.1) | 71.1 (65.1–77.6) | 72.7 (66.0–80.0) |

Values are presented in median (interquartile range). CT = cardiothoracic; DAO = descending aorta; LAA = left atrial appendage; SVC/AO = superior vena cava/ascending aorta.

**eFigure 7. Normal range of cardiovascular borders in the Normal Korean men**

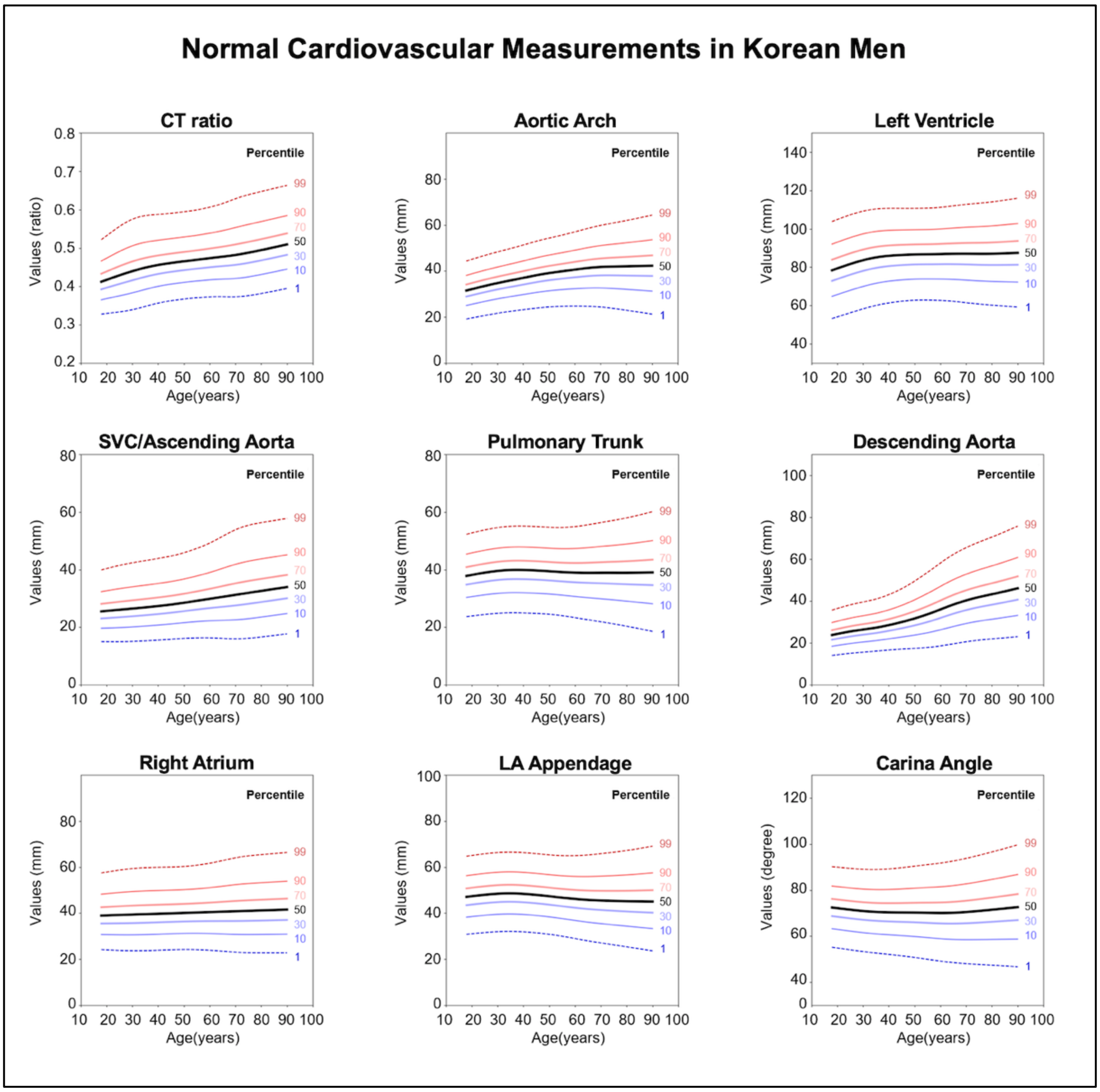

CT = cardiothoracic; LA = left atrial; SVC = superior vena cava

**eFigure 8. Normal range of cardiovascular borders in the Normal Korean women**

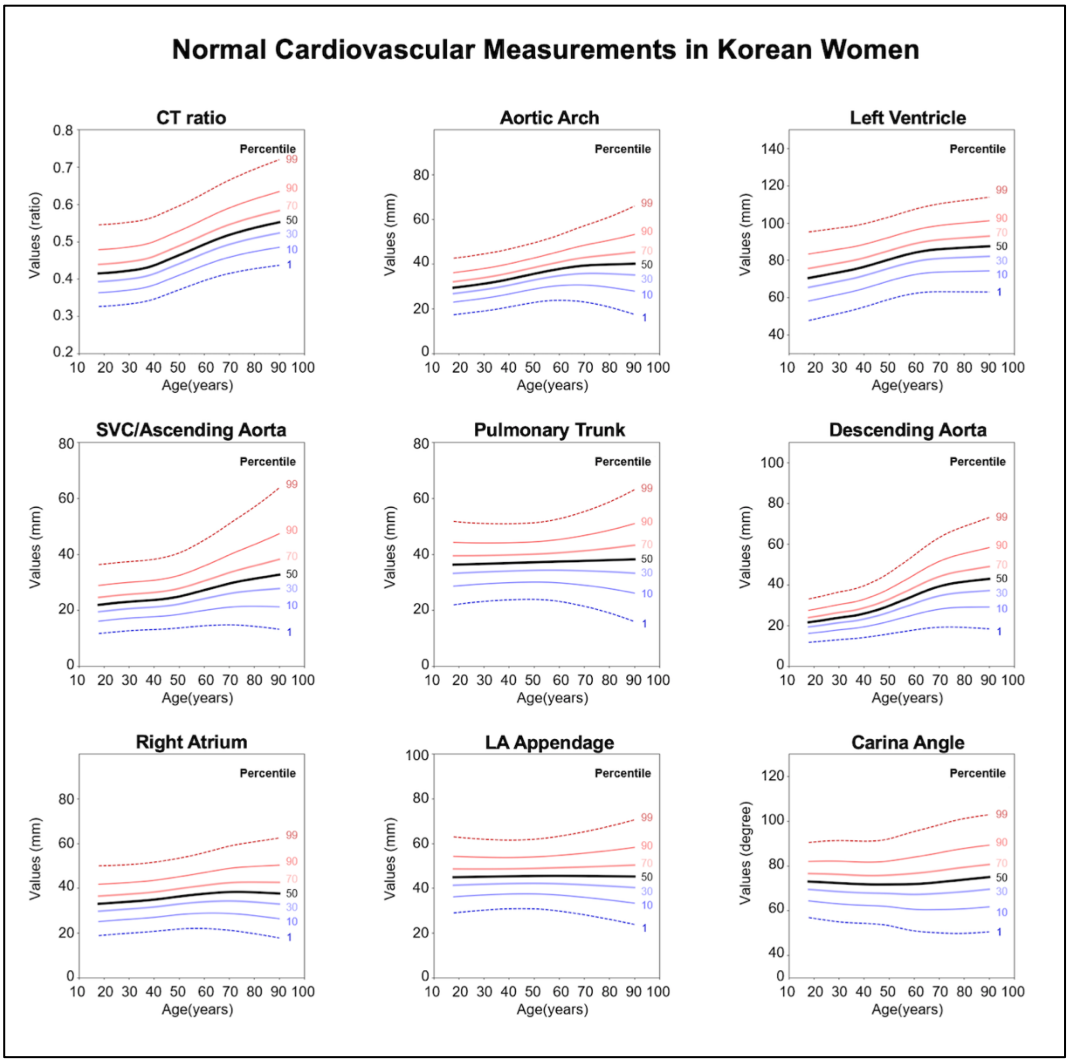

CT = cardiothoracic; LA = left atrial; SVC = superior vena cava

**eFigure 9. Normal range of cardiovascular borders in the Normal American men**

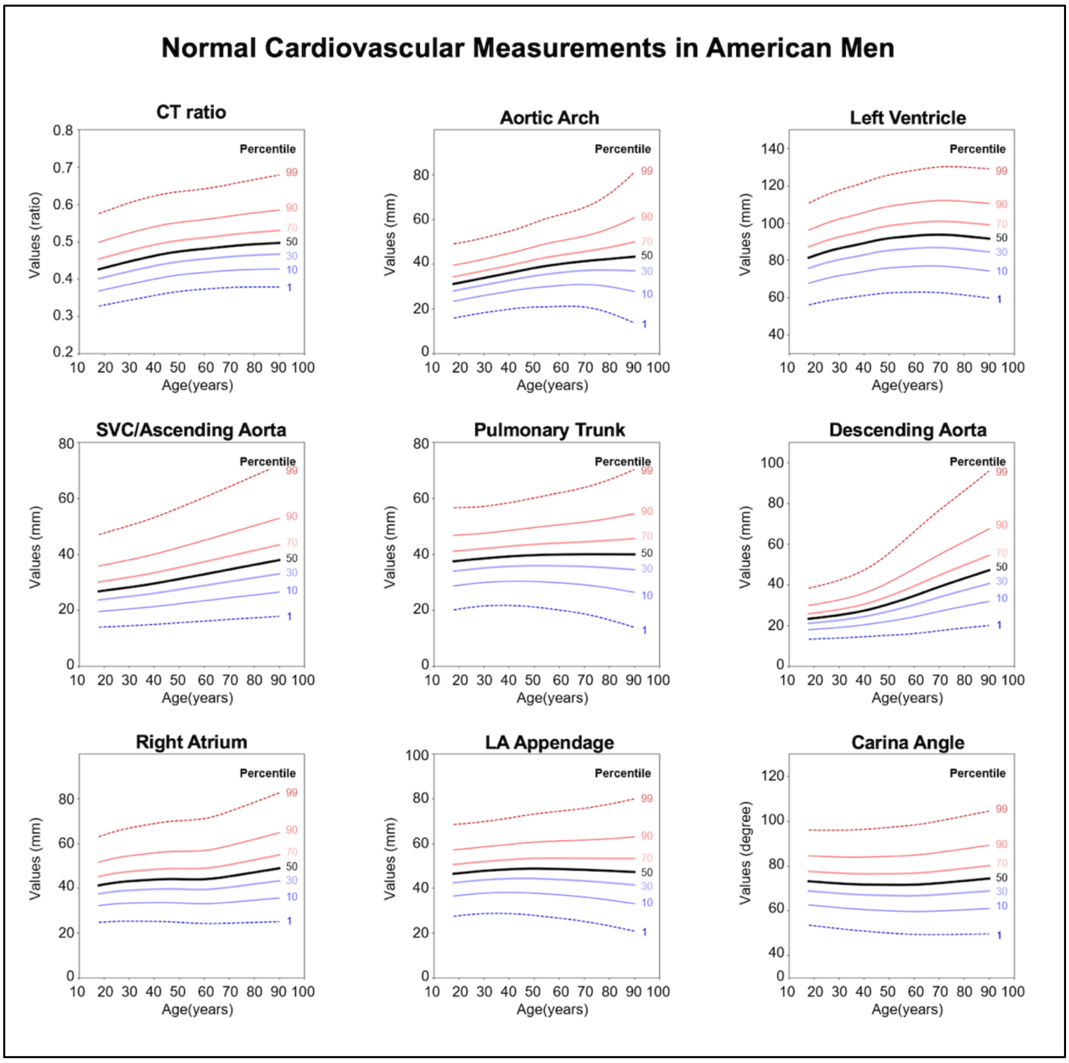

CT = cardiothoracic; LA = left atrial; SVC = superior vena cava

**eFigure 10. Normal range of cardiovascular borders in the Normal American women**

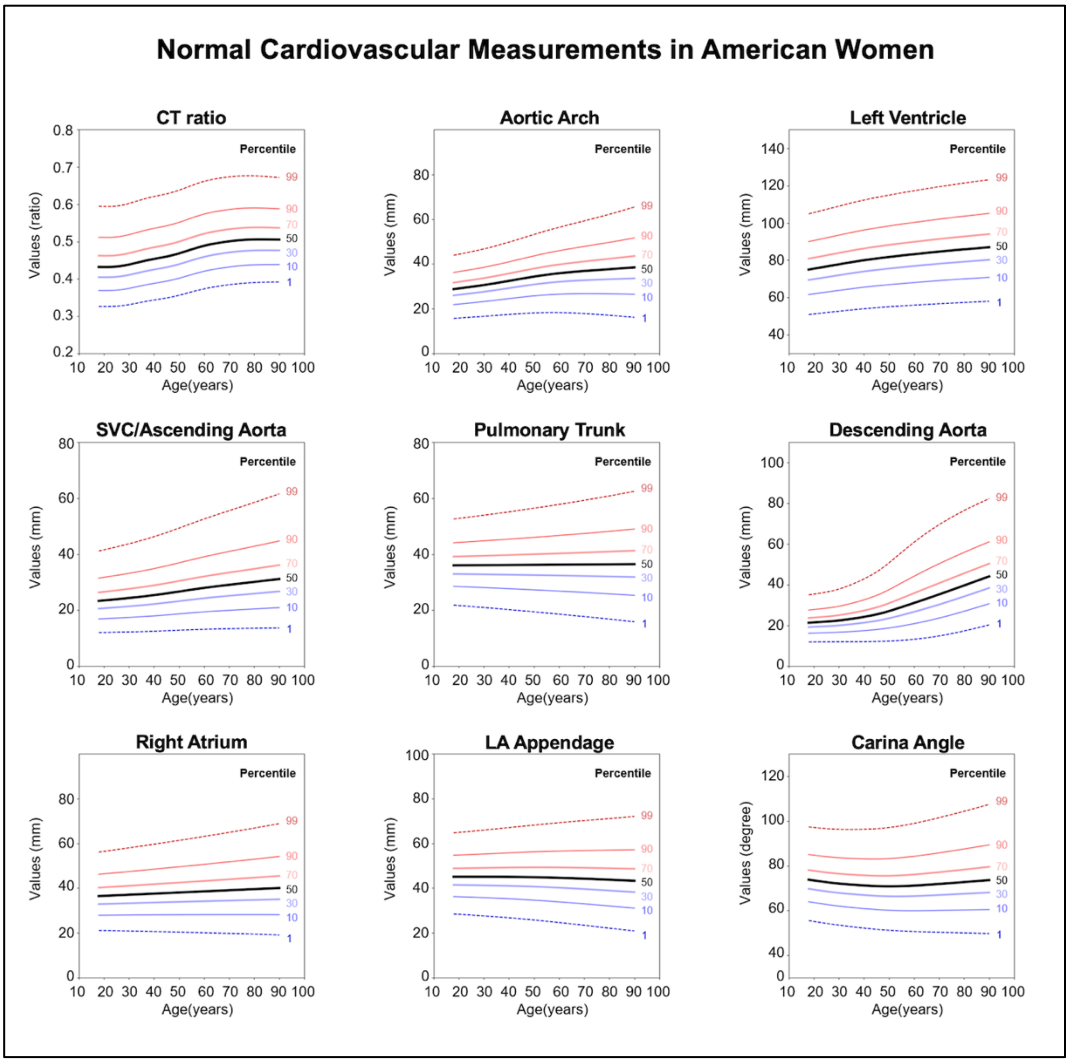

CT = cardiothoracic; LA = left atrial; SVC = superior vena cava

**eTable 9. Mean z-scores of cardiovascular borders in each disease group.**

| Disease group | Valvular heart disease | Coronary artery disease | Congenital heart disease | Aortic aneurysm | Mediastinal mass |
|---|---|---|---|---|---|
| CT ratio | 1.40 (1.38 - 1.43) | 0.39 (0.38 - 0.40) | 1.27 (1.20 - 1.34) | 1.50 (1.37 - 1.62) | 0.20 (0.00 - 0.41) |
| SVC/AO | 0.68 (0.65 - 0.70) | 0.24 (0.23 - 0.25) | 0.29 (0.23 - 0.36) | 1.18 (1.02 - 1.33) | 1.04 (0.77 - 1.32) |
| Right atrium | 0.95 (0.93 - 0.98) | 0.23 (0.22 - 0.24) | 0.70 (0.62 - 0.78) | 0.98 (0.82 - 1.13) | 0.28 (0.07 - 0.49) |
| Aortic arch | 0.34 (0.31 - 0.36) | 0.24 (0.23 - 0.25) | 0.01 (-0.06 - 0.07) | 1.95 (1.76 - 2.15) | 0.55 (0.30 - 0.80) |
| Pulmonary trunk | 0.78 (0.76 - 0.81) | 0.20 (0.19 - 0.22) | 1.34 (1.27 - 1.41) | 1.07 (0.90 - 1.24) | 1.03 (0.76 - 1.29) |
| Left atrial appendage | 1.01 (0.98 - 1.03) | 0.22 (0.21 - 0.23) | 1.53 (1.46 - 1.60) | 0.95 (0.79 - 1.10) | 0.90 (0.66 - 1.14) |
| Left ventricle | 1.26 (1.24 - 1.29) | 0.39 (0.38 - 0.41) | 1.48 (1.41 - 1.55) | 1.60 (1.46 - 1.75) | 0.25 (0.06 - 0.44) |
| Descending aorta | 1.37 (1.34 - 1.40) | 0.60 (0.59 - 0.62) | 0.40 (0.32 - 0.49) | 2.65 (2.47 - 2.83) | 0.42 (0.17 - 0.67) |
| Carinal angle | 0.85 (0.82 - 0.87) | 0.13 (0.11 - 0.14) | 0.66 (0.59 - 0.73) | 1.06 (0.91 - 1.21) | 0.71 (0.52 - 0.90) |
| Valvular heart disease subgroup | Aortic stenosis | Aortic regurgitation | Mitral stenosis | Mitral regurgitation | Tricuspid valve disease |
| CT ratio | 1.39 (1.34 - 1.45) | 1.39 (1.34 - 1.45) | 1.57 (1.53 - 1.62) | 1.13 (1.08 - 1.18) | 1.89 (1.81 - 1.97) |
| SVC (AO) | 1.02 (0.96 - 1.08) | 0.93 (0.87 - 0.99) | 0.63 (0.59 - 0.67) | 0.38 (0.34 - 0.42) | 0.83 (0.74 - 0.92) |
| Right atrium | 0.88 (0.82 - 0.94) | 0.81 (0.75 - 0.88) | 1.17 (1.13 - 1.22) | 0.74 (0.69 - 0.79) | 1.46 (1.36 - 1.56) |
| Aortic arch | 0.53 (0.46 - 0.59) | 0.73 (0.67 - 0.79) | 0.07 (0.03 - 0.11) | 0.25 (0.21 - 0.30) | 0.41 (0.32 - 0.50) |
| Pulmonary trunk | 0.27 (0.21 - 0.34) | 0.43 (0.38 - 0.49) | 1.17 (1.12 - 1.21) | 0.74 (0.69 - 0.78) | 1.22 (1.13 - 1.32) |
| Left atrial appendage | 0.33 (0.27 - 0.40) | 0.53 (0.47 - 0.59) | 1.54 (1.50 - 1.58) | 0.97 (0.93 - 1.02) | 1.48 (1.39 - 1.58) |
| Left ventricle | 1.13 (1.07 - 1.19) | 1.38 (1.32 - 1.44) | 1.34 (1.29 - 1.38) | 1.08 (1.03 - 1.13) | 1.71 (1.62 - 1.80) |
| Descending aorta | 1.45 (1.39 - 1.52) | 1.55 (1.48 - 1.61) | 1.50 (1.45 - 1.54) | 1.14 (1.09 - 1.19) | 1.36 (1.26 - 1.47) |
| Carinal angle | 0.67 (0.61 - 0.74) | 0.55 (0.49 - 0.60) | 1.10 (1.06 - 1.14) | 0.81 (0.77 - 0.86) | 1.09 (1.00 - 1.17) |

Values are presented in mean (95% confidence interval). CT = cardiothoracic; SVC/AO = superior vena cava/ascending aorta.

**eTable 10. Characteristics of coronary artery disease group and normal control**

| | Normal n=5937 | Coronary artery disease n=1979 | p-value |
|---|---|---|---|
| Demographics | | | |
| Age, years | 54.2 (11.3) | 63.3 (10.0) | <0.001 |
| Male gender, number (%) | 3599 (60.6) | 1424 (72.0) | <0.001 |
| Height, cm | 164.0 (8.8) | 163.5 (8.7) | 0.024 |
| Weight, kg | 63.9 (11.0) | 66.7 (10.7) | <0.001 |
| Body mass index, kg/$m^2$ | 23.6 (3.0) | 24.9 (3.0) | <0.001 |
| Echocardiography | | | |
| LV EDV index, mL/$m^2$ | 83.1 (20.4) | 91.4 (28.8) | <0.001 |
| LV ESV index, mL/$m^2$ | 30.7 (8.5) | 35.8 (18.0) | <0.001 |
| LV ejection fraction, % | 63.1 (3.7) | 61.7 (6.4) | <0.001 |
| LA diameter, mm | 33.6 (3.5) | 37.8 (5.1) | <0.001 |
| LV mass index, g/$m^2$ | 79.8 (15.9) | 92.3 (22.4) | <0.001 |
| Ascending aorta diameter, mm | 32.1 (3.4) | 34.0 (3.8) | <0.001 |
| Chest X-ray | | | |
| CT ratio | 0.48 (0.05) | 0.51 (0.06) | <0.001 |
| SVC/AO, mm | 28.4 (6.8) | 31.7 (7.7) | <0.001 |
| Right atrium, mm | 39.2 (7.8) | 42.1 (8.5) | <0.001 |
| Aortic arch, mm | 38.3 (6.7) | 40.5 (7.2) | <0.001 |
| Pulmonary trunk, mm | 38.5 (6.3) | 40.1 (7.3) | <0.001 |
| Left atrial appendage, mm | 46.4 (7.3) | 48.4 (8.2) | <0.001 |
| Left ventricle, mm | 84.6 (10.4) | 91.3 (11.2) | <0.001 |
| Descending aorta, mm | 33.4 (8.6) | 39.4 (10.1) | <0.001 |
| Carinal angle, degree | 70.9 (8.6) | 71.8 (10.2) | <0.001 |

Values are mean (SD) or number (%). CT = cardiothoracic; EDV = end-diastolic volume; ESV = end-systolic volume; LA = left atrium; LV = left ventricle; SVC/AO = superior vena cava/ascending aorta

**eTable 11. Characteristics of valvular heart disease group and normal control**

| | Normal n=11696 | Valvular heart disease n=3898 | p-value |
|---|---|---|---|
| Demographics | | | |
| Age, years | 54.4 (11.4) | 57.0 (15.3) | <0.001 |
| Male gender, number (%) | 7051 (60.3) | 1697 (43.5) | <0.001 |
| Height, cm | 164.2 (8.7) | 160.4 (9.2) | <0.001 |
| Weight, kg | 63.8 (10.6) | 59.9 (10.8) | <0.001 |
| Body mass index, $kg/m^2$ | 23.6 (2.9) | 23.3 (3.4) | <0.001 |
| Echocardiography | | | |
| LV EDV index, $mL/m^2$ | 83.6 (21.0) | 108.1 (50.9) | <0.001 |
| LV ESV index, $mL/m^2$ | 30.8 (8.6) | 47.2 (32.2) | <0.001 |
| LV ejection fraction, % | 63.1 (3.6) | 59.2 (10.2) | <0.001 |
| LA diameter, mm | 33.6 (3.5) | 42.2 (8.8) | <0.001 |
| LV mass index, $g/m^2$ | 79.8 (16.0) | 112.7 (37.3) | <0.001 |
| Ascending aorta diameter, mm | 32.1 (3.5) | 33.2 (9.3) | <0.001 |
| Chest X-ray | | | |
| CT ratio | 0.48 (0.05) | 0.55 (0.08) | <0.001 |
| SVC/AO, mm | 28.4 (6.7) | 32.6 (9.3) | <0.001 |
| Right atrium, mm | 39.3 (7.8) | 46.2 (11.1) | <0.001 |
| Aortic arch, mm | 38.3 (6.7) | 39.3 (8.0) | <0.001 |
| Pulmonary trunk, mm | 38.3 (6.4) | 43.2 (8.3) | <0.001 |
| Left atrial appendage, mm | 46.2 (7.3) | 53.8 (9.9) | <0.001 |
| Left ventricle, mm | 84.5 (10.4) | 96.5 (13.6) | <0.001 |
| Descending aorta, mm | 33.3 (8.6) | 42.1 (11.1) | <0.001 |
| Carinal angle, degree | 71.0 (8.7) | 79.0 (11.2) | <0.001 |

Values are mean (SD) or number (%). CT = cardiothoracic; EDV = end-diastolic volume; ESV = end-systolic volume; LA = left atrium; LV = left ventricle; SVC/AO = superior vena cava/ascending aorta

**eTable 12. Characteristics of congenital heart disease group and normal control**

| | Normal<br>n=1454 | Congenital heart disease<br>n=484 | p-value |
|---|---|---|---|
| Demographics | | | |
| Age, years | 54.3 (11.3) | 46.9 (14.5) | <0.001 |
| Male gender, number (%) | 830 (57.1) | 198 (40.9) | <0.001 |
| Height, cm | 163.8 (8.6) | 162.2 (9.1) | <0.001 |
| Weight, kg | 63.3 (10.5) | 59.9 (10.8) | <0.001 |
| Body mass index, kg/m$^2$ | 23.5 (2.9) | 22.6 (3.2) | <0.001 |
| Echocardiography | | | |
| LV EDV index, mL/m$^2$ | 82.3 (20.4) | 84.4 (30.7) | 0.25 |
| LV ESV index, mL/m$^2$ | 30.3 (8.3) | 32.7 (14.6) | 0.001 |
| LV ejection fraction, % | 63.1 (3.5) | 61.7 (6.2) | <0.001 |
| LA diameter, mm | 33.4 (3.5) | 39.3 (8.4) | <0.001 |
| LV mass index, g/m$^2$ | 80.2 (15.2) | 81.6 (22.6) | 0.311 |
| Ascending aorta diameter, mm | 32.1 (3.4) | 31.5 (4.6) | 0.048 |
| Chest X-ray | | | |
| CT ratio | 0.48 (0.05) | 0.55 (0.08) | <0.001 |
| SVC/AO, mm | 28.4 (6.9) | 29.7 (8.6) | 0.001 |
| Right atrium, mm | 39.1 (7.8) | 44.8 (12.1) | <0.001 |
| Aortic arch, mm | 38.5 (6.6) | 36.7 (7.6) | <0.001 |
| Pulmonary trunk, mm | 38.7 (6.6) | 46.2 (8.9) | <0.001 |
| Left atrial appendage, mm | 46.7 (7.5) | 57.3 (10.3) | <0.001 |
| Left ventricle, mm | 84.4 (10.7) | 98.4 (14.9) | <0.001 |
| Descending aorta, mm | 33.4 (8.4) | 34.5 (11.2) | 0.022 |
| Carinal angle, degree | 70.8 (8.8) | 77.2 (10.9) | <0.001 |

Values are mean (SD) or number (%). CT = cardiothoracic; EDV = end-diastolic volume; ESV = end-systolic volume; LA = left atrium; LV = left ventricle; SVC/AO = superior vena cava/ascending aorta

**eTable 13. Characteristics of aortic valve disease group and normal control**

| | Normal<br>n=3786 | Aortic valve disease<br>n=1262 | p-value |
|---|---|---|---|
| Demographics | | | |
| Age, years | 53.9 (11.4) | 62.7 (14.6) | <0.001 |
| Male gender, number (%) | 2251 (59.5) | 694 (55.0) | 0.006 |
| Height, cm | 164.1 (8.7) | 160.8 (9.7) | <0.001 |
| Weight, kg | 63.6 (10.7) | 61.6 (10.9) | <0.001 |
| Body mass index, kg/$m^2$ | 23.5 (2.9) | 23.8 (3.3) | 0.008 |
| Echocardiography | | | |
| LV EDV index, mL/$m^2$ | 83.1 (20.3) | 109.8 (52.9) | <0.001 |
| LV ESV index, mL/$m^2$ | 30.7 (8.4) | 48.0 (33.9) | <0.001 |
| LV ejection fraction, % | 63.1 (3.6) | 58.4 (10.3) | <0.001 |
| LA diameter, mm | 33.5 (3.6) | 39.8 (7.5) | <0.001 |
| LV mass index, g/$m^2$ | 79.6 (16.0) | 118.2 (39.2) | <0.001 |
| Ascending aorta diameter, mm | 32.0 (3.4) | 33.9 (4.9) | <0.001 |
| Chest X-ray | | | |
| CT ratio | 0.47 (0.05) | 0.55 (0.07) | <0.001 |
| SVC/AO, mm | 28.2 (6.8) | 35.0 (9.5) | <0.001 |
| Right atrium, mm | 39.1 (7.6) | 45.7 (10.4) | <0.001 |
| Aortic arch, mm | 38.1 (6.5) | 41.7 (8.5) | <0.001 |
| Pulmonary trunk, mm | 38.3 (6.3) | 40.8 (8.4) | <0.001 |
| Left atrial appendage, mm | 46.2 (7.1) | 49.9 (9.8) | <0.001 |
| Left ventricle, mm | 84.4 (10.2) | 97.2 (13.5) | <0.001 |
| Descending aorta, mm | 33.3 (8.5) | 43.2 (11.2) | <0.001 |
| Carinal angle, degree | 70.8 (8.7) | 76.8 (11.0) | <0.001 |

Values are mean (SD) or number (%). CT = cardiothoracic; EDV = end-diastolic volume; ESV = end-systolic volume; LA = left atrium; LV = left ventricle; SVC/AO = superior vena cava/ascending aorta

**eTable 14. Characteristics of mitral valve disease group and normal control**

| | Normal n=6921 | Mitral valve disease n=2307 | p-value |
|---|---|---|---|
| Demographics | | | |
| Age, years | 54.3 (11.5) | 53.1 (14.3) | <0.001 |
| Male gender, number (%) | 4150 (60.0) | 861 (37.3) | <0.001 |
| Height, cm | 164.0 (8.7) | 160.6 (8.8) | <0.001 |
| Weight, kg | 63.7 (10.8) | 59.5 (10.6) | <0.001 |
| Body mass index, $kg/m^2$ | 23.5 (2.9) | 23.1 (3.3) | <0.001 |
| Echocardiography | | | |
| LV EDV index, $mL/m^2$ | 83.1 (20.8) | 104.5 (43.7) | <0.001 |
| LV ESV index, $mL/m^2$ | 30.6 (8.6) | 45.3 (28.0) | <0.001 |
| LV ejection fraction, % | 63.2 (3.7) | 59.8 (9.9) | <0.001 |
| LA diameter, mm | 33.6 (3.6) | 44.7 (9.5) | <0.001 |
| LV mass index, $g/m^2$ | 80.0 (16.1) | 102.7 (31.8) | <0.001 |
| Ascending aorta diameter, mm | 32.1 (3.4) | 31.9 (4.4) | 0.264 |
| Chest X-ray | | | |
| CT ratio | 0.48 (0.05) | 0.55 (0.08) | <0.001 |
| SVC/AO, mm | 28.3 (6.7) | 30.7 (8.2) | <0.001 |
| Right atrium, mm | 39.2 (7.7) | 45.5 (10.8) | <0.001 |
| Aortic arch, mm | 38.3 (6.6) | 37.8 (7.6) | 0.011 |
| Pulmonary trunk, mm | 38.5 (6.4) | 44.0 (8.1) | <0.001 |
| Left atrial appendage, mm | 46.5 (7.3) | 55.6 (9.6) | <0.001 |
| Left ventricle, mm | 84.7 (10.6) | 95.6 (13.7) | <0.001 |
| Descending aorta, mm | 33.4 (8.5) | 41.1 (10.5) | <0.001 |
| Carinal angle, degree | 71.3 (8.6) | 80.1 (10.9) | <0.001 |

Values are mean (SD) or number (%). CT = cardiothoracic; EDV = end-diastolic volume; ESV = end-systolic volume; LA = left atrium; LV = left ventricle; SVC/AO = superior vena cava/ascending aorta

**eTable 15. Characteristics of tricuspid valve disease group and normal control**

| | Normal n=900 | Tricuspid valve disease n=300 | p-value |
|---|---|---|---|
| Demographics | | | |
| Age, years | 55.1 (10.9) | 58.1 (15.2) | <0.001 |
| Male gender, number (%) | 545 (60.6) | 136 (45.3) | <0.001 |
| Height, cm | 164.5 (8.9) | 161.0 (9.4) | <0.001 |
| Weight, kg | 64.4 (10.9) | 60.9 (11.3) | <0.001 |
| Body mass index, kg/m$^2$ | 23.7 (2.8) | 23.4 (3.6) | 0.309 |
| Echocardiography | | | |
| LV EDV index, mL/m$^2$ | 82.8 (21.6) | 79.5 (35.3) | 0.201 |
| LV ESV index, mL/m$^2$ | 30.5 (8.9) | 31.3 (19.1) | 0.453 |
| LV ejection fraction, % | 63.2 (3.7) | 61.4 (7.1) | <0.001 |
| LA diameter, mm | 33.5 (3.5) | 42.6 (9.9) | <0.001 |
| LV mass index, g/m$^2$ | 78.1 (16.8) | 91.3 (32.6) | <0.001 |
| Ascending aorta diameter, mm | 32.0 (3.5) | 32.3 (4.5) | 0.497 |
| Chest X-ray | | | |
| CT ratio | 0.48 (0.05) | 0.59 (0.08) | <0.001 |
| SVC/AO, mm | 28.6 (6.9) | 33.8 (9.6) | <0.001 |
| Right atrium, mm | 39.3 (7.6) | 51.5 (13.3) | <0.001 |
| Aortic arch, mm | 38.3 (6.8) | 39.9 (8.3) | 0.001 |
| Pulmonary trunk, mm | 38.3 (6.5) | 46.3 (9.5) | <0.001 |
| Left atrial appendage, mm | 46.2 (7.4) | 57.2 (10.8) | <0.001 |
| Left ventricle, mm | 84.6 (10.4) | 101.3 (13.9) | <0.001 |
| Descending aorta, mm | 33.7 (8.7) | 42.4 (11.7) | <0.001 |
| Carinal angle, degree | 70.9 (8.8) | 81.1 (11.6) | <0.001 |

Values are mean (SD) or number (%). CT = cardiothoracic; EDV = end-diastolic volume; ESV = end-systolic volume; LA = left atrium; LV = left ventricle; SVC/AO = superior vena cava/ascending aorta

**eTable 16. Diagnostic performance in coronary artery disease, valvular heart disease, and congenital heart disease**

| Coronary artery disease<br>disease n=1979(25%)<br>control n=5937 (75%) | AUC<br>(95% CI) | Cut-off<br>z-score | Sensitivity | Specificity | Accuracy | Positive<br>predictive value | Negative<br>predictive value |
|---|---|---|---|---|---|---|---|
| CT ratio | 0.67 (0.66 - 0.69) | 0.54 | 0.57 (0.55 - 0.60) | 0.68 (0.67 - 0.69) | 0.65 (0.64 - 0.66) | 0.38 (0.36 - 0.39) | 0.83 (0.81 - 0.84) |
| SVC/AO | 0.60 (0.59 - 0.62) | 0.29 | 0.57 (0.55 - 0.59) | 0.60 (0.59 - 0.61) | 0.59 (0.58 - 0.60) | 0.32 (0.31 - 0.34) | 0.81 (0.79 - 0.82) |
| Right atrium | 0.58 (0.56 - 0.59) | 0.59 | 0.41 (0.39 - 0.43) | 0.71 (0.70 - 0.72) | 0.63 (0.62 - 0.65) | 0.32 (0.30 - 0.34) | 0.78 (0.77 - 0.79) |
| Aortic arch | 0.58 (0.56 - 0.59) | 0.22 | 0.56 (0.54 - 0.58) | 0.57 (0.56 - 0.58) | 0.57 (0.56 - 0.58) | 0.30 (0.29 - 0.32) | 0.79 (0.78 - 0.80) |
| PT | 0.55 (0.54 - 0.57) | 0.62 | 0.36 (0.34 - 0.38) | 0.73 (0.72 - 0.74) | 0.64 (0.63 - 0.65) | 0.31 (0.29 - 0.33) | 0.77 (0.76 - 0.78) |
| Left atrial appendage | 0.56 (0.55 - 0.58) | 0.64 | 0.36 (0.34 - 0.38) | 0.74 (0.73 - 0.75) | 0.64 (0.63 - 0.65) | 0.32 (0.30 - 0.34) | 0.77 (0.76 - 0.78) |
| LV | 0.65 (0.64 - 0.67) | 0.47 | 0.55 (0.52 - 0.57) | 0.67 (0.66 - 0.68) | 0.64 (0.63 - 0.65) | 0.36 (0.34 - 0.37) | 0.81 (0.80 - 0.82) |
| Carina | 0.52 (0.50 - 0.53) | 0.98 | 0.22 (0.20 - 0.24) | 0.84 (0.83 - 0.85) | 0.68 (0.67 - 0.69) | 0.32 (0.29 - 0.34) | 0.76 (0.75 - 0.77) |
| Descending aorta | 0.66 (0.64 - 0.67) | 0.82 | 0.55 (0.53 - 0.57) | 0.68 (0.67 - 0.70) | 0.65 (0.64 - 0.66) | 0.37 (0.35 - 0.38) | 0.82 (0.81 - 0.83) |
| Logistic Model | | | | | | | |
| LV + SVC/AO | 0.69 (0.68 - 0.70) | - | 0.70 (0.68 - 0.72) | 0.57 (0.56 - 0.59) | 0.61 (0.59 - 0.62) | 0.35 (0.34 - 0.37) | 0.85 (0.84 - 0.86) |
| LV + SVC/AO + Carina | 0.69 (0.68 - 0.70) | - | 0.64 (0.62 - 0.66) | 0.63 (0.62 - 0.65) | 0.64 (0.62 - 0.65) | 0.37 (0.35 - 0.38) | 0.84 (0.83 - 0.85) |
| Valvular heart disease<br>disease n=3898 (25%)<br>control n=11696 (75%) | AUC<br>(95% CI) | Cut-off<br>z-score | Sensitivity | Specificity | Accuracy | Positive<br>predictive value | Negative<br>predictive value |
| CT ratio | 0.79 (0.78 - 0.80) | 0.94 | 0.65 (0.64 - 0.67) | 0.80 (0.79 - 0.81) | 0.76 (0.76 - 0.77) | 0.52 (0.51 - 0.54) | 0.87 (0.86 - 0.88) |
| SVC/AO | 0.64 (0.63 - 0.65) | 0.74 | 0.48 (0.46 - 0.50) | 0.74 (0.74 - 0.75) | 0.68 (0.67 - 0.69) | 0.39 (0.37 - 0.40) | 0.81 (0.80 - 0.82) |
| Right atrium | 0.71 (0.70 - 0.72) | 0.82 | 0.53 (0.52 - 0.55) | 0.78 (0.78 - 0.79) | 0.72 (0.71 - 0.73) | 0.45 (0.44 - 0.47) | 0.83 (0.83 - 0.84) |
| Aortic arch | 0.56 (0.55 - 0.57) | 0.35 | 0.48 (0.46 - 0.49) | 0.61 (0.60 - 0.62) | 0.58 (0.57 - 0.59) | 0.29 (0.28 - 0.30) | 0.78 (0.77 - 0.78) |
| PT | 0.69 (0.68 - 0.70) | 0.73 | 0.51 (0.49 - 0.52) | 0.77 (0.76 - 0.78) | 0.70 (0.70 - 0.71) | 0.43 (0.41 - 0.44) | 0.82 (0.81 - 0.83) |
| Left atrial appendage | 0.73 (0.72 - 0.74) | 0.62 | 0.62 (0.61 - 0.64) | 0.73 (0.73 - 0.74) | 0.71 (0.70 - 0.71) | 0.44 (0.43 - 0.45) | 0.85 (0.84 - 0.86) |
| LV | 0.78 (0.77 - 0.79) | 0.72 | 0.67 (0.66 - 0.69) | 0.75 (0.74 - 0.76) | 0.73 (0.72 - 0.74) | 0.47 (0.46 - 0.49) | 0.87 (0.86 - 0.88) |
| Carina | 0.69 (0.68 - 0.70) | 0.54 | 0.58 (0.56 - 0.60) | 0.71 (0.70 - 0.71) | 0.67 (0.67 - 0.68) | 0.40 (0.39 - 0.41) | 0.83 (0.82 - 0.84) |
| Descending aorta | 0.73 (0.72 - 0.74) | 1.09 | 0.60 (0.59 - 0.62) | 0.75 (0.74 - 0.76) | 0.71 (0.71 - 0.72) | 0.45 (0.44 - 0.46) | 0.85 (0.84 - 0.85) |
| Logistic Model | | | | | | | |
| LV + Right atrium | 0.82 (0.82 - 0.83) | - | 0.70 (0.69 - 0.72) | 0.81 (0.80 - 0.81) | 0.78 (0.78 - 0.79) | 0.55 (0.54 - 0.57) | 0.89 (0.88 - 0.89) |
| LV + SVC/AO | 0.81 (0.80 - 0.81) | - | 0.70 (0.69 - 0.72) | 0.77 (0.76 - 0.78) | 0.75 (0.75 - 0.76) | 0.51 (0.49 - 0.52) | 0.88 (0.88 - 0.89) |
| Congenital heart disease<br>disease n=484 (25%)<br>control n=1454 (75%) | AUC<br>(95% CI) | Cut-off<br>z-score | Sensitivity | Specificity | Accuracy | Positive<br>predictive value | Negative<br>predictive value |

| | | | | | | | |
|---|---|---|---|---|---|---|---|
| CT ratio | 0.77 (0.74 - 0.79) | 0.82 | 0.64 (0.60 - 0.68) | 0.79 (0.76 - 0.81) | 0.75 (0.73 - 0.77) | 0.50 (0.46 - 0.54) | 0.87 (0.85 - 0.88) |
| SVC/AO | 0.54 (0.51 - 0.57) | 0.97 | 0.29 (0.26 - 0.34) | 0.79 (0.77 - 0.81) | 0.66 (0.64 - 0.68) | 0.32 (0.28 - 0.36) | 0.77 (0.75 - 0.79) |
| Right atrium | 0.66 (0.63 - 0.69) | 0.59 | 0.54 (0.49 - 0.58) | 0.71 (0.68 - 0.73) | 0.67 (0.64 - 0.69) | 0.38 (0.35 - 0.42) | 0.82 (0.80 - 0.84) |
| Aortic arch | 0.55 (0.52 - 0.58) | -0.01 | 0.44 (0.39 - 0.48) | 0.46 (0.43 - 0.49) | 0.45 (0.43 - 0.48) | 0.21 (0.19 - 0.24) | 0.71 (0.68 - 0.74) |
| PT | 0.76 (0.73 - 0.79) | 0.94 | 0.60 (0.56 - 0.65) | 0.81 (0.79 - 0.83) | 0.76 (0.74 - 0.78) | 0.52 (0.48 - 0.56) | 0.86 (0.84 - 0.88) |
| Left atrial appendage | 0.81 (0.78 - 0.83) | 1.01 | 0.66 (0.61 - 0.70) | 0.83 (0.81 - 0.85) | 0.79 (0.77 - 0.81) | 0.57 (0.53 - 0.61) | 0.88 (0.86 - 0.89) |
| LV | 0.80 (0.77 - 0.82) | 0.80 | 0.70 (0.65 - 0.73) | 0.77 (0.74 - 0.79) | 0.75 (0.73 - 0.77) | 0.50 (0.46 - 0.54) | 0.88 (0.86 - 0.90) |
| Carina | 0.65 (0.62 - 0.68) | 0.86 | 0.41 (0.37 - 0.46) | 0.81 (0.79 - 0.83) | 0.71 (0.69 - 0.73) | 0.42 (0.38 - 0.47) | 0.80 (0.78 - 0.82) |
| Descending aorta | 0.49 (0.46 - 0.52) | -0.88 | 0.75 (0.71 - 0.78) | 0.16 (0.14 - 0.18) | 0.31 (0.29 - 0.33) | 0.23 (0.21 - 0.25) | 0.66 (0.61 - 0.71) |
| Logistic Model | | | | | | | |
| CT ratio + PT + Carina | 0.81 (0.79 - 0.84) | - | 0.71 (0.67 - 0.75) | 0.79 (0.77 - 0.81) | 0.77 (0.75 - 0.79) | 0.53 (0.49 - 0.57) | 0.89 (0.87 - 0.90) |
| CT ratio + PT | 0.81 (0.78 - 0.83) | - | 0.66 (0.61 - 0.70) | 0.84 (0.82 - 0.85) | 0.79 (0.77 - 0.81) | 0.58 (0.53 - 0.62) | 0.88 (0.86 - 0.89) |

AUC = area under the receiver operating characteristic curve; Carina = carina angle; CT = cardiothoracic; LV = left ventricle; PT = pulmonary trunk; SVC/AO = superior vena cava/ascending aorta

**eTable 17. Diagnostic performance in valvular heart disease subgroups**

| Aortic valve disease<br>disease n=1262 (25%)<br>control n=3786 (75%) | AUROC<br>(95% CI) | Cut-off<br>z-score | Sensitivity | Specificity | Accuracy | Positive<br>predictive value | Negative<br>predictive value |
|---|---|---|---|---|---|---|---|
| CT ratio | 0.81 (0.79 - 0.82) | 0.96 | 0.66 (0.64 - 0.69) | 0.81 (0.79 - 0.82) | 0.77 (0.76 - 0.78) | 0.54 (0.51 - 0.56) | 0.88 (0.86 - 0.89) |
| SVC/AO | 0.71 (0.69 - 0.73) | 0.74 | 0.58 (0.55 - 0.61) | 0.74 (0.73 - 0.76) | 0.70 (0.69 - 0.72) | 0.43 (0.41 - 0.46) | 0.84 (0.83 - 0.85) |
| Right atrium | 0.70 (0.68 - 0.72) | 0.72 | 0.55 (0.53 - 0.58) | 0.76 (0.74 - 0.77) | 0.71 (0.69 - 0.72) | 0.43 (0.41 - 0.46) | 0.83 (0.82 - 0.84) |
| Aortic arch | 0.64 (0.62 - 0.66) | 0.78 | 0.45 (0.42 - 0.48) | 0.76 (0.75 - 0.78) | 0.69 (0.67 - 0.70) | 0.39 (0.37 - 0.42) | 0.80 (0.79 - 0.82) |
| PT | 0.59 (0.57 - 0.61) | 1.00 | 0.31 (0.28 - 0.33) | 0.84 (0.83 - 0.85) | 0.71 (0.69 - 0.72) | 0.40 (0.37 - 0.43) | 0.78 (0.77 - 0.79) |
| Left atrial appendage | 0.62 (0.60 - 0.64) | 0.44 | 0.52 (0.49 - 0.54) | 0.68 (0.66 - 0.69) | 0.64 (0.62 - 0.65) | 0.35 (0.33 - 0.37) | 0.81 (0.79 - 0.82) |
| LV | 0.79 (0.77 - 0.81) | 0.80 | 0.66 (0.63 - 0.68) | 0.79 (0.78 - 0.80) | 0.76 (0.74 - 0.77) | 0.51 (0.49 - 0.54) | 0.87 (0.86 - 0.88) |
| Carina | 0.65 (0.63 - 0.67) | 0.45 | 0.54 (0.52 - 0.57) | 0.69 (0.67 - 0.70) | 0.65 (0.64 - 0.67) | 0.37 (0.35 - 0.39) | 0.82 (0.80 - 0.83) |
| Descending aorta | 0.75 (0.74 - 0.77) | 0.73 | 0.72 (0.69 - 0.74) | 0.67 (0.65 - 0.68) | 0.68 (0.67 - 0.69) | 0.42 (0.40 - 0.44) | 0.87 (0.86 - 0.89) |
| Logistic Model | | | | | | | |
| LV + SVC/AO | 0.84 (0.83 - 0.86) | - | 0.75 (0.73 - 0.77) | 0.78 (0.76 - 0.79) | 0.77 (0.76 - 0.78) | 0.53 (0.51 - 0.55) | 0.90 (0.89 - 0.91) |
| LV + SVC/AO + Carina | 0.84 (0.83 - 0.86) | - | 0.71 (0.69 - 0.74) | 0.82 (0.80 - 0.83) | 0.79 (0.78 - 0.80) | 0.57 (0.54 - 0.59) | 0.89 (0.88 - 0.90) |
| Mitral valve disease<br>disease n=2307(25%)<br>control n=6921 (75%) | AUROC<br>(95% CI) | Cut-off<br>z-score | Sensitivity | Specificity | Accuracy | Positive<br>predictive value | Negative<br>predictive value |
| CT ratio | 0.78 (0.77 - 0.79) | 0.94 | 0.64 (0.62 - 0.66) | 0.80 (0.79 - 0.81) | 0.76 (0.75 - 0.77) | 0.52 (0.50 - 0.54) | 0.87 (0.86 - 0.88) |
| SVC/AO | 0.60 (0.59 - 0.62) | 0.59 | 0.46 (0.44 - 0.48) | 0.70 (0.69 - 0.71) | 0.64 (0.63 - 0.65) | 0.34 (0.33 - 0.36) | 0.79 (0.78 - 0.80) |
| Right atrium | 0.70 (0.69 - 0.72) | 0.86 | 0.48 (0.46 - 0.50) | 0.82 (0.81 - 0.83) | 0.74 (0.73 - 0.74) | 0.48 (0.46 - 0.50) | 0.82 (0.81 - 0.83) |
| Aortic arch | 0.51 (0.49 - 0.52) | 1.36 | 0.15 (0.13 - 0.16) | 0.89 (0.88 - 0.90) | 0.70 (0.69 - 0.71) | 0.32 (0.29 - 0.35) | 0.75 (0.75 - 0.76) |
| PT | 0.72 (0.71 - 0.73) | 0.68 | 0.58 (0.56 - 0.60) | 0.75 (0.74 - 0.76) | 0.71 (0.70 - 0.72) | 0.44 (0.42 - 0.46) | 0.84 (0.83 - 0.85) |
| Left atrial appendage | 0.78 (0.77 - 0.80) | 0.73 | 0.68 (0.66 - 0.70) | 0.76 (0.75 - 0.77) | 0.74 (0.73 - 0.75) | 0.49 (0.47 - 0.51) | 0.87 (0.87 - 0.88) |
| LV | 0.76 (0.75 - 0.78) | 0.59 | 0.69 (0.67 - 0.71) | 0.71 (0.70 - 0.72) | 0.71 (0.70 - 0.72) | 0.45 (0.43 - 0.46) | 0.87 (0.86 - 0.88) |
| Carina | 0.71 (0.70 - 0.73) | 0.58 | 0.60 (0.58 - 0.62) | 0.72 (0.71 - 0.73) | 0.69 (0.68 - 0.70) | 0.42 (0.40 - 0.43) | 0.84 (0.83 - 0.85) |
| Descending aorta | 0.72 (0.70 - 0.73) | 1.04 | 0.60 (0.58 - 0.62) | 0.74 (0.73 - 0.75) | 0.71 (0.70 - 0.72) | 0.44 (0.42 - 0.46) | 0.84 (0.84 - 0.85) |
| Logistic Model | | | | | | | |
| CT ratio + PT + Carina | 0.82 (0.81 - 0.83) | - | 0.70 (0.69 - 0.72) | 0.80 (0.79 - 0.81) | 0.77 (0.77 - 0.78) | 0.54 (0.52 - 0.56) | 0.89 (0.88 - 0.89) |
| LV + SVC/AO | 0.84 (0.83 - 0.85) | - | 0.73 (0.71 - 0.74) | 0.81 (0.80 - 0.82) | 0.79 (0.78 - 0.80) | 0.56 (0.55 - 0.58) | 0.90 (0.89 - 0.90) |
| Tricuspid valve disease<br>disease n=300 (25%)<br>control n=900(75%) | AUROC<br>(95% CI) | Cut-off<br>z-score | Sensitivity | Specificity | Accuracy | Positive<br>predictive value | Negative<br>predictive value |

| | | | | | | | |
|---|---|---|---|---|---|---|---|
| CT ratio | 0.87 (0.84 - 0.89) | 0.82 | 0.84 (0.79 - 0.87) | 0.75 (0.72 - 0.78) | 0.77 (0.75 - 0.80) | 0.53 (0.49 - 0.58) | 0.93 (0.91 - 0.95) |
| SVC/AO | 0.67 (0.63 - 0.71) | 0.96 | 0.49 (0.43 - 0.54) | 0.82 (0.79 - 0.84) | 0.74 (0.71 - 0.76) | 0.48 (0.42 - 0.54) | 0.83 (0.80 - 0.85) |
| Right atrium | 0.80 (0.77 - 0.83) | 1.07 | 0.64 (0.58 - 0.69) | 0.86 (0.83 - 0.88) | 0.80 (0.78 - 0.82) | 0.60 (0.55 - 0.66) | 0.87 (0.85 - 0.89) |
| Aortic arch | 0.58 (0.54 - 0.62) | 1.23 | 0.25 (0.21 - 0.30) | 0.87 (0.85 - 0.89) | 0.72 (0.69 - 0.74) | 0.40 (0.34 - 0.48) | 0.77 (0.75 - 0.80) |
| PT | 0.76 (0.73 - 0.80) | 0.92 | 0.63 (0.57 - 0.68) | 0.82 (0.79 - 0.84) | 0.77 (0.75 - 0.79) | 0.54 (0.49 - 0.59) | 0.86 (0.84 - 0.89) |
| Left atrial appendage | 0.80 (0.77 - 0.83) | 0.83 | 0.70 (0.65 - 0.75) | 0.81 (0.79 - 0.84) | 0.78 (0.76 - 0.81) | 0.56 (0.51 - 0.61) | 0.89 (0.87 - 0.91) |
| LV | 0.86 (0.83 - 0.88) | 0.92 | 0.77 (0.72 - 0.81) | 0.81 (0.79 - 0.84) | 0.80 (0.78 - 0.82) | 0.58 (0.53 - 0.63) | 0.91 (0.89 - 0.93) |
| Carina | 0.74 (0.71 - 0.77) | 0.52 | 0.66 (0.60 - 0.71) | 0.71 (0.67 - 0.73) | 0.69 (0.67 - 0.72) | 0.43 (0.38 - 0.47) | 0.86 (0.83 - 0.88) |
| Descending aorta | 0.72 (0.68 - 0.75) | 1.40 | 0.55 (0.50 - 0.61) | 0.83 (0.80 - 0.85) | 0.76 (0.73 - 0.78) | 0.52 (0.46 - 0.57) | 0.84 (0.82 - 0.87) |
| Logistic Model | | | | | | | |
| LV + Right atrium | 0.91 (0.89 - 0.93) | - | 0.81 (0.76 - 0.85) | 0.87 (0.84 - 0.89) | 0.85 (0.83 - 0.87) | 0.67 (0.62 - 0.72) | 0.93 (0.91 - 0.94) |
| CT ratio + PT | 0.89 (0.87 - 0.91) | - | 0.85 (0.81 - 0.89) | 0.80 (0.77 - 0.82) | 0.81 (0.79 - 0.83) | 0.58 (0.54 - 0.63) | 0.94 (0.92 - 0.95) |

AUROC = area under the receiver operating characteristic curve; Carina = carina angle; CT = cardiothoracic; LV = left ventricle; PT = pulmonary trunk; SVC/AO = superior vena cava/ascending aorta

**eTable 18. Adjusted hazard ratio of cardiovascular borders and cardiovascular risk factors for predicting clinical outcome in coronary artery disease group**

| Variables | Adjusted hazard ratio (95% CI) | p value |
|---|---|---|
| CT ratio z-score (vs. <-1) | reference | |
| -1≤ z-score < 0 | 1.39 (0.79-2.46) | 0.249 |
| 0≤ z-score < 1 | 1.94 (1.13-3.34) | 0.016 |
| 1≤ z-score < 2 | 2.16 (1.24-3.75) | 0.006 |
| 2≤ z-score | 3.73 (2.09-6.64) | <0.001 |
| Framingham risk score (vs. low) | | |
| Intermediate | 1.40 (1.12-1.74) | 0.002 |
| High | 1.55 (1.12-2.15) | 0.008 |
| Body mass index (vs. normal) | | |
| Underweight (<18.5 kg/m2) | 2.20 (1.15-4.21) | 0.015 |
| Overweight (≥25.0 kg/m2) | 0.88 (0.71-1.10) | 0.287 |
| Diabetes mellitus | 1.80 (1.46-2.23) | <0.001 |
| Cerebrovascular disease | 0.89 (0.59-1.34) | 0.589 |
| Estimated GFR <60 mL/min | 2.16 (1.70-2.76) | <0.001 |
| Symptoms at CCTA | 1.99 (1.59-2.49) | <0.001 |
| Obstructive CAD on CCTA | 1.58 (1.25-1.98) | <0.001 |
| SVC/AO z-score (vs. <-1) | reference | |
| -1≤ z-score < 0 | 1.21 (0.78-1.86) | 0.378 |
| 0≤ z-score < 1 | 1.49 (0.99-2.25) | 0.054 |
| 1≤ z-score < 2 | 1.97 (1.30-3.00) | 0.001 |
| 2≤ z-score | 2.71 (1.67-4.41) | <0.001 |
| Framingham risk score (vs. low) | | |
| Intermediate | 1.36 (1.09-1.69) | 0.005 |
| High | 1.49 (1.07-2.07) | 0.015 |
| Body mass index (vs. normal) | | |
| Underweight (<18.5 kg/m2) | 2.00 (1.05-3.79) | 0.033 |
| Overweight (≥25.0 kg/m2) | 0.92 (0.74-1.14) | 0.461 |
| Diabetes mellitus | 1.82 (1.48-2.25) | <0.001 |
| Cerebrovascular disease | 0.90 (0.60-1.36) | 0.641 |
| Estimated GFR <60 mL/min | 2.31 (1.82-2.93) | <0.001 |
| Symptoms at CCTA | 2.06 (1.65-2.57) | <0.001 |
| Obstructive CAD on CCTA | 1.58 (1.25-1.98) | <0.001 |
| Right atrium z-score (vs. <-1) | reference | |
| -1≤ z-score < 0 | 1.10 (0.74-1.63) | 0.612 |
| 0≤ z-score < 1 | 1.17 (0.80-1.70) | 0.407 |
| 1≤ z-score < 2 | 1.55 (1.05-2.30) | 0.026 |
| 2≤ z-score | 2.78 (1.79-4.32) | <0.001 |
| Framingham risk score (vs. low) | | |
| Intermediate | 1.39 (1.11-1.73) | 0.003 |
| High | 1.56 (1.13-2.17) | 0.006 |
| Body mass index (vs. normal) | | |
| Underweight (<18.5 kg/m2) | 1.97 (1.04-3.75) | 0.037 |
| Overweight (≥25.0 kg/m2) | 0.94 (0.76-1.16) | 0.581 |
| Diabetes mellitus | 1.82 (1.48-2.25) | <0.001 |
| Cerebrovascular disease | 0.90 (0.60-1.36) | 0.639 |
| Estimated GFR <60 mL/min | 2.33 (1.83-2.95) | <0.001 |
| Symptoms at CCTA | 2.06 (1.65-2.56) | <0.001 |
| Obstructive CAD on CCTA | 1.60 (1.28-2.02) | <0.001 |

| | | |
|---|---|---|
| Aortic arch z-score (vs. <-1) | reference | |
| -1≤ z-score < 0 | 0.80 (0.57-1.12) | 0.202 |
| 0≤ z-score < 1 | 0.82 (0.59-1.14) | 0.254 |
| 1≤ z-score < 2 | 0.77 (0.53-1.11) | 0.164 |
| 2≤ z-score | 0.90 (0.56-1.43) | 0.667 |
| Framingham risk score (vs. low) | | |
| Intermediate | 1.35 (1.09-1.69) | 0.006 |
| High | 1.50 (1.08-2.08) | 0.014 |
| Body mass index (vs. normal) | | |
| Underweight (<18.5 kg/m2) | 1.79 (0.94-3.41) | 0.072 |
| Overweight (≥25.0 kg/m2) | 1.00 (0.81-1.24) | 0.961 |
| Diabetes mellitus | 1.86 (1.50-2.29) | <0.001 |
| Cerebrovascular disease | 0.94 (0.62-1.42) | 0.774 |
| Estimated GFR <60 mL/min | 2.46 (1.94-3.12) | <0.001 |
| Symptoms at CCTA | 2.16 (1.73-2.69) | <0.001 |
| Obstructive CAD on CCTA | 1.62 (1.28-2.03) | <0.001 |
| Pulmonary trunk (vs. <-1) | reference | |
| -1≤ z-score < 0 | 0.64 (0.46-0.89) | 0.008 |
| 0≤ z-score < 1 | 0.73 (0.53-0.99) | 0.045 |
| 1≤ z-score < 2 | 0.88 (0.63-1.23) | 0.485 |
| 2≤ z-score | 1.22 (0.79-1.89) | 0.357 |
| Framingham risk score (vs. low) | | |
| Intermediate | 1.35 (1.08-1.69) | 0.006 |
| High | 1.50 (1.08-2.08) | 0.014 |
| Body mass index (vs. normal) | | |
| Underweight (<18.5 kg/m2) | 1.84 (0.97-3.49) | 0.060 |
| Overweight (≥25.0 kg/m2) | 0.97 (0.78-1.21) | 0.841 |
| Diabetes mellitus | 1.83 (1.48-2.26) | <0.001 |
| Cerebrovascular disease | 0.92 (0.61-1.39) | 0.705 |
| Estimated GFR <60 mL/min | 2.42 (1.91-3.06) | <0.001 |
| Symptoms at CCTA | 2.13 (1.71-2.65) | <0.001 |
| Obstructive CAD on CCTA | 1.62 (1.29-2.03) | <0.001 |
| LA appendage (vs. <-1) | reference | |
| -1≤ z-score < 0 | 0.82 (0.59-1.14) | 0.251 |
| 0≤ z-score < 1 | 0.83 (0.60-1.15) | 0.280 |
| 1≤ z-score < 2 | 1.02 (0.72-1.44) | 0.892 |
| 2≤ z-score | 1.18 (0.74-1.88) | 0.461 |
| Framingham risk score (vs. low) | | |
| Intermediate | 1.35 (1.09-1.69) | 0.006 |
| High | 1.50 (1.08-2.08) | 0.014 |
| Body mass index (vs. normal) | | |
| Underweight (<18.5 kg/m2) | 1.81 (0.95-3.44) | 0.067 |
| Overweight (≥25.0 kg/m2) | 0.97 (0.78-1.21) | 0.841 |
| Diabetes mellitus | 1.83 (1.48-2.27) | <0.001 |
| Cerebrovascular disease | 0.92 (0.61-1.40) | 0.726 |
| Estimated GFR <60 mL/min | 2.44 (1.93-3.10) | <0.001 |
| Symptoms at CCTA | 2.13 (1.71-2.66) | <0.001 |
| Obstructive CAD on CCTA | 1.61 (1.28-2.03) | <0.001 |
| Left ventricle (vs. <-1) | reference | |
| -1≤ z-score < 0 | 1.07 (0.70-1.61) | 0.741 |
| 0≤ z-score < 1 | 0.98 (0.65-1.47) | 0.936 |
| 1≤ z-score < 2 | 1.30 (0.86-1.96) | 0.207 |

| | | |
|---|---|---|
| 2≤ z-score | 1.38 (0.86-2.21) | 0.176 |
| Framingham risk score (vs. low) | | |
| Intermediate | 1.36 (1.09-1.70) | 0.005 |
| High | 1.51 (1.09-2.10) | 0.012 |
| Body mass index (vs. normal) | | |
| Underweight (<18.5 kg/m2) | 1.90 (0.99-3.62) | 0.051 |
| Overweight (≥25.0 kg/m2) | 0.95 (0.76-1.18) | 0.647 |
| Diabetes mellitus | 1.83 (1.48-2.26) | <0.001 |
| Cerebrovascular disease | 0.92 (0.61-1.39) | 0.714 |
| Estimated GFR <60 mL/min | 2.41 (1.90-3.06) | <0.001 |
| Symptoms at CCTA | 2.12 (1.70-2.64) | <0.001 |
| Obstructive CAD on CCTA | 1.60 (1.27-2.01) | <0.001 |
| Descending aorta (vs. <-1) | reference | |
| -1≤ z-score < 0 | 1.20 (0.72-1.99) | 0.472 |
| 0≤ z-score < 1 | 1.45 (0.89-2.34) | 0.128 |
| 1≤ z-score < 2 | 1.50 (0.92-2.44) | 0.096 |
| 2≤ z-score | 2.04 (1.25-3.32) | 0.003 |
| Framingham risk score (vs. low) | | |
| Intermediate | 1.31 (1.05-1.64) | 0.013 |
| High | 1.43 (1.03-1.98) | 0.031 |
| Body mass index (vs. normal) | | |
| Underweight (<18.5 kg/m2) | 2.01 (1.06-3.83) | 0.031 |
| Overweight (≥25.0 kg/m2) | 0.95 (0.77-1.18) | 0.685 |
| Diabetes mellitus | 1.87 (1.52-2.31) | <0.001 |
| Cerebrovascular disease | 0.89 (0.59-1.35) | 0.600 |
| Estimated GFR <60 mL/min | 2.24 (1.76-2.86) | <0.001 |
| Symptoms at CCTA | 2.09 (1.67-2.60) | <0.001 |
| Obstructive CAD on CCTA | 1.59 (1.27-2.00) | <0.001 |
| Carinal angle (vs. <-1) | reference | |
| -1≤ z-score < 0 | 0.99 (0.69-1.42) | 0.965 |
| 0≤ z-score < 1 | 1.38 (0.98-1.94) | 0.061 |
| 1≤ z-score < 2 | 1.82 (1.27-2.60) | 0.001 |
| 2≤ z-score | 2.68 (1.75-4.10) | <0.001 |
| Framingham risk score (vs. low) | | |
| Intermediate | 1.37 (1.10-1.71) | 0.004 |
| High | 1.52 (1.09-2.10) | 0.011 |
| Body mass index (vs. normal) | | |
| Underweight (<18.5 kg/m2) | 1.90 (1.00-3.61) | 0.047 |
| Overweight (≥25.0 kg/m2) | 0.94 (0.76-1.16) | 0.581 |
| Diabetes mellitus | 1.81 (1.46-2.23) | <0.001 |
| Cerebrovascular disease | 0.89 (0.59-1.34) | 0.583 |
| Estimated GFR <60 mL/min | 2.36 (1.86-3.00) | <0.001 |
| Symptoms at CCTA | 2.09 (1.68-2.61) | <0.001 |
| Obstructive CAD on CCTA | 1.64 (1.30-2.06) | <0.001 |

CAD = coronary artery disease; CCTA = coronary computed tomography angiography; GFR = glomerular filtration rate

**eFigure 11. All-cause death or myocardial infarction stratified by superior vena cava/ascending aorta (SVC/AO) in chest X-ray**

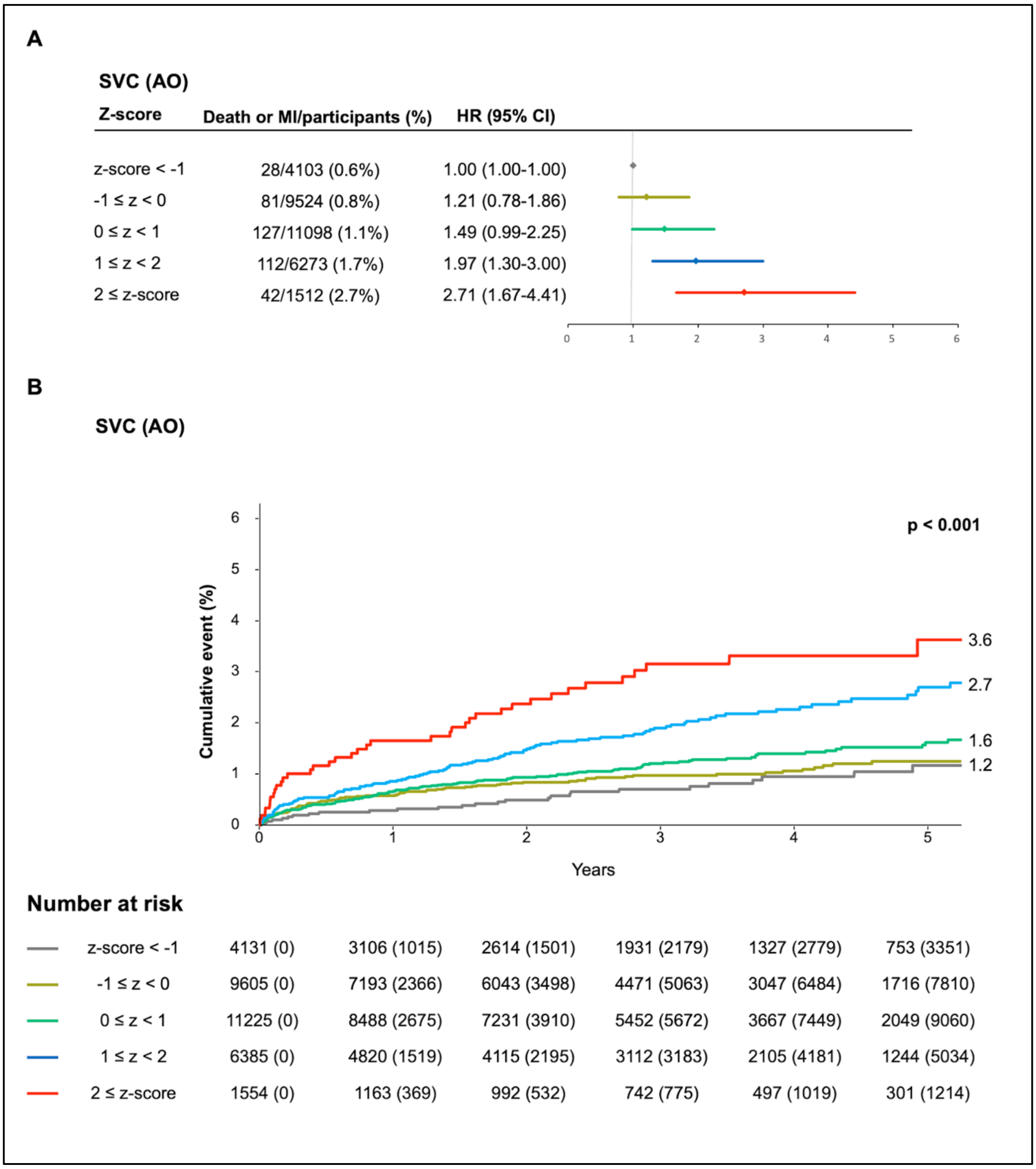

**eFigure 12. All-cause death or myocardial infarction stratified by right atrium in chest X-ray**

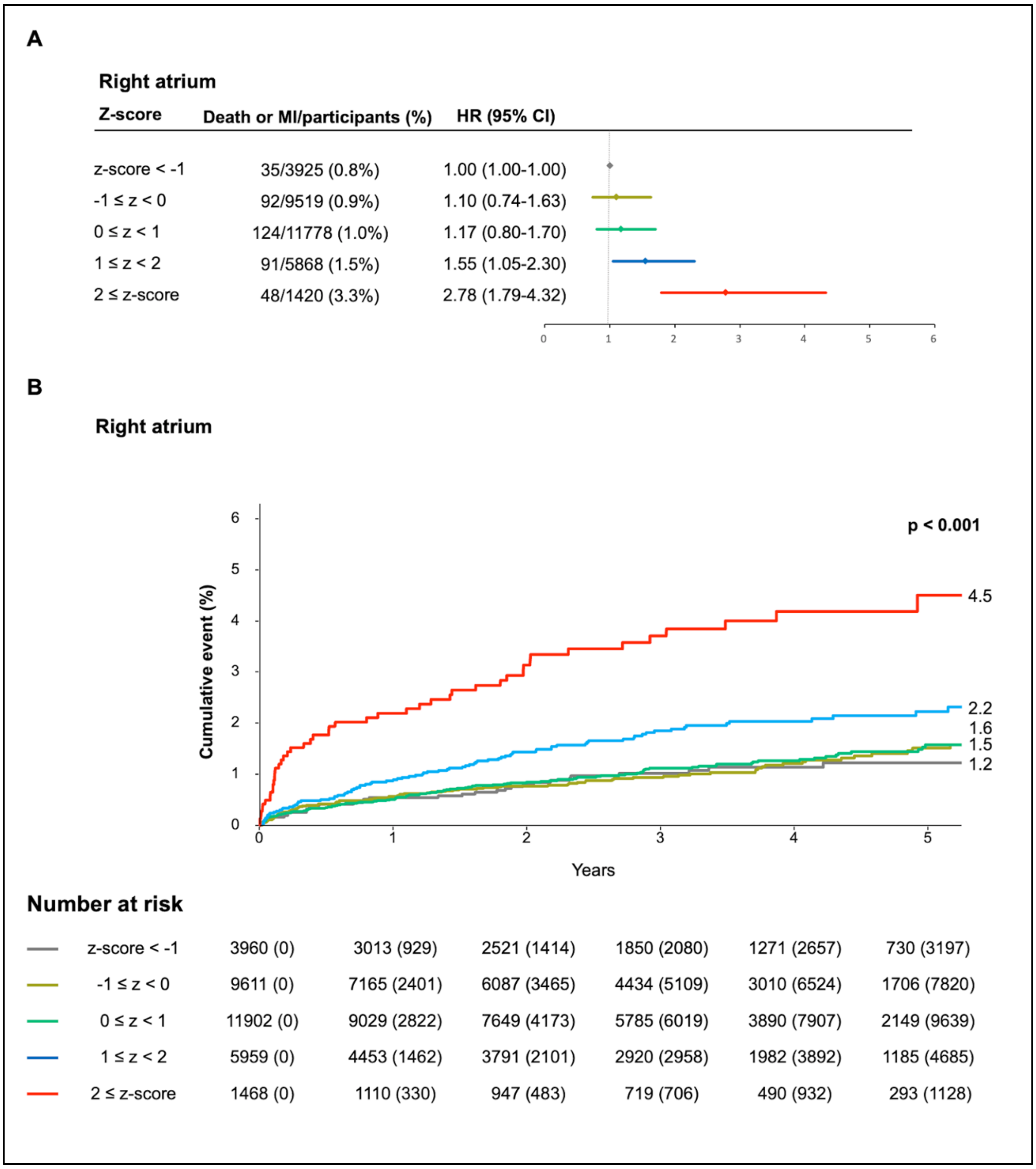

**eFigure 13. All-cause death or myocardial infarction stratified by aortic arch in chest X-ray**

**A**

**Aortic arch**

| Z-score | Death or MI/participants (%) | HR (95% CI) |
|---|---|---|
| z-score < -1 | 50/3914 (1.2%) | 1.00 (1.00-1.00) |
| -1 ≤ z < 0 | 105/9639 (1.0%) | 0.80 (0.57-1.12) |
| 0 ≤ z < 1 | 133/11225 (1.1%) | 0.82 (0.59-1.14) |
| 1 ≤ z < 2 | 74/6044 (1.2%) | 0.77 (0.53-1.11) |
| 2 ≤ z-score | 28/1688 (1.6%) | 0.90 (0.56-1.43) |

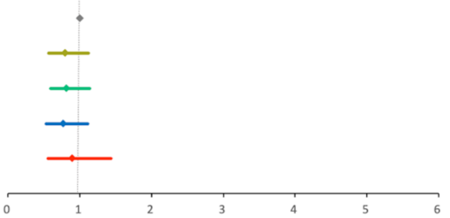

**B**

**Aortic arch**

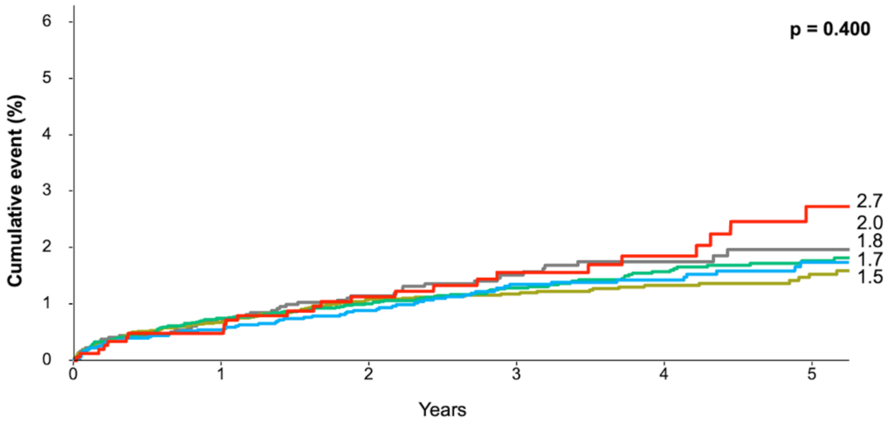

**Number at risk**

| | | | | | | |
|---|---|---|---|---|---|---|
| z-score < -1 | 3964 (0) | 2887 (1053) | 2453 (1476) | 1808 (2113) | 1181 (2736) | 638 (3277) |
| -1 ≤ z < 0 | 9744 (0) | 7351 (2337) | 6216 (3444) | 4672 (4983) | 3140 (6510) | 1738 (7907) |
| 0 ≤ z < 1 | 11358 (0) | 8627 (2660) | 7327 (3939) | 5423 (5825) | 3681 (7554) | 2106 (9123) |
| 1 ≤ z < 2 | 6118 (0) | 4624 (1466) | 3919 (2156) | 2980 (3079) | 2061 (3997) | 1226 (4826) |
| 2 ≤ z-score | 1716 (0) | 1281 (428) | 1080 (621) | 825 (872) | 580 (1115) | 355 (1336) |

**eFigure 14. All-cause death or myocardial infarction stratified by pulmonary trunk in chest X-ray**

**A**

**Pulmonary trunk**

| Z-score | Death or MI/participants (%) | HR (95% CI) |
|---|---|---|
| z-score < -1 | 62/3994 (1.5%) | 1.00 (1.00-1.00) |
| -1 ≤ z < 0 | 93/9825 (0.9%) | 0.64 (0.46-0.89) |
| 0 ≤ z < 1 | 121/11426 (1.0%) | 0.73 (0.53-0.99) |
| 1 ≤ z < 2 | 83/5869 (1.4%) | 0.88 (0.63-1.23) |
| 2 ≤ z-score | 31/1396 (2.2%) | 1.22 (0.79-1.89) |

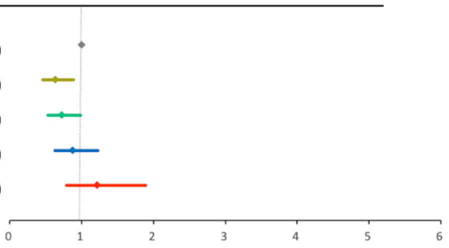

**B**

**Pulmonary trunk**

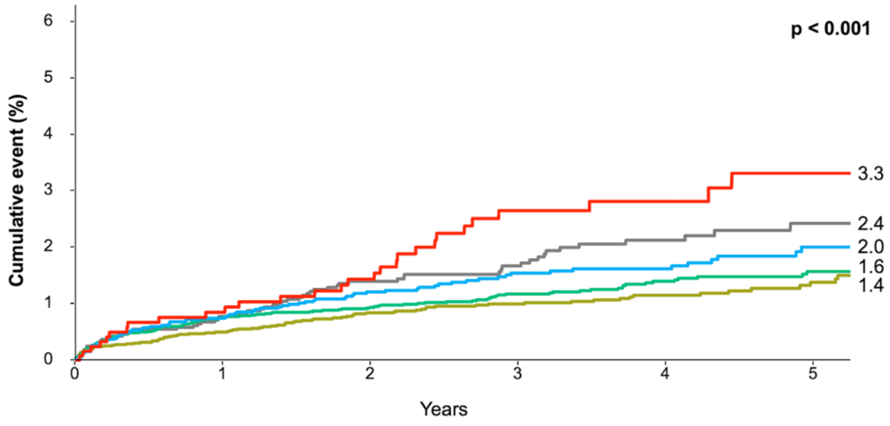

**Number at risk**

| | 0 | 1 | 2 | 3 | 4 | 5 |
|---|---|---|---|---|---|---|
| z-score < -1 | 4056 (0) | 3017 (1013) | 2556 (1456) | 1906 (2100) | 1283 (2716) | 724 (3271) |
| -1 ≤ z < 0 | 9918 (0) | 7427 (2450) | 6322 (3531) | 4725 (5119) | 3175 (6663) | 1766 (8067) |
| 0 ≤ z < 1 | 11547 (0) | 8718 (2757) | 7409 (4051) | 5521 (5924) | 3756 (7680) | 2115 (9315) |
| 1 ≤ z < 2 | 5952 (0) | 4532 (1383) | 3796 (2099) | 2877 (3007) | 1957 (3925) | 1164 (4712) |
| 2 ≤ z-score | 1427 (0) | 1076 (341) | 912 (499) | 679 (722) | 472 (928) | 294 (1104) |

**eFigure 15. All-cause death or myocardial infarction stratified by left atrial (LA) appendage in chest X-ray**

**A**

**LA appendage**

| Z-score | Death or MI/participants (%) | HR (95% CI) |
|---|---|---|
| z-score < -1 | 55/3969 (1.3%) | 1.00 (1.00-1.00) |
| -1 ≤ z < 0 | 102/9690 (1.0%) | 0.82 (0.59-1.14) |
| 0 ≤ z < 1 | 121/11388 (1.0%) | 0.83 (0.60-1.15) |
| 1 ≤ z < 2 | 84/6013 (1.4%) | 1.02 (0.72-1.44) |
| 2 ≤ z-score | 28/1450 (1.9%) | 1.18 (0.74-1.88) |

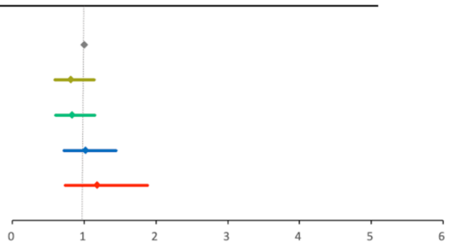

**B**

**LA appendage**

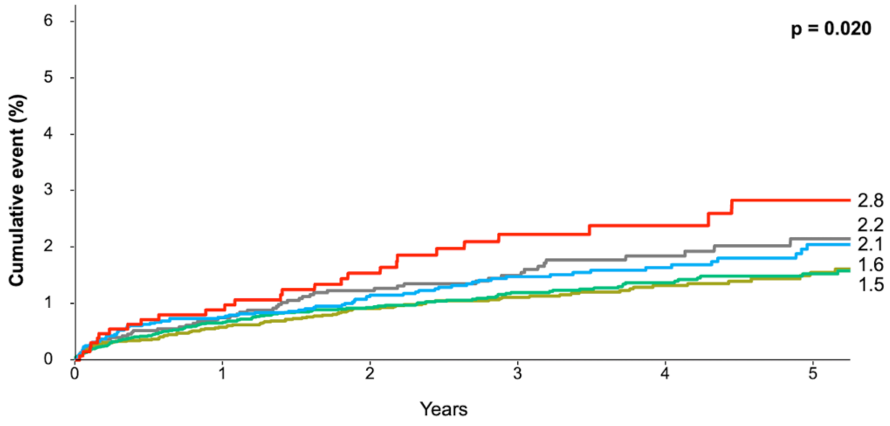

**Number at risk**

| | | | | | | |
|---|---|---|---|---|---|---|
| z-score < -1 | 4024 (0) | 2997 (1002) | 2543 (1442) | 1866 (2113) | 1258 (2715) | 728 (3242) |
| -1 ≤ z < 0 | 9792 (0) | 7346 (2399) | 6248 (3474) | 4686 (5025) | 3148 (6556) | 1716 (7982) |
| 0 ≤ z < 1 | 11509 (0) | 8658 (2787) | 7340 (4083) | 5501 (5905) | 3732 (7666) | 2129 (9264) |
| 1 ≤ z < 2 | 6097 (0) | 4645 (1413) | 3905 (2136) | 2929 (3101) | 1983 (4044) | 1164 (4856) |
| 2 ≤ z-score | 1478 (0) | 1124 (343) | 959 (501) | 726 (728) | 522 (931) | 326 (1125) |

**eFigure 16. All-cause death or myocardial infarction stratified by left ventricle in chest X-ray**

**A**

**Left ventricle**

| Z-score | Death or MI/participants (%) | HR (95% CI) |
|---|---|---|
| z-score < -1 | 31/3320 (0.9%) | 1.00 (1.00-1.00) |
| -1 ≤ z < 0 | 89/8456 (1.0%) | 1.07 (0.70-1.61) |
| 0 ≤ z < 1 | 117/11308 (1.0%) | 0.98 (0.65-1.47) |
| 1 ≤ z < 2 | 107/7022 (1.5%) | 1.30 (0.86-1.96) |
| 2 ≤ z-score | 46/2404 (1.9%) | 1.38 (0.86-2.21) |

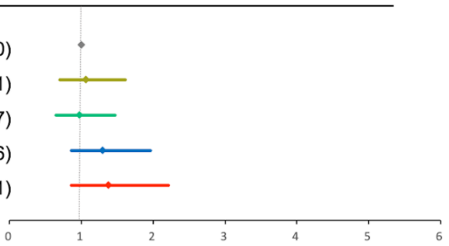

**B**

**Left ventricle**

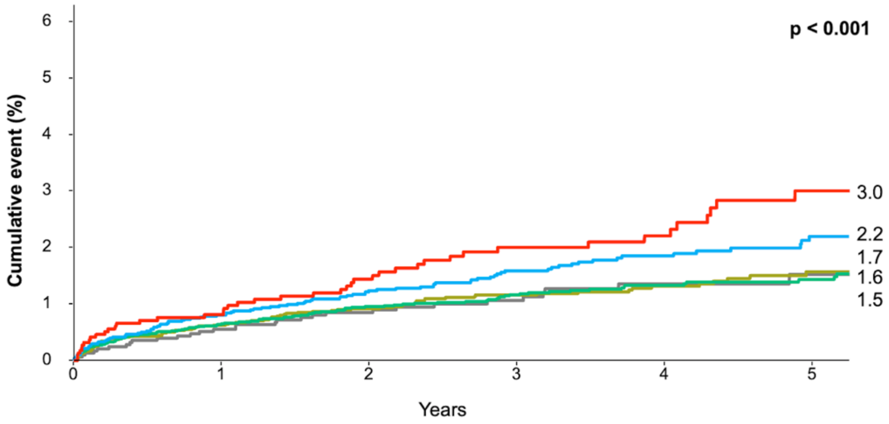

**Number at risk**

| | | | | | | |
|---|---|---|---|---|---|---|
| z-score < -1 | 3351 (0) | 2465 (871) | 2077 (1252) | 1501 (1824) | 993 (2329) | 553 (2767) |
| -1 ≤ z < 0 | 8545 (0) | 6410 (2091) | 5433 (3050) | 4030 (4441) | 2691 (5575) | 1445 (7016) |
| 0 ≤ z < 1 | 11425 (0) | 8650 (2713) | 7351 (3987) | 5494 (5831) | 3718 (7599) | 2117 (9197) |
| 1 ≤ z < 2 | 7129 (0) | 5399 (1682) | 4571 (2488) | 3493 (3552) | 2380 (4657) | 1387 (5644) |
| 2 ≤ z-score | 2450 (0) | 1846 (587) | 1563 (859) | 1190 (1224) | 861 (1552) | 561 (1845) |

**eFigure 17. All-cause death or myocardial infarction stratified by descending aorta in chest X-ray**

**A**

**Descending aorta**

| Z-score | Death or MI/participants (%) | HR (95% CI) |
|---|---|---|
| z-score < -1 | 20/3141 (0.6%) | 1.00 (1.00-1.00) |
| -1 ≤ z < 0 | 62/7758 (0.8%) | 1.20 (0.72-1.99) |
| 0 ≤ z < 1 | 102/9499 (1.0%) | 1.45 (0.89-2.34) |
| 1 ≤ z < 2 | 102/7469 (1.3%) | 1.50 (0.92-2.44) |
| 2 ≤ z-score | 104/4643 (2.2%) | 2.04 (1.25-3.32) |

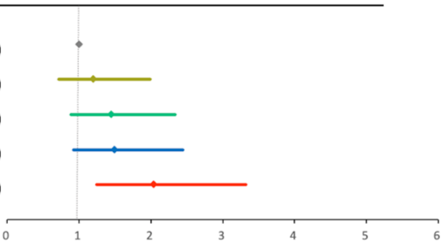

**B**

**Descending aorta**

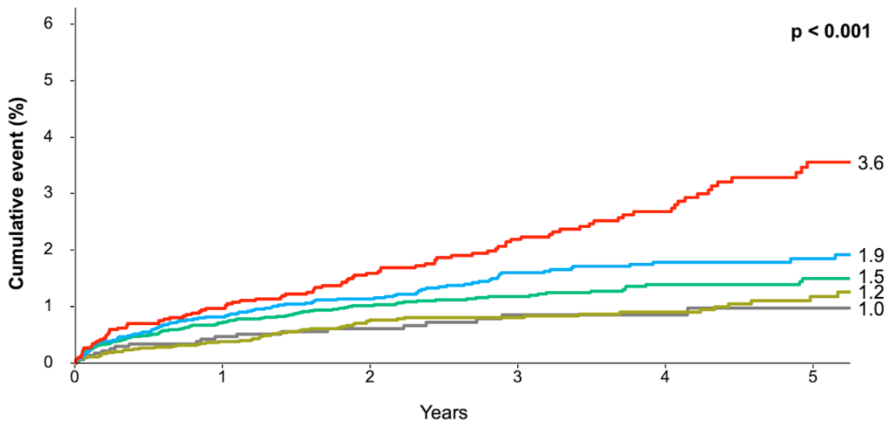

**Number at risk**

| | | | | | | |
|---|---|---|---|---|---|---|
| z-score < -1 | 3161 (0) | 2264 (885) | 1899 (1247) | 1385 (1757) | 935 (2207) | 466 (2675) |
| -1 ≤ z < 0 | 7820 (0) | 5929 (1866) | 4980 (2794) | 3697 (4075) | 2440 (5330) | 1339 (6425) |
| 0 ≤ z < 1 | 9601 (0) | 7228 (2315) | 6158 (3365) | 4584 (4930) | 3079 (6427) | 1725 (7779) |
| 1 ≤ z < 2 | 7571 (0) | 5768 (1751) | 4909 (2592) | 3732 (3749) | 2568 (4907) | 1510 (5964) |
| 2 ≤ z-score | 4747 (0) | 3581 (1127) | 3049 (1638) | 2310 (2361) | 1621 (3041) | 1023 (3626) |

**eFigure 18. All-cause death or myocardial infarction stratified by carinal angle in chest X-ray**

**A**

**Carinal angle**

| Z-score | Death or MI/participants (%) | HR (95% CI) |
|---|---|---|
| z-score < -1 | 45/4933 (0.9%) | 1.00 (1.00-1.00) |
| -1 ≤ z < 0 | 86/10102 (0.8%) | 0.99 (0.69-1.42) |
| 0 ≤ z < 1 | 126/10762 (1.1%) | 1.38 (0.98-1.94) |
| 1 ≤ z < 2 | 91/5332 (1.7%) | 1.82 (1.27-2.60) |
| 2 ≤ z-score | 42/1381 (3.0%) | 2.68 (1.75-4.10) |

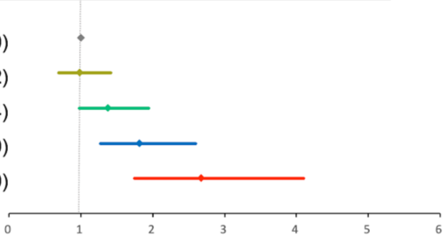

**B**

**Carinal angle**

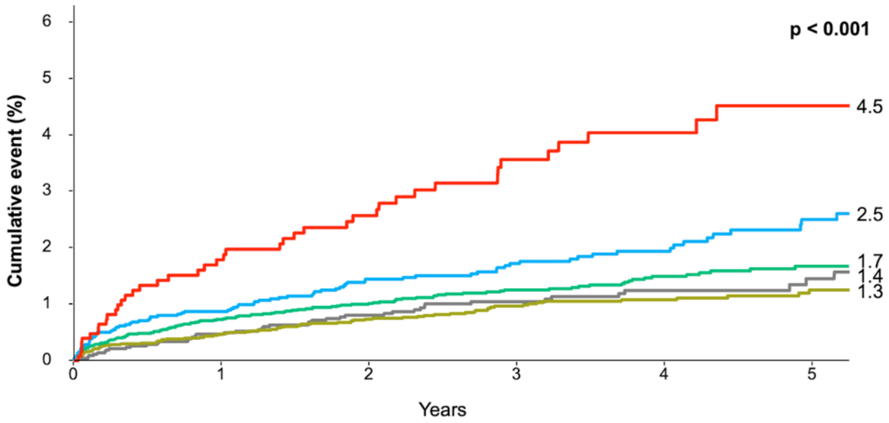

**Number at risk**

| | | | | | | |
|---|---|---|---|---|---|---|
| z-score < -1 | 4978 (0) | 3762 (1197) | 3184 (1763) | 2365 (2575) | 1589 (3347) | 908 (4026) |
| -1 ≤ z < 0 | 10188 (0) | 7660 (2488) | 6526 (3603) | 4853 (5263) | 3298 (6814) | 1832 (8275) |
| 0 ≤ z < 1 | 10888 (0) | 8209 (2613) | 6969 (3831) | 5225 (5560) | 3512 (7264) | 2040 (8730) |
| 1 ≤ z < 2 | 5423 (0) | 4083 (1300) | 3419 (1942) | 2595 (2758) | 1769 (3579) | 1016 (4324) |
| 2 ≤ z-score | 1423 (0) | 1056 (346) | 897 (497) | 670 (716) | 475 (908) | 267 (1114) |

**eFigure 19. (Case 1) Aortic stenosis with dilated ascending aorta**

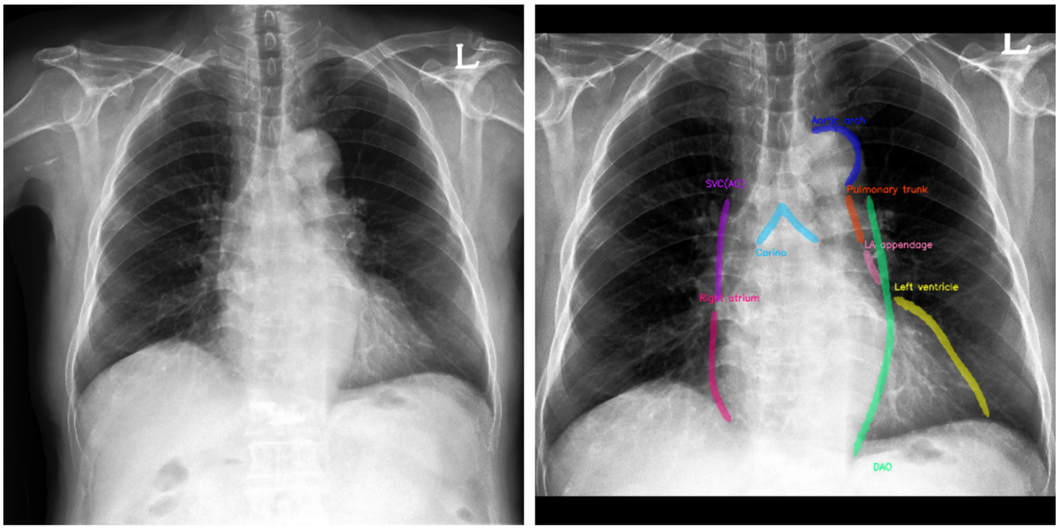

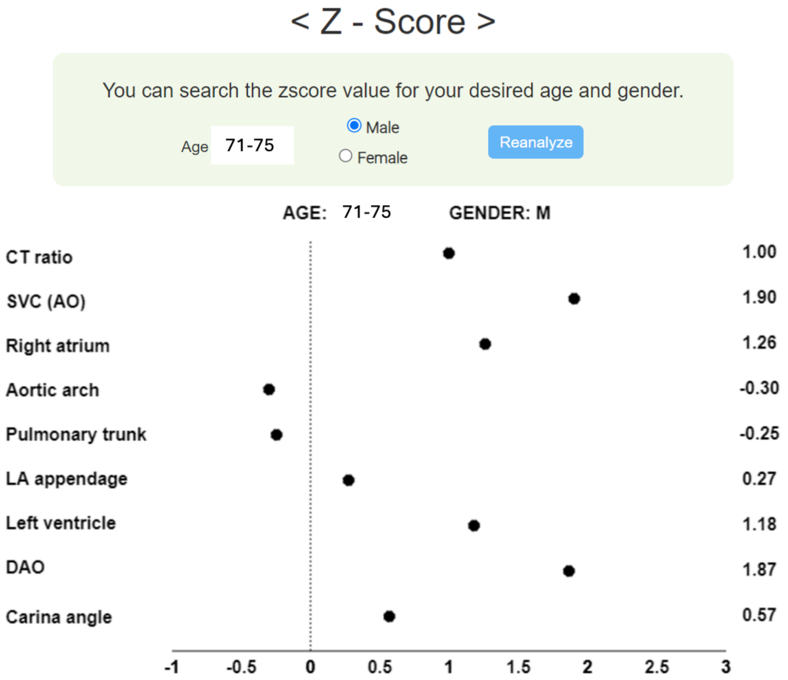

| Case Summary | Severe degenerative aortic stenosis, Age range 71-75/M |
|---|---|
| Echocardiography | Transaortic valvular velocity: 4.1m/sec<br>LV hypertrophy: LV mass 233.7g<br>Dilated ascending aorta: 43mm |
| CXR interpretation | The z-score for SVC/AO has increased to 1.9, consistent with the appearance of a dilated ascending aorta. The z-scores for the CT ratio, RA, LV, and DAO have also generally increased. |

**eFigure 20. (Case 2) Mitral stenosis and regurgitation with dilated both left atrium and ventricle combined with pulmonary hypertension**

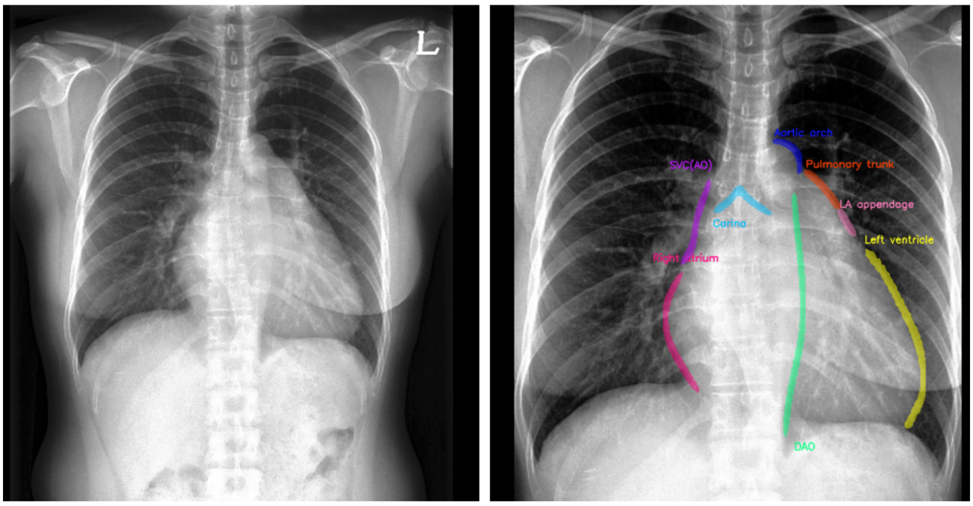

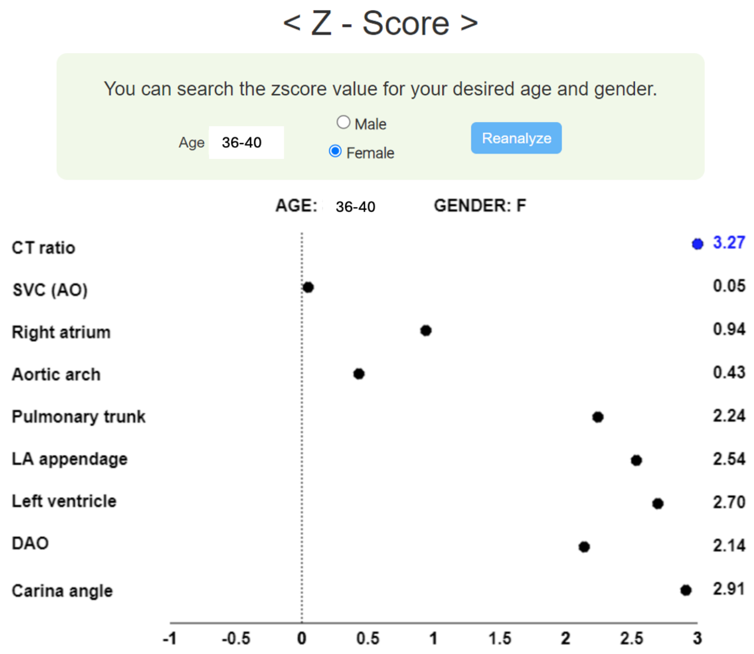

| Case Summary | Mitral stenosis and regurgitation, Age range 36-40/F |
|---|---|
| Echocardiography | Rheumatic valvular heart disease with severe mitral stenosis<br>Mild to moderate regurgitation involving mitral, aortic, and tricuspid valve<br>Severe resting pulmonary hypertension (TR velocity 4.2 m/sec)<br>Enlarged LA (51mm) |
| CXR interpretation | The CT ratio z-score of 3.27 indicates severe cardiomegaly. There are significant increases in the LA appendage (z-score 2.54) and carina angle (2.91), suggesting left atrial enlargement. The enlargement of the LV (z-score 2.70) is thought to be a result of associated mitral regurgitation. Increases in the pulmonary trunk (2.24) and RA (0.94) are considered to reflect the presence of associated pulmonary hypertension. |

**eFigure 21. (Case 3) Aortic regurgitation with dilated ascending aorta**

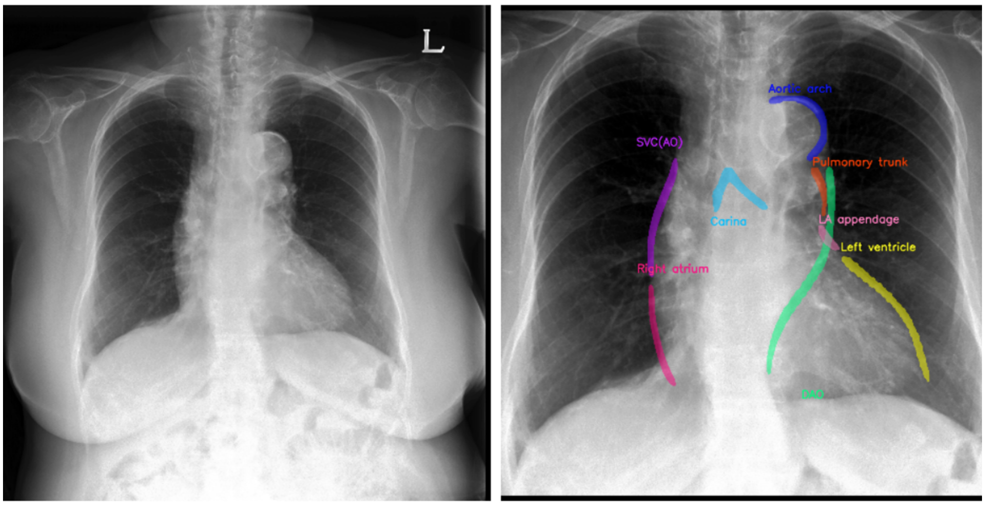

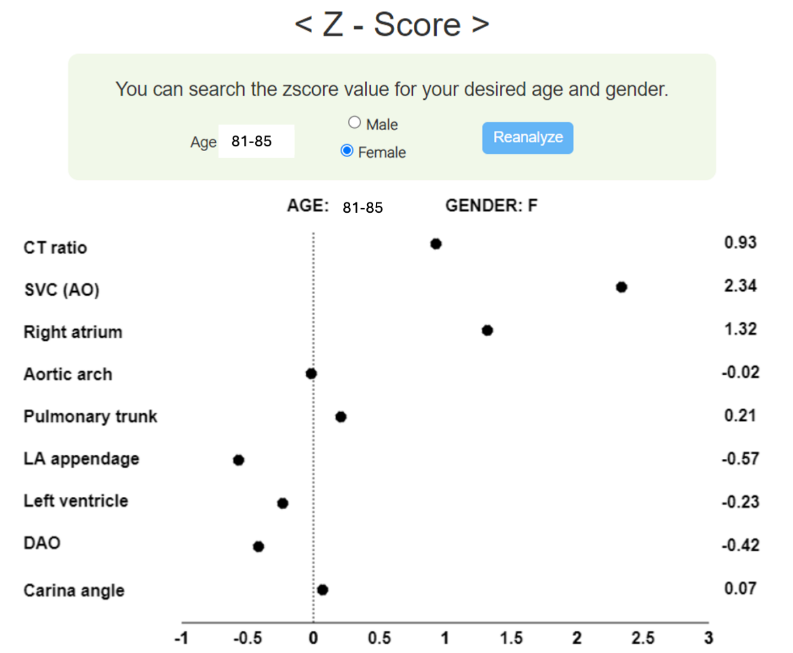

| | |
|---|---|
| Case Summary | Mild aortic regurgitation with dilatation ascending aorta, Age range 81-85/F |
| Echocardiography | Dilated ascending aorta: 47mm<br>Normal LV end-diastolic volume 69 ml and LV mass 123.0 g<br>Mild aortic regurgitation |
| CXR interpretation | The CT ratio shows a borderline increase (z-score 0.93). The SVC/AO is significantly increased at 2.34. On the CXR, the RA border has increased to 1.32, which is likely due to the enlarged boundary area caused by the aorta shadow. According to echocardiography findings, there was no increase in the right atrium. |

**eFigure 22. (Case 4) Atrial Septal Defect with dilated pulmonary trunk**

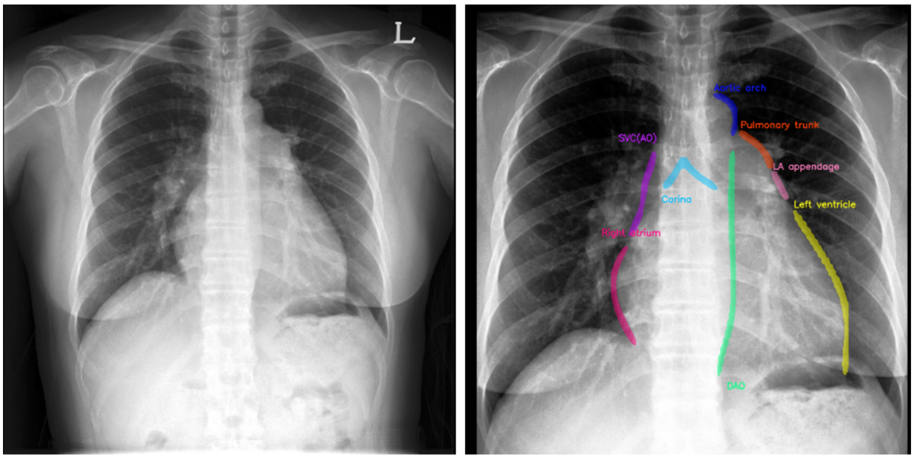

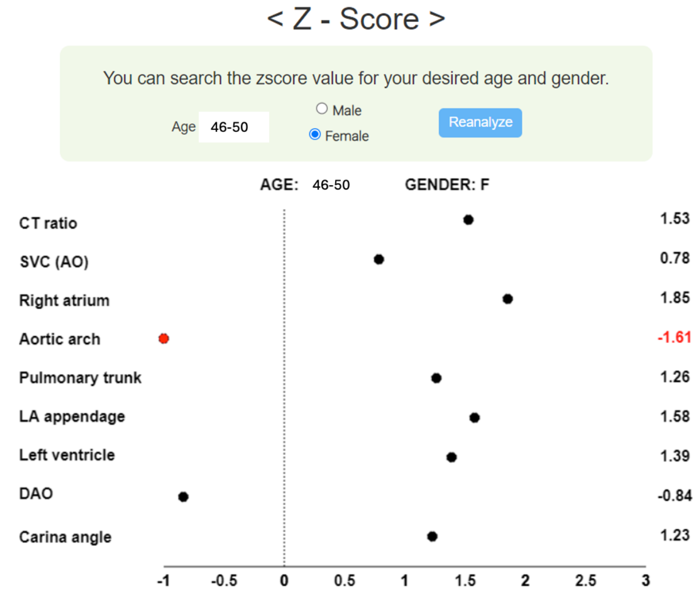

| Case Summary | Large secundum atrial septal defect, Age range 55-60/F |
|---|---|
| Echocardiography | Large secundum ASD with left-to-right shunt<br>Moderate tricuspid regurgitation with moderate resting pulmonary hypertension (TR velocity 3.4 m/s)<br>Enlarged RV; normal left ventricle; borderline LA (38mm) |
| CXR interpretation | On the CXR, the increased borders of RA (z-score 1.85) and pulmonary trunk (1.26) align well with the presence of right cardiac chamber enlargement and pulmonary hypertension on echocardiography. Interestingly, the aortic arch shows a decrease (-1.61), which may be associated with reduced left ventricular output due to a left-to-right shunt. Echocardiography shows the LV size as normal, suggesting that the increased LV border (1.39) could be a secondary finding due to RV enlargement. The enlargement in the LAA (1.58) is thought to be more a result of its proximity to the enlarged pulmonary trunk rather than a direct indication of LA enlargement. |

**eFigure 23. (Case 5) Aortic Dissection with Aneurysm in the Arch and Descending Aorta.**

**A.**

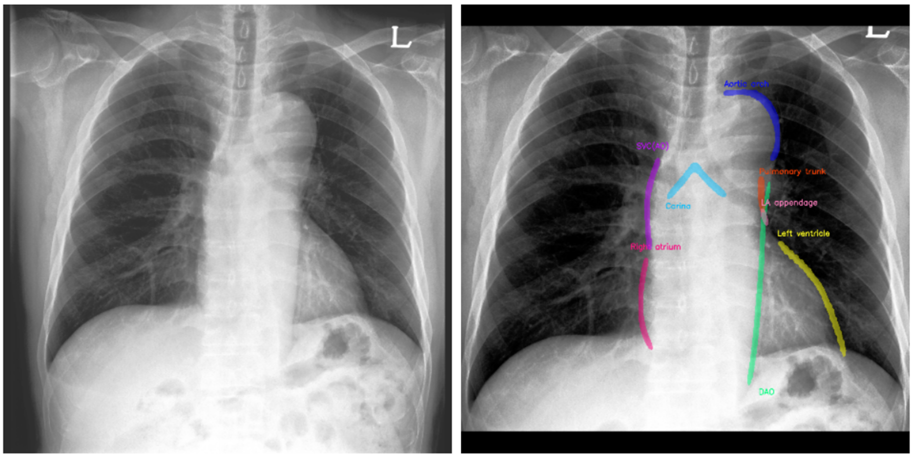

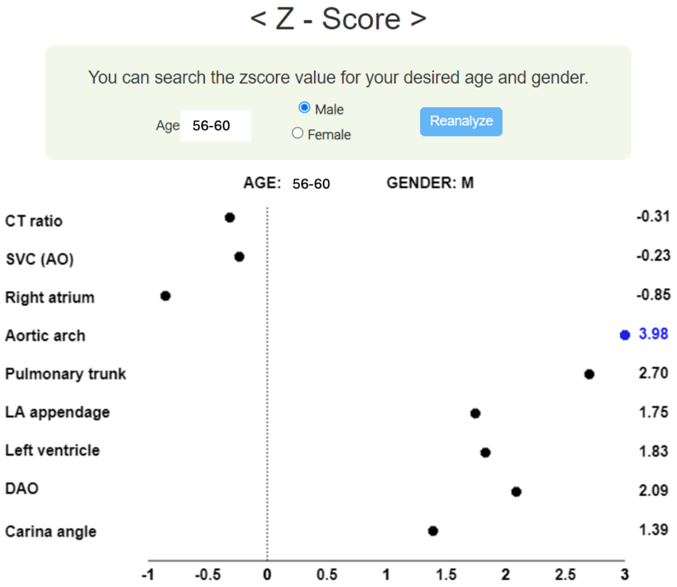

| Case Summary | Aortic dissection with aneurysm, Age range 55-60/M |
|---|---|
| Aorta CT | Type B aortic dissection<br>Dilated proximal descending thoracic aorta (49mm), just behind left subclavian artery origin |
| Echocardiography | Global hypokinesia in LV (LV ejection fraction 38%)<br>LV EDV 118 ml |
| CXR interpretation | On the CXR, the z-score for the aortic arch significantly increased to 3.98, aligning well with the dilation of the proximal descending aorta observed on CT. The z-score for the descending aorta (DAO) also increased to 2.09. The CT ratio (-0.31) did not increase, but the LV (1.83) showed an increase, which could be associated with the LV dilatation and dysfunction observed in echocardiography. The increase in the pulmonary trunk (2.70) observed on the CXR could not be explained by findings on CT and echocardiography, suggesting it may be a false positive finding. |

**B.**

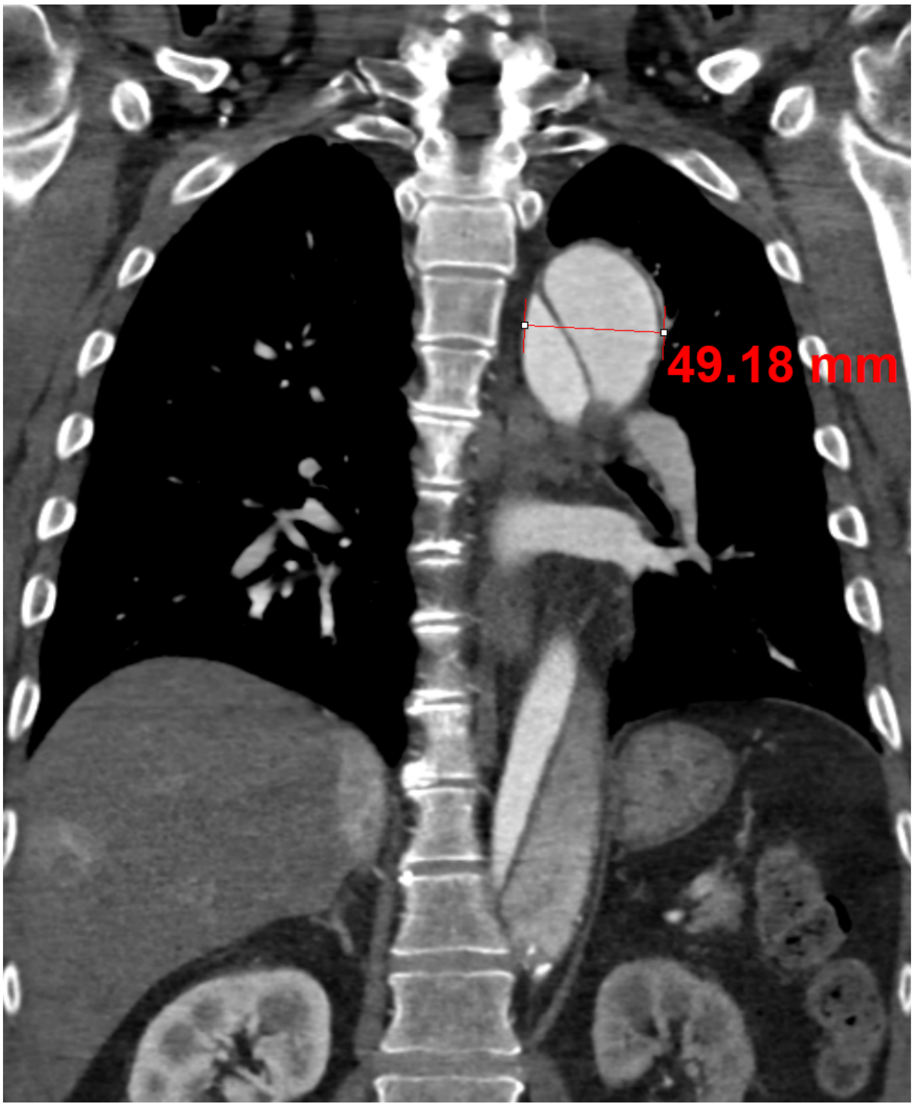

**eFigure 24. (Case 6) Mediastinal lymphoma showing positive silhouette sign between mass and right cardiac border of SVC/AO**

**A.**

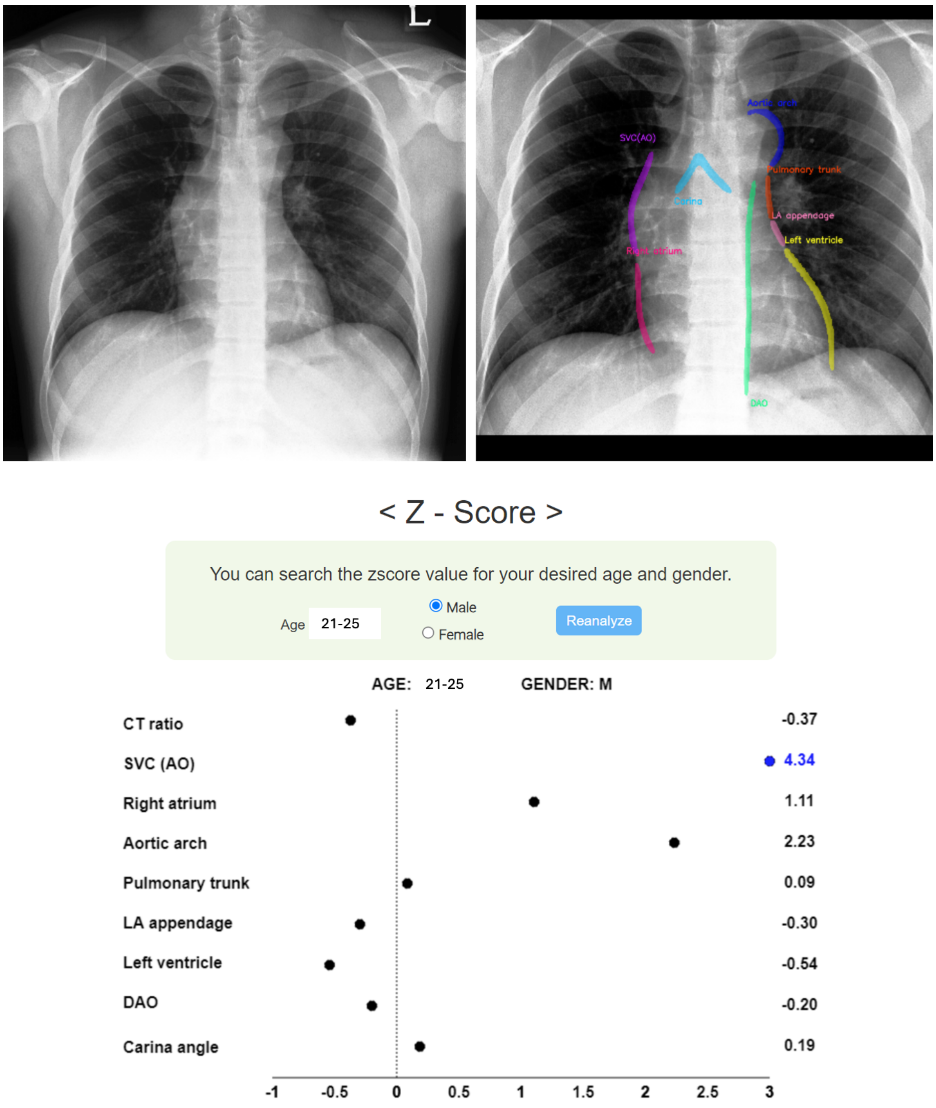

| Case Summary | Anterior mediastinal lymphoma, Age range 20-25/M |
|---|---|
| Chest CT | Mediastinal mass (lymphoma) in abutting SVC, ascending aorta, and aortic arch |
| Echocardiography | Normal heart and thoracic aorta |
| CXR interpretation | On the CXR, the boundary between the mass and SVC/AO is obscured, making it indistinguishable, which means a positive silhouette sign. The cardiovascular border analysis AI recognized the boundary of the mass as part of the SVC(AO), resulting in a significantly increased z-score of 4.34. The mass appears distinct from the aortic arch (red dotted line), indicating a negative silhouette sign. However, the AI misinterpreted the mass boundary, which lies further out, as the border of the arch, leading to an increased z-score of 2.23 for the arch. Other metrics like the CT ratio, pulmonary trunk, and LV were all within normal ranges. |

**B.**

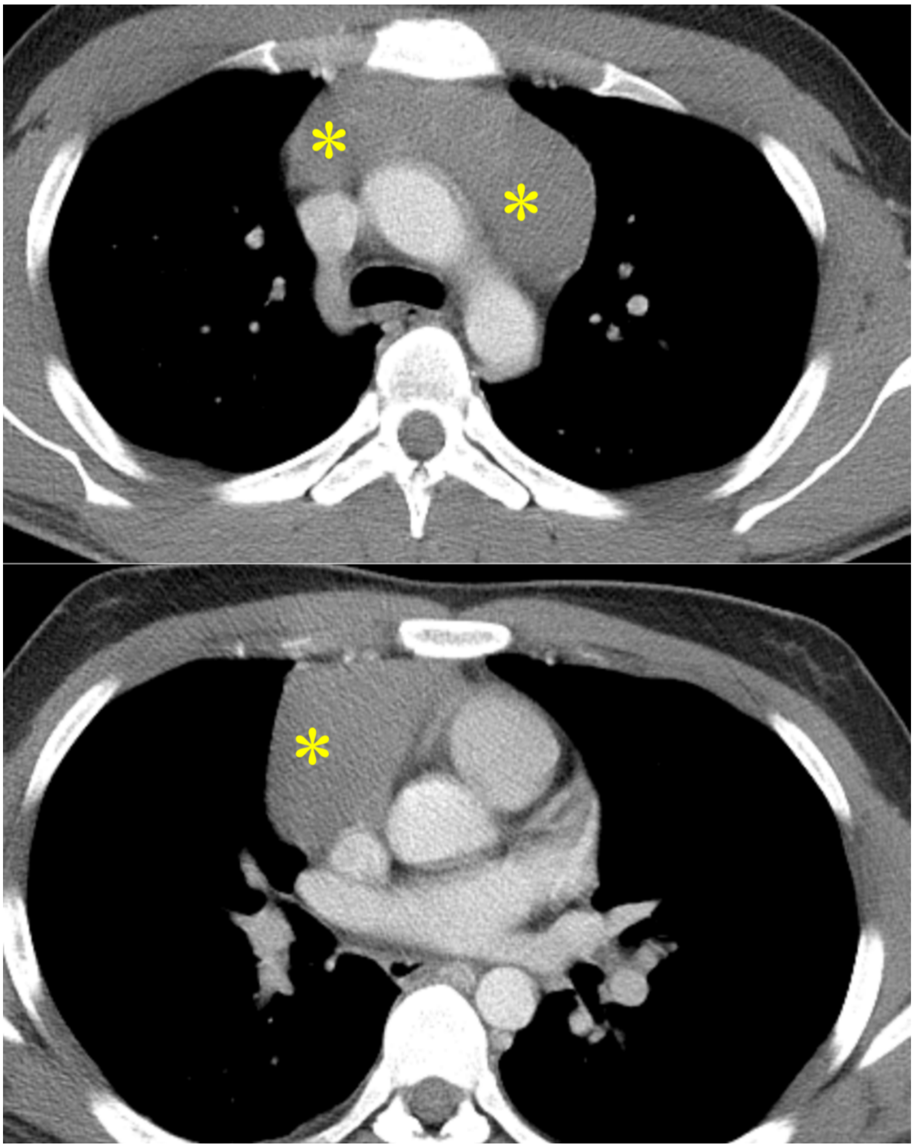

**eFigure 25. (Case 7) Mediastinal thymoma showing negative silhouette sign between mass and pulmonary trunk**

**A.**

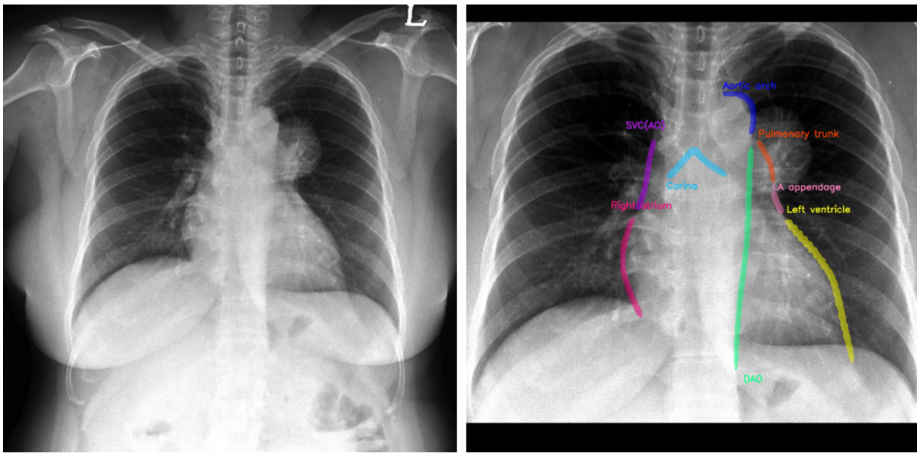

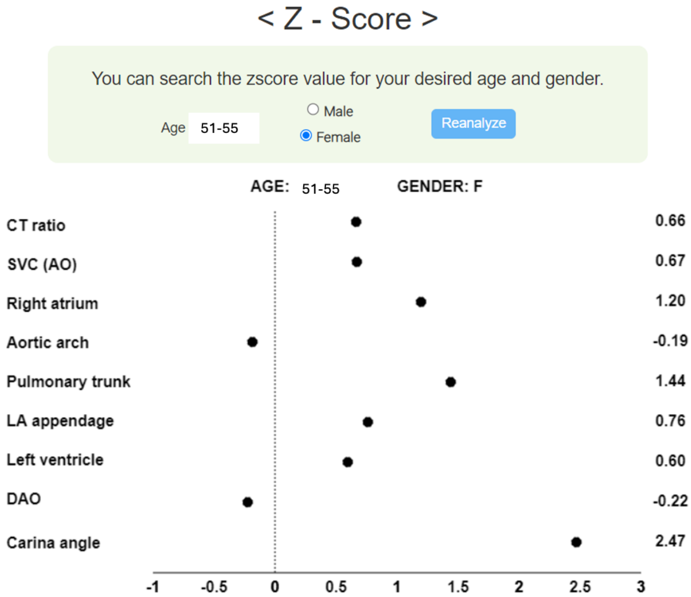

| | |
|---|---|
| Case Summary | Mediastinal thymoma showing negative silhouette sign, Age range 51-55/F |
| Chest CT | Anterior mediastinal thymoma |
| Echocardiography | Borderline, resting pulmonary hypertension (TR velocity 3.0 m/s)<br>Dilated left atrium (40 mm) |
| CXR interpretation | On the CXR, a mass shadow maintaining its border with the pulmonary trunk shadow (silhouette sign negative) was observed, which was confirmed as a thymoma located in the left-sided anterior mediastinum (Figure S25B). The cardiovascular border analysis AI accurately delineated the border of the pulmonary trunk and did not react to the mass shadow. Increased z-scores for the pulmonary trunk (1.44), RA (1.20), and carinal angle (2.47) appear to align with the echocardiography findings of pulmonary hypertension and borderline LA enlargement. |

**B.**

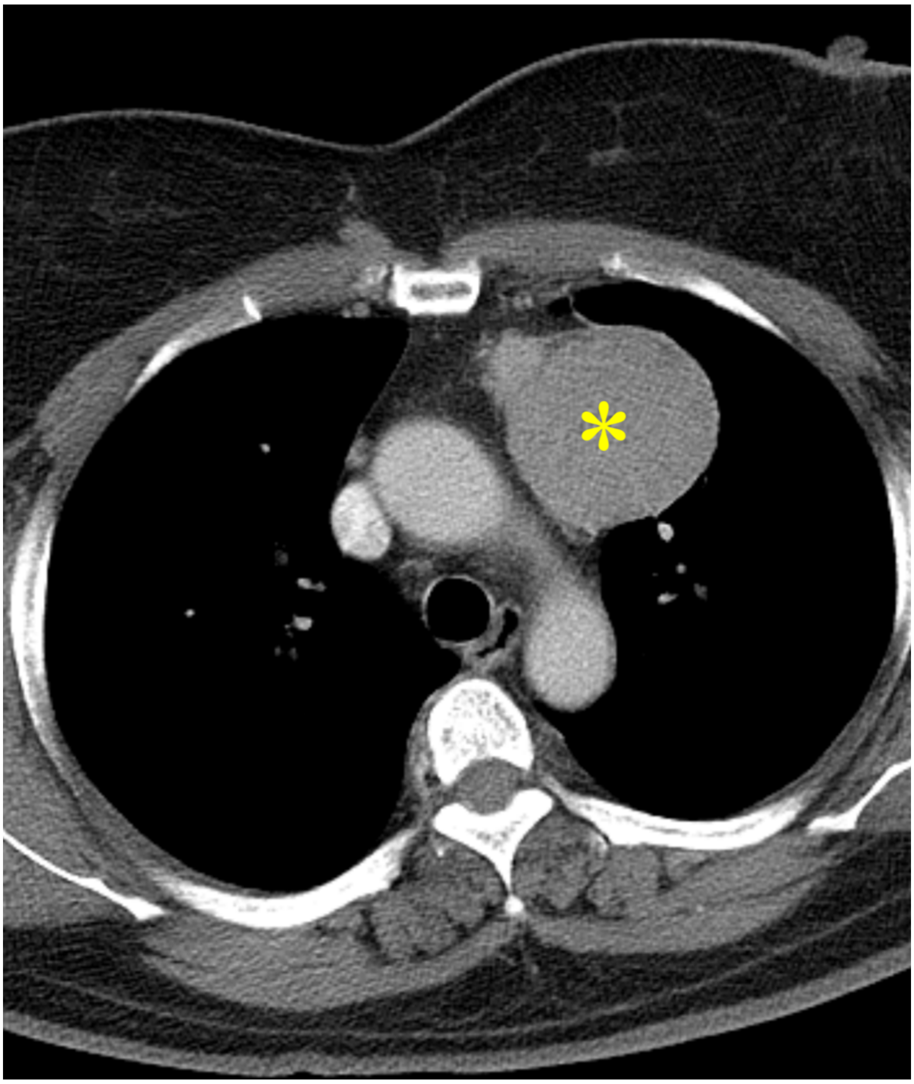

**eFigure 26. (Case 8) Dilated cardiomyopathy, negative results in a commercialized AI software of CXR**

**A.**

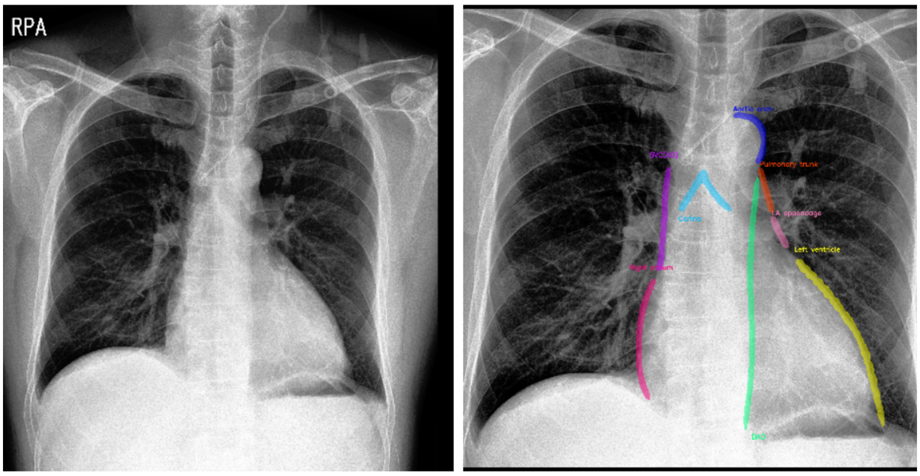

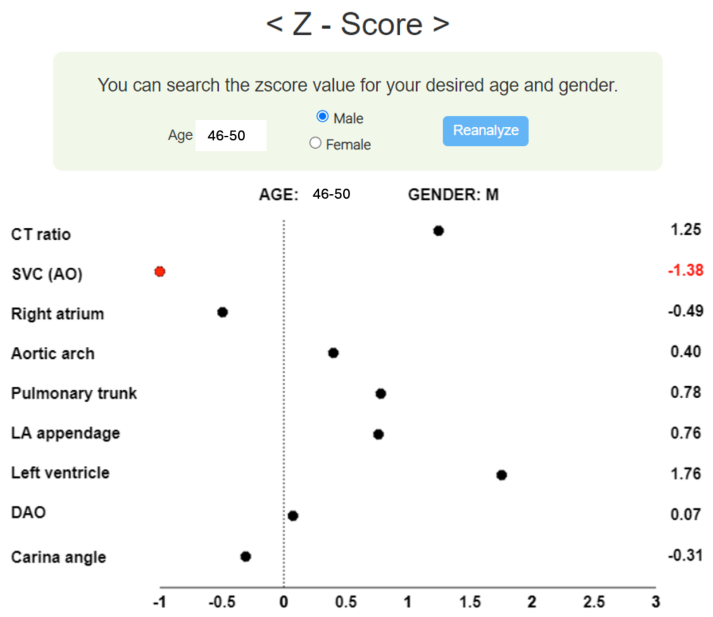

| | |
|---|---|
| Case Summary | Dilated cardiomyopathy with LV dilatation, Age range 46-50/M<br>Negative findings of cardiomegaly from a commercialized AI software for CXR interpretation. |
| Cardiac MRI | Severely dilated LV: end diastolic volume 208ml<br>Extensive mid-wall delayed enhancement suggesting dilated cardiomyopathy |
| CXR interpretation | The CT ratio (z-score 1.25) and LV (1.76) border on the CXR are increased. Based on the z-score, this patient likely has cardiomegaly. However, according to the analysis by a commercialized CXR AI software (Figure S26B), cardiomegaly was not detected in this patient. |

**B.**

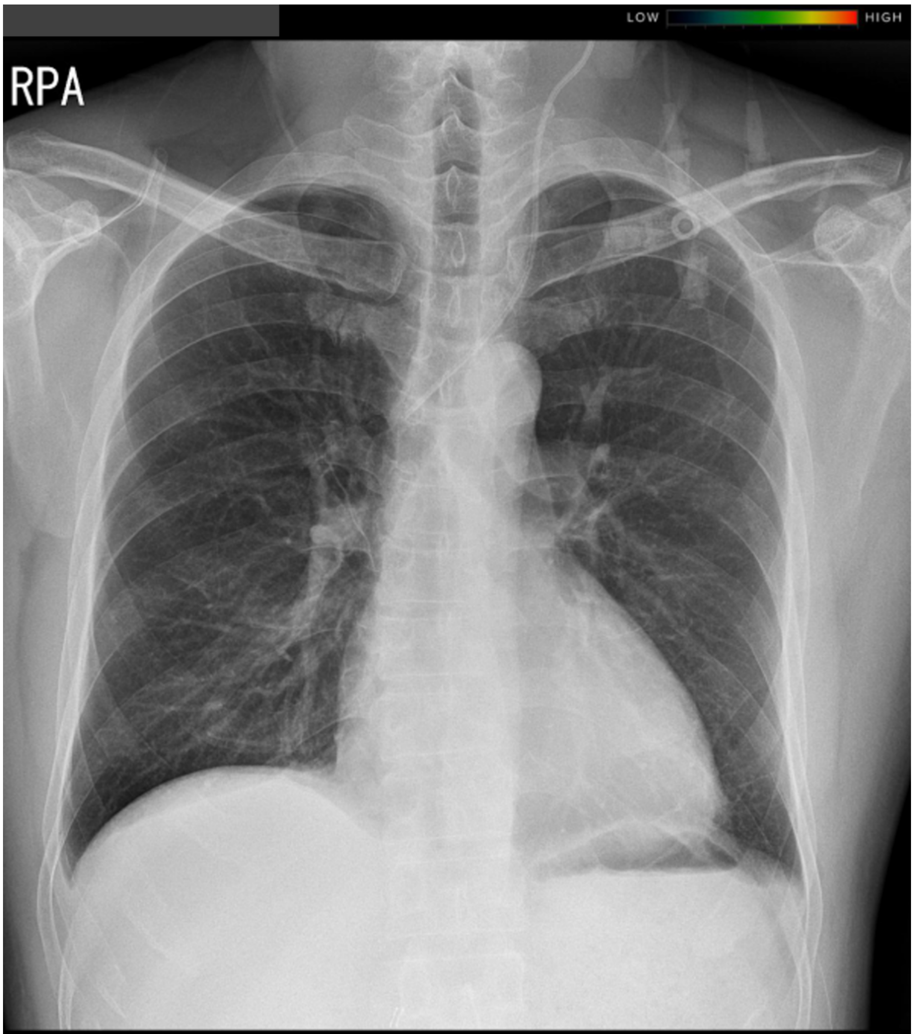

CASE REPORT

**Abnormality Score** Low

| | | | |
|---|---|---|---|
| Atl | Atelectasis | Low | - |
| Cm | Cardiomegaly | Low | - |
| Csn | Consolidation | Low | - |
| Fib | Fibrosis | Low | - |
| Ndl | Nodule | Low | - |
| PEf | Pleural effusion | Low | - |
| Ppm | Pneumoperitoneum | Low | - |
| Ptx | Pneumothorax | Low | - |

### eAppendix References

1. Ahn I, Na W, Kwon O, et al. CardioNet: a manually curated database for artificial intelligence-based research on cardiovascular diseases. *BMC Med Inform Decis Mak* 2021;21(1):29. doi: 10.1186/s12911-021-01392-2 [published Online First: 20210128]
2. Wang X, Peng Y, Lu L, et al. ChestX-Ray8: Hospital-Scale Chest X-Ray Database and Benchmarks on Weakly-Supervised Classification and Localization of Common Thorax Diseases. 2017 IEEE Conference on Computer Vision and Pattern Recognition (CVPR): IEEE Computer Society, 2017:3462-71.
3. Chexpert: A large chest radiograph dataset with uncertainty labels and expert comparison. Proceedings of the AAAI conference on artificial intelligence; 2019.
4. Cho MS, Roh JH, Park H, et al. Practice Pattern, Diagnostic Yield, and Long-Term Prognostic Impact of Coronary Computed Tomographic Angiography. *J Am Heart Assoc* 2020;9(18):e016620. doi: 10.1161/JAHA.120.016620 [published Online First: 20200908]
5. Mask r-cnn. Proceedings of the IEEE international conference on computer vision; 2017.
6. Kim C, Lee G, Oh H, et al. A deep learning-based automatic analysis of cardiovascular borders on chest radiographs of valvular heart disease: development/external validation. *Eur Radiol* 2022;32(3):1558-69. doi: 10.1007/s00330-021-08296-9 [published Online First: 20211013]
